# Integrating Common and Rare Variants Improves Polygenic Risk Prediction Across Diverse Populations

**DOI:** 10.1101/2024.11.05.24316779

**Authors:** Jacob Williams, Tony Chen, Xing Hua, Wendy Wong, Kai Yu, Peter Kraft, Xihao Li, Haoyu Zhang

## Abstract

Polygenic risk scores (PRS) predict complex traits by aggregating genetic effects across the genome, yet most models focus on common variants, overlooking rare variants that may contribute to hidden heritability. We developed RICE, a new PRS framework integrating both common and rare variants to improve genetic risk prediction across diverse ancestries. RICE constructs separate PRSs: for common variants, it integrates methods using ensemble learning; for rare variants, it uses gene-level testing with functional annotations and penalized regression. We evaluated RICE using simulated datasets and sequencing data from UK Biobank and All of Us, involving up to 740 million genetic variants from 361,939 individuals across diverse ancestries and 11 complex traits. In real data analysis, RICE improved predictive accuracy by an average of 25.7% compared to leading common variant PRS methods. Our findings demonstrate that incorporating rare variants significantly enhances PRS, providing a more accurate and inclusive approach to genetic risk prediction.

## Introduction

Polygenic risk scores (PRSs) aggregate the effect sizes of numerous genetic variants across the genome to predict an individual’s risk of developing complex traits and diseases^1–6^. PRSs have shown promise in clinical applications, enabling personalized risk prediction and informing prevention strategies for conditions such as cardiovascular diseases, type 2 diabetes, and breast cancer^7–10^. By identifying individuals at higher genetic risk, PRSs can facilitate early interventions and improve patient outcomes.

Existing PRS methods focus primarily on common genetic variants, which are well-represented in large genome-wide association studies (GWAS). Due to their higher allele frequencies, common variants provide robust effect size estimates from large-scale GWAS, supporting the development of reliable risk prediction models. Advances in genotyping technologies, imputation methods, and the availability of extensive GWAS data have contributed to the success of common variant-based PRS models. To further refine risk prediction, methods accounting for linkage disequilibrium (LD) between variants, such as clumping and thresholding^11–13^, penalization-based models^14–17^, and Bayesian approaches^18–22^, have been developed. Recent approaches that jointly model data across multiple ancestries have improved performance in diverse cohorts^23–27^.

However, rare variants (minor allele frequency [MAF] < 1%) are often excluded from PRSs due to low statistical power in single-variant association testing, historical limitations in large-scale sequencing data, limited integration methods, and challenges in accurately estimating their effect sizes. While some rare variants have larger, well-documented effect sizes (e.g., mutations in *BRCA1/2*^28^, *APOB*^29^, *PCSK9*^29,30^ or *BSN*^31^), many others tend to have modest effects that collectively contribute to disease risk^32–36^. Excluding these variants may underestimate risk, as the cumulative effect of multiple modest-effect rare variants can be significant^37,38^. Incorporating rare variants into PRSs can enhance risk prediction and provide a more comprehensive understanding of genetic risk, especially for complex traits influenced by numerous genetic factors.

Advancements in whole-genome sequencing (WGS) and whole-exome sequencing (WES) biobanks, such as UK Biobank (UKB) and All of Us Research Program (AoU), have greatly expanded opportunities for evaluating rare variants in both coding and noncoding regions^33,39–42^. To address analytical challenges posed by rare variants, methods such as the burden test, sequence kernel association test (SKAT), and variant-Set Test for Association using Annotation Information (STAAR)^43–48^ have been developed to analyze groups of rare variants collectively. Approaches like STAARpipeline leverage functional annotations to define variant sets for both coding and noncoding rare variants^47,49–51^, improving statistical power and interpretability of identified genes. Despite these advancements in association testing for rare variants, methods that simultaneously integrate rare variants into PRSs and support multi-ancestry data are notably absent.

To address these limitations, we present RICE (polygenic Risk predictions Integrating Common and rarE variants), a new framework designed for biobank sequencing data that efficiently integrates both common and rare variants. RICE combines ensemble learning techniques with advanced association testing methods to construct comprehensive PRSs that capture genetic risk from both common and rare variants. RICE significantly enhances PRS, providing a more accurate and inclusive approach to genetic risk prediction.

We evaluated RICE using extensive simulated datasets and large-scale WES and WGS data from UKB and AoU, with up to 740 million genetic variants from 361,939 independent individuals across African (AFR), Admixed American or Latino (AMR), European (EUR), Middle Eastern (MID) and South Asian (SAS) ancestries. Our analyses spanned 11 complex traits—including height, body mass index (BMI), breast cancer, and type 2 diabetes (T2D). Simulation studies demonstrated that RICE effectively models rare variant signals, achieving substantial improvements in PRS accuracy across all ancestries compared to leading common variant methods. In real data analyses, RICE improved predictive accuracy by an average of 25.7% compared to top existing methods.

## Results

### Method Overview

RICE predicts risk of complex traits and diseases by incorporating common (RICE-CV) and rare variants (RICE-RV). The framework has three steps (**Figure 1**): (1) RICE-CV combines multiple polygenic risk scores (PRSs) for common variants using ensemble learning to generate a single PRS; (2) RICE-RV identifies significant rare variant sets conditioned on RICE-CV, creates PRSs based on burden scores, and combines them via ensemble learning; (3) RICE-CV and RICE-RV PRSs are jointly evaluated in a regression model with covariates. RICE requires three datasets: (1) training for generating GWAS summary statistics, rare variant p-values, and PRS models; (2) tuning for optimizing parameters; (3) validation for assessing prediction performance.

**Figure 1.**
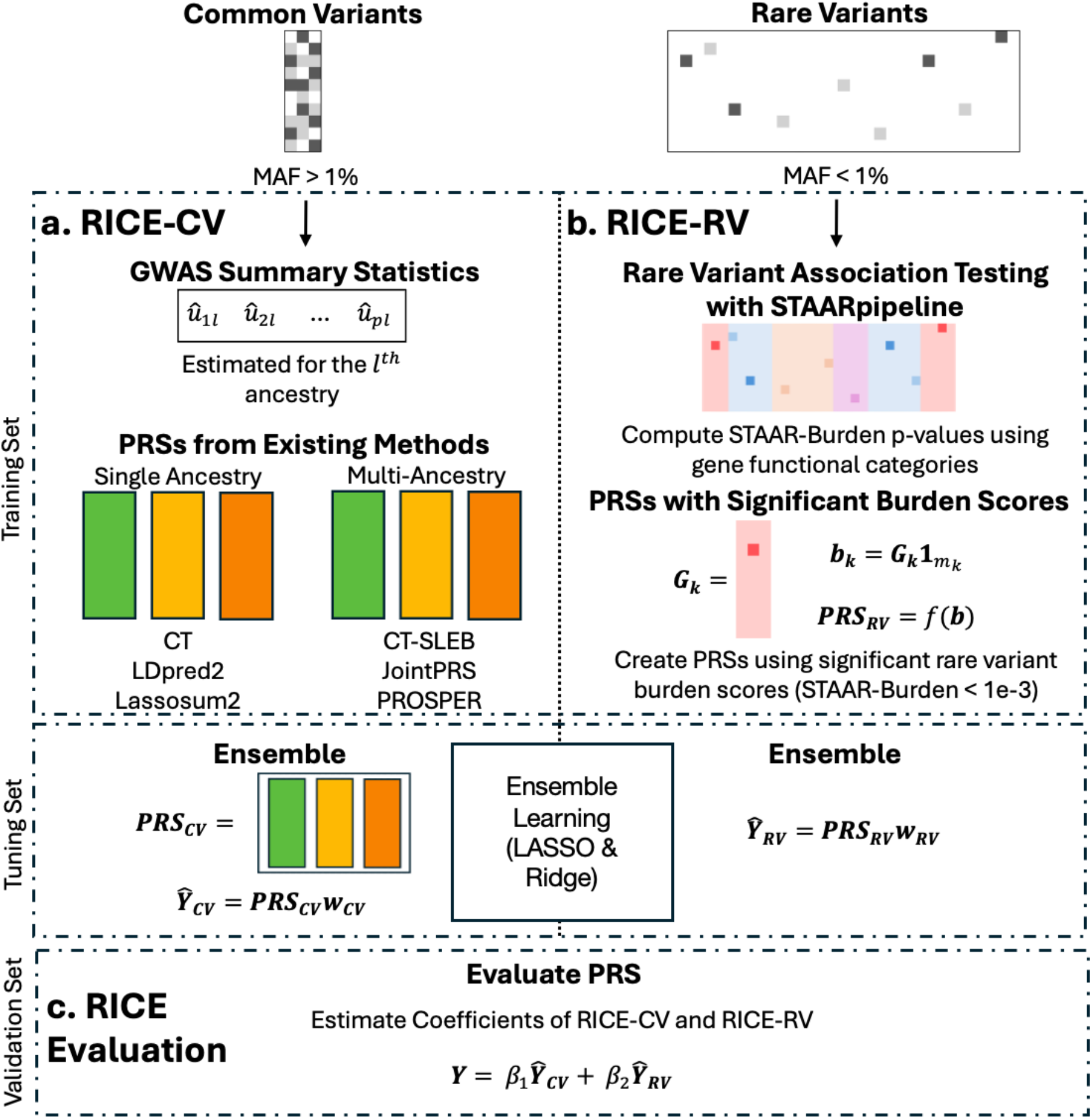
Overview of the RICE framework for polygenic risk prediction. RICE integrates common and rare variants in three steps: (a) RICE-CV (common variant PRS) is generated by combining PRSs from multiple existing methods, such as clumping and thresholding (CT), LDpred2, and Lassosum2 for single-ancestry data, or CT-SLEB, JointPRS, and PROSPER for multi-ancestry data. These PRSs are then combined using ensemble learning (LASSO and ridge regression) to produce a single robust common variant PRS. (b) RICE-RV (rare variant PRS) identifies significant rare variant sets using STAARpipeline (p-value < 10^−3^) conditioned on the RICE-CV PRS. These significant sets are collapsed into burden scores and modeled using LASSO and ridge regression. The resulting PRSs are combined through ensemble learning (LASSO and ridge regression) to create a single rare variant PRS. (c) Final evaluation: RICE-CV and RICE-RV are evaluated together in a linear or logistic regression model, adjusting for covariates such as age, sex, and principal components. The regression coefficients of RICE-CV and RICE-RV are reported separately to assess their individual contributions to predictive performance. The data are divided into three independent sets: a training set used to derive genome-wide association study (GWAS) summary statistics and perform rare variant association testing; a tuning set used to optimize model parameters and train the ensemble models; and a validation set used to evaluate the final predictive performance of the integrated PRS.

#### RICE-CV

In the first step (**Figure 1a**), RICE-CV uses the training dataset to identify common variants (MAF > 0.01 in any ancestry group) and compute ancestry-specific GWAS summary statistics. Using these summary statistics, existing PRS methods are used to generate multiple PRSs. To leverage the advantages of different methodological approaches, we select a representative method from each category: clumping-based approach, penalization-based approach and Bayesian-based approach. For single-ancestry data (e.g. primarily EUR populations), PRSs are obtained from clumping and thresholding (CT)^11–13^, LDpred2^52^, and Lassosum2^16^. For multi-ancestry data, PRSs are obtained from CT-SLEB^24^, JointPRS^53^, and PROSPER^27^. After computing PRSs from different approaches under various tuning parameters, RICE-CV performs ensemble learning using the SuperLearner^54^ package to combine all PRSs into a single robust common variant PRS, using LASSO and ridge regression as base learners.

#### RICE-RV

In the second step (**Figure 1b**), RICE-RV uses the training set to fit a baseline model adjusting for covariates, including top 10 principal components (PCs), sex, age, age squared, and RICE-CV to prioritize rare variant signals independent of common variant effects. Using residuals from this baseline model, RICE-RV performs variant set analyses with STAARpipeline to identify significant rare variant sets^49^, which defines rare variant sets using functional categories within each protein-coding gene. In WES datasets, RICE-RV conducts gene-centric analysis with rare variants in coding region only. In WGS datasets, RICE-RV analyzes rare variants in both coding and noncoding regions. Significant rare variant sets are slected as those with a STAAR-Burden p-value less than 1 × 10^−3^. These sets are then collapsed into burden scores, which approximately represent the combined allele frequencies (**Methods**). These burden scores are jointly modeled using LASSO and ridge regression under different tuning parameters. PRSs are then generated on the tuning dataset using these burden scores and weights estimated from different penalized regression models. Finally, the PRSs are combined through ensemble learning via the SuperLearner ^54^ package, with LASSO and ridge regression as base learners, to create a single rare variant PRS.

#### RICE Evaluation

In the final step (**Figure 1c**), RICE-CV and RICE-RV are jointly evaluated on the validation set using a linear or logistic regression model, adjusting for covariates including top 10 PCs, sex, age and age squared. The regression coefficients of RICE-CV and RICE-RV are reported separately as measures of predictive performance.

### Evaluation of Prediction Performance

PRSs are traditionally evaluated using metrics like the coefficient of determination (*R*^2^) for continuous traits or area under the receiver operating characteristic curve (AUC) for binary traits^2^. These measures assess how well the PRS explains variation in a trait on average across the entire population. However, because rare variants are present in only a relatively small subset of individuals, their impact may not significantly influence these average-based metrics. Despite this, rare variants can have substantial effects on individuals who carry them, which may be overlooked when using traditional evaluation methods.

To better capture the contribution of rare variants, we propose a new evaluation metric: the regression coefficient of the standardized PRS on the standardized trait (**Methods**). This coefficient provides a direct measure of the effect size of both common and rare variant PRSs on the trait. Notably, the estimated regression coefficient can be shown to correspond to the square root of the heritability explained by the PRS (**Supplementary Note**). This approach focuses on the effect size of the PRS in predicting the trait, providing a more nuanced assessment of predictive performance, especially for rare variants.

Figure 2 illustrates the estimated effect coefficients of the standardized PRS for both RICE-CV and RICE-RV using high-density lipoprotein cholesterol (HDL) levels from the UKB WGS dataset. For RICE-CV, a one-unit increase in the PRS is associated with a 0.388 standard deviation (SD) increase in HDL levels (β_1_ = 0.388), while a one-unit increase in the RICE-RV PRS corresponds to a 0.111 SD increase (β_2_ = 0.111). Notably, the distribution of RICE-RV PRS values is more dispersed compared to RICE-CV, with approximately 0.6% of individuals exhibiting PRS values five to ten units above the population mean. This demonstrates that while rare variants affect a smaller segment of the population, their impact on those individuals can be substantial.

### Simulation Study Results

We evaluated RICE’s ability to detect and predict rare variant effects using extensive simulation studies based on UKB WES data with 140,080 unrelated individuals (**Methods**). Genetic ancestries were inferred using 1000 Genomes Phase 3 Project 3 data (1000G, **Methods**)^55^. Since over 90% of individuals were of EUR ancestry, we designed simulations with an EUR-only training dataset (N = 98,343), while the tuning (N = 20,869) and validation datasets (N = 20,868) included individuals from AFR, AMR, EUR, and SAS ancestries (sample size details in **Supplementary Table 1**).

**Figure 2.**
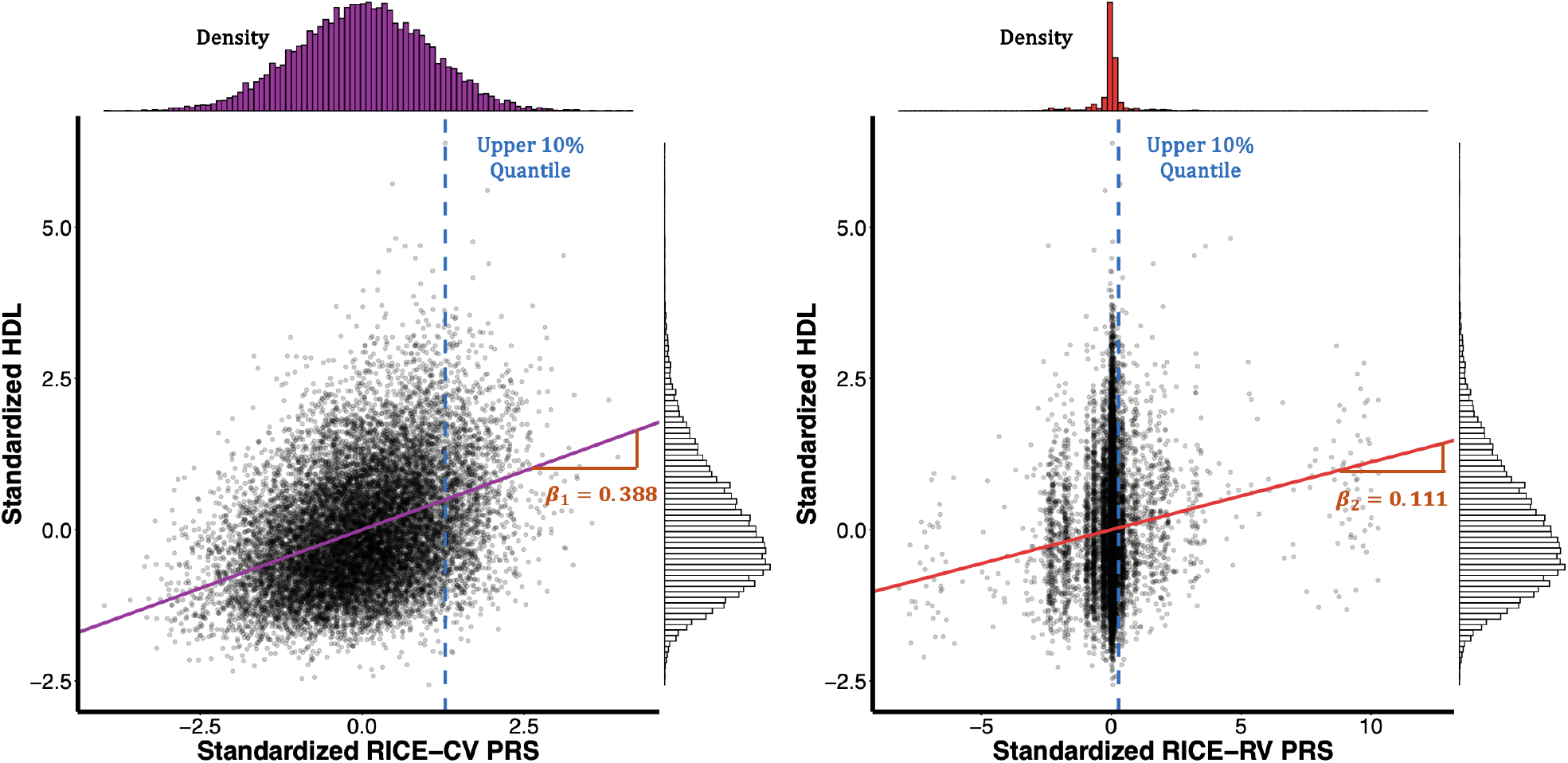
Comparison of standardized PRSs from RICE-CV and RICE-RV for high-density lipoproteins cholesterol (HDL). Standardized PRSs for HDL were computed using UK Biobank whole genome sequencing data for 13,839 individuals of European ancestry from the validation dataset. The left panel shows the standardized observed HDL levels plotted against the standardized PRS from RICE-CV (common variants), while the right panel shows the same comparison using RICE-RV (rare variants). In both figures, the blue dashed line represents the upper 10% quantile cutoff. The estimated regression coefficients (*β*_1_ for RICE-CV and *β*_2_ for RICE-RV) are displayed in orange.

RICE-CV was constructed by applying single-ancestry common variant PRS methods: CT, LDpred2, and Lassosum2 to the EUR training data. We compared the performance of RICE-CV and RICE-RV to these methods across different ancestries. The rare variant analysis was conducted using STAARpipeline with protein-coding genes within coding regions. RICE was evaluated across various simulation settings, including different causal variants proportion (0.01, 0.05, and 0.2), training sample size (N = 49,173 and 98,343) and effect size distributions (strong negative selection and no negative selection) (**Methods**), RICE-CV consistently demonstrated robust performance (**Figure 3 and Supplementary Figure 1**). By leveraging the strengths of different common variant PRS methods, RICE-CV provides a robust PRS that slightly outperformed the best existing PRS methods across all ancestries, achieving an average increase of 1.5% in the beta coefficient compared to the best existing approach.

**Figure 3.**
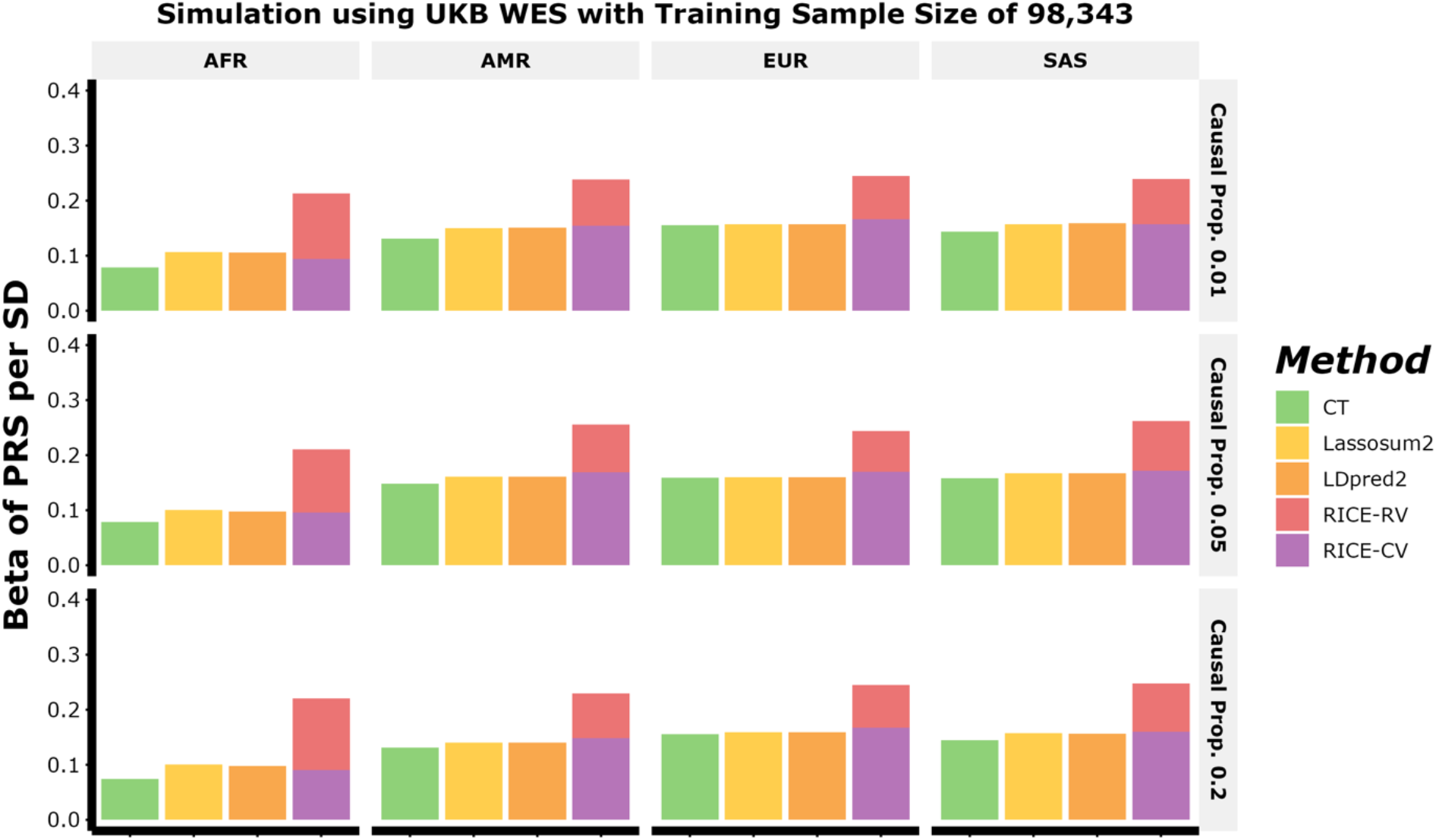
Simulation results comparing the predictive performance of PRSs for four ancestral groups from the UK Biobank (UKB). The training data (N = 98,343) included only individuals of European ancestry (EUR), while the tuning (N = 20,869) and validation datasets (N = 20,868) contained individuals of African (AFR), Admixed American or Latino (AMR), European (EUR), and South Asian (SAS) ancestries (**Supplementary Table 1**). Simulations assumed a common variant heritability of 0.05 and a rare variant set heritability of 0.0125, under the assumption of no negative selection effect size distribution (**Methods**). Causal proportions for both common variants and rare variant sets varied across three levels: 0.01 (top), 0.05 (middle), and 0.2 (bottom). Data were generated using unrelated individuals from UK Biobank whole-exome sequencing data (WES), with simulation based on chromosome 22. PRS performance is reported as the “Beta of PRS per standard deviation (SD)”, derived from the regression model *Y* ∼ *PRS* × *β*, with *β* representing the effect of standardized PRS on the standardized outcome (**Methods**). For RICE, the model used was *Y* ∼ *PRS*_*CV*_ × *β*_*CV*_ + *PRS*_*RV*_ × *β*_*RV*_ Beta values can be interpreted as the square root of the heritability 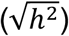 of the outcome explained by the PRS (**Supplementary Note**).

RICE-RV also performed strongly, effectively identifying causal rare variant signals and predicting their effects across both EUR and non-EUR populations (**Figure 3 and Supplementary Figure 1**). On top of RICE-CV, RICE-RV contributed an additional average increase of 53.2%, 32.4%, 28.4%, and 32.4% in predictive accuracy for AFR, AMR, EUR, and SAS, respectively, as measured by the effect size per SD of PRS. Notably, RICE-RV showed particularly strong performance in AFR individuals, as the randomly selected causal variants and rare variant sets across the genome led to higher average burden scores (combined allele frequencies) in this ancestry (**Supplementary Figure 2**), driving more notable rare variant prediction performance. Overall, RICE-CV achieved the highest prediction accuracy among common variant PRSs across all populations in most scenarios, while RICE-RV correctly identified causal rare variants and accurately predicted their impact.

### UKB WES and WGS Results

We applied RICE to compute PRSs for 11 complex traits using UKB WES and WGS data. The analyses included six continuous traits: BMI, HDL, height, low-density lipoprotein (LDL), log-transformed triglycerides (log(TG)), and total cholesterol (TC), and five binary traits: asthma, breast cancer, coronary artery disease (CAD), and T2D. Training data included 98,103 EUR individuals, while tuning (N = 20,869) and validation (N = 20,868) datasets included individuals from AFR, AMR, EUR, and SAS ancestries (sample size details in **Supplementary Table 2-3**). For both WES and WGS, we conducted association analyses as follows: for common variants, we adjusted for top 10 PCs, sex, age and age squared. For rare variant association testing, we further included RICE-CV as an additional covariate to ensure independence between RICE-RV and RICE-CV (**Methods**). The genomic inflation factor was well controlled, with λ_1000_ ranging between 1 to 1.008 (**Supplementary Table 4**).

#### UKB WES Results

Across six continuous traits, RICE-CV consistently achieved the highest performance among common variant PRS methods for almost all traits, with an average increase of 1.5% in the estimated effect size of the standardized PRS compared to the best alternative approach (**Figure 4, Supplementary Table 6**). In EUR individuals, RICE-RV contributed substantially to the prediction of lipid traits such as HDL, LDL, log(TG), and TC, adding approximately 27.0%, 29.4 %, 26.4%, and 30.4% to the respective RICE-CV effect sizes. The improvements from RICE-RV in lipid traits were driven by significant contributions from genes like *APOC3, APOB*, and *LDLR*, which had substantial weights in our model (e.g., *APOC3* weight for HDL = 0.144, 2.4 standard deviations above the average effect size; full model weights are provided in **Supplementary Data**), reinforcing the accuracy of the predictions^56–58^. However, BMI and height did not show noticeable improvement when adding rare variants in EUR individuals. For binary traits, the performance of binary traits showed limited improvement (**Supplementary Figure 3**), likely due to smaller case numbers (**Supplementary Table 2**).

**Figure 4.**
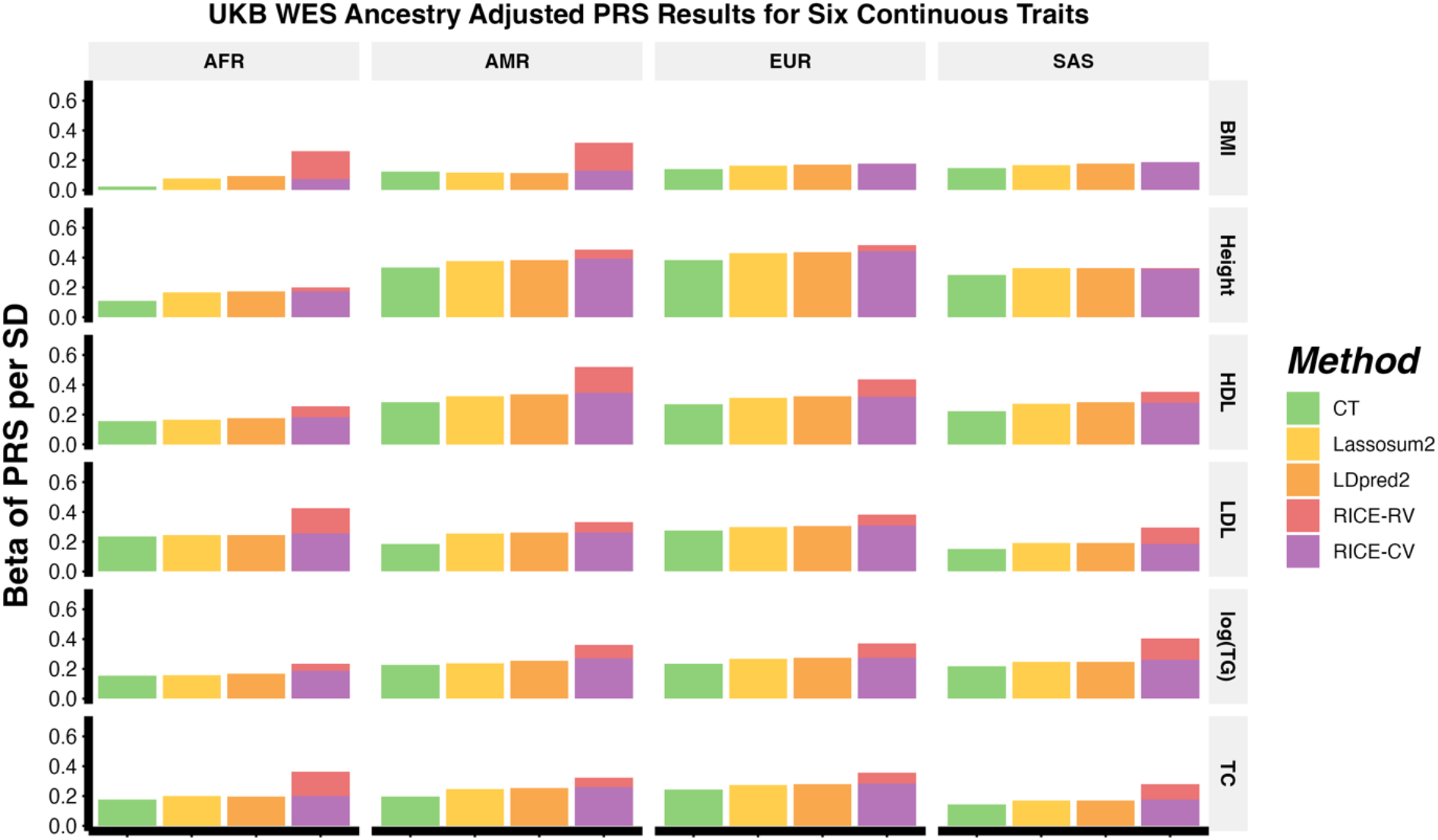
Predictive performance of ancestry-adjusted PRSs for continuous traits across four ancestral groups from UK Biobank (UKB) whole-exome sequencing (WES) data. The traits analyzed include body mass index (BMI), height, high-density lipoproteins cholesterol (HDL), low-density lipoproteins cholesterol (LDL), the natural logarithm of triglyceride cholesterol (log(TG)), and total cholesterol (TC). Results are shown for individuals of African (AFR), Admixed American or Latino (AMR), European (EUR), and South Asian (SAS) ancestries. The training data consisted solely of individuals of European ancestry, while tuning and validation sets included all four ancestries. Full sample sizes details for each ancestry are provided in **Supplementary Table 2**.

**Figure 5.**
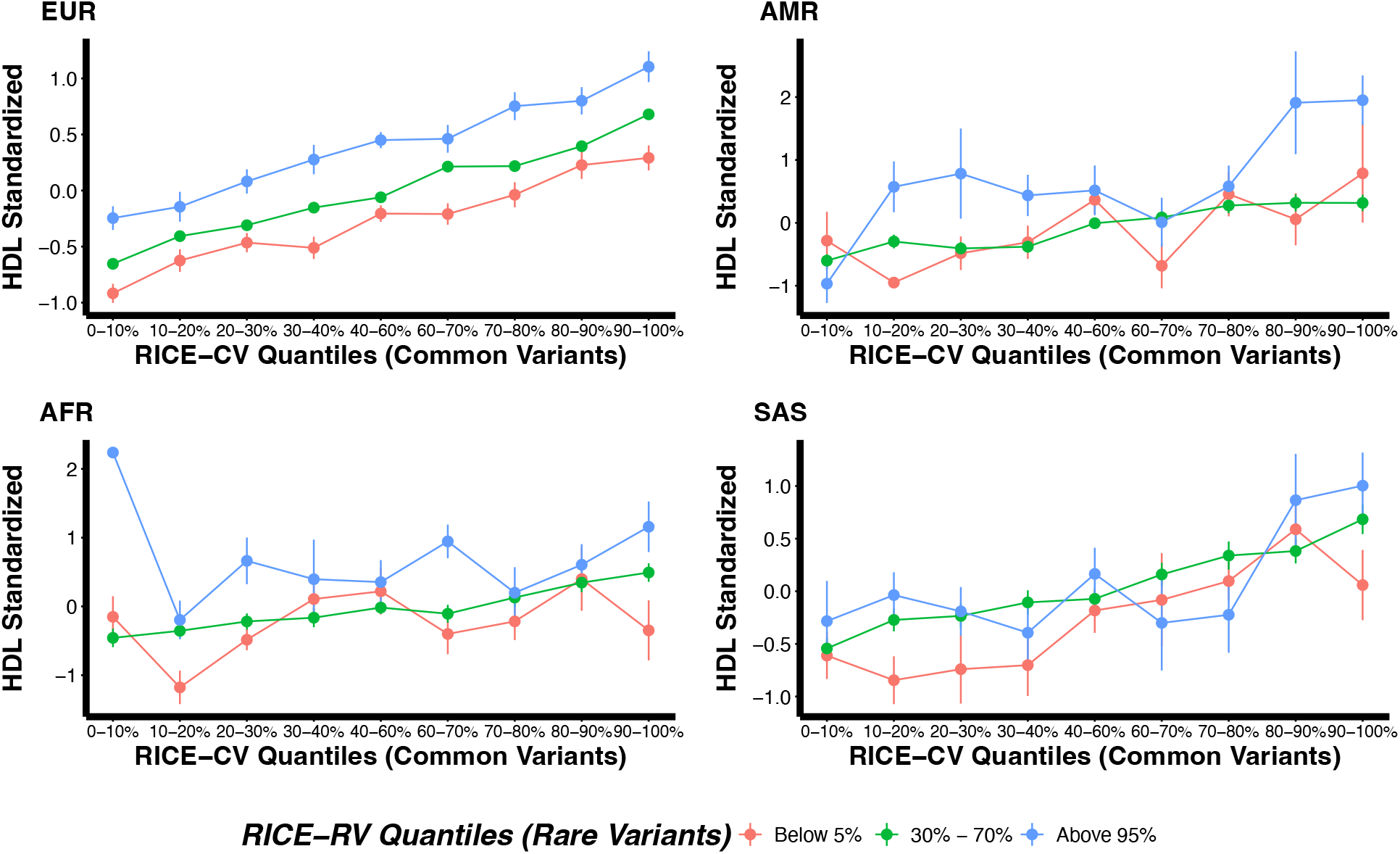
Relationship between ancestry-adjusted common and rare variant PRSs and standardized high-density lipoprotein cholesterol (HDL) levels across four ancestral groups from UK Biobank (UKB) whole-genome sequencing (WGS) data. PRS quantiles for RICE-CV (common variants) are plotted on the x-axis, and standardized HDL levels are on the y-axis. Data are stratified by rare variant PRS quantiles from RICE-RV (red: below 5%, green: 20–70%, blue: above 95%). Results are shown for individuals of African (AFR), Admixed American or Latino (AMR), European (EUR), and South Asian (SAS) ancestries. The training data consisted solely of individuals of European ancestry, while the tuning and validation sets included all four ancestries. Full sample sizes details for each ancestry are provided in **Supplementary Table 3**.

We tested two different approaches for standardizing PRSs: (1) adjusting for PCs using a regression-based approach (**Methods**), and (2) standardizing within each genetically inferred ancestry group using ancestry-specific means and standard deviations. The performance of the two approaches was consistent (**Supplementary Figure 4**); therefore, we used the regression-based approach to avoid categorizing individuals into ancestry groups when standardizing PRSs.

In continuous traits like HDL and log(TG), significant differences were observed between RICE-RV quantiles (below 5%, 30%–70%, and above 95%). In EUR individuals, those in the lowest RICE-RV quantile had notably lower standardized trait values, with average decreases of 0.35 units for HDL and 0.22 units for log(TG) compared to the middle group. Conversely, individuals in the highest quantile had significantly higher trait values, with average increases of 0.35 units for HDL and 0.18 units for log(TG), respectively (**Supplementary Figures 6b and 6e**). This suggests that rare variants can have both protective and deleterious effects, influencing an individual’s risk status.

**Figure 6.**
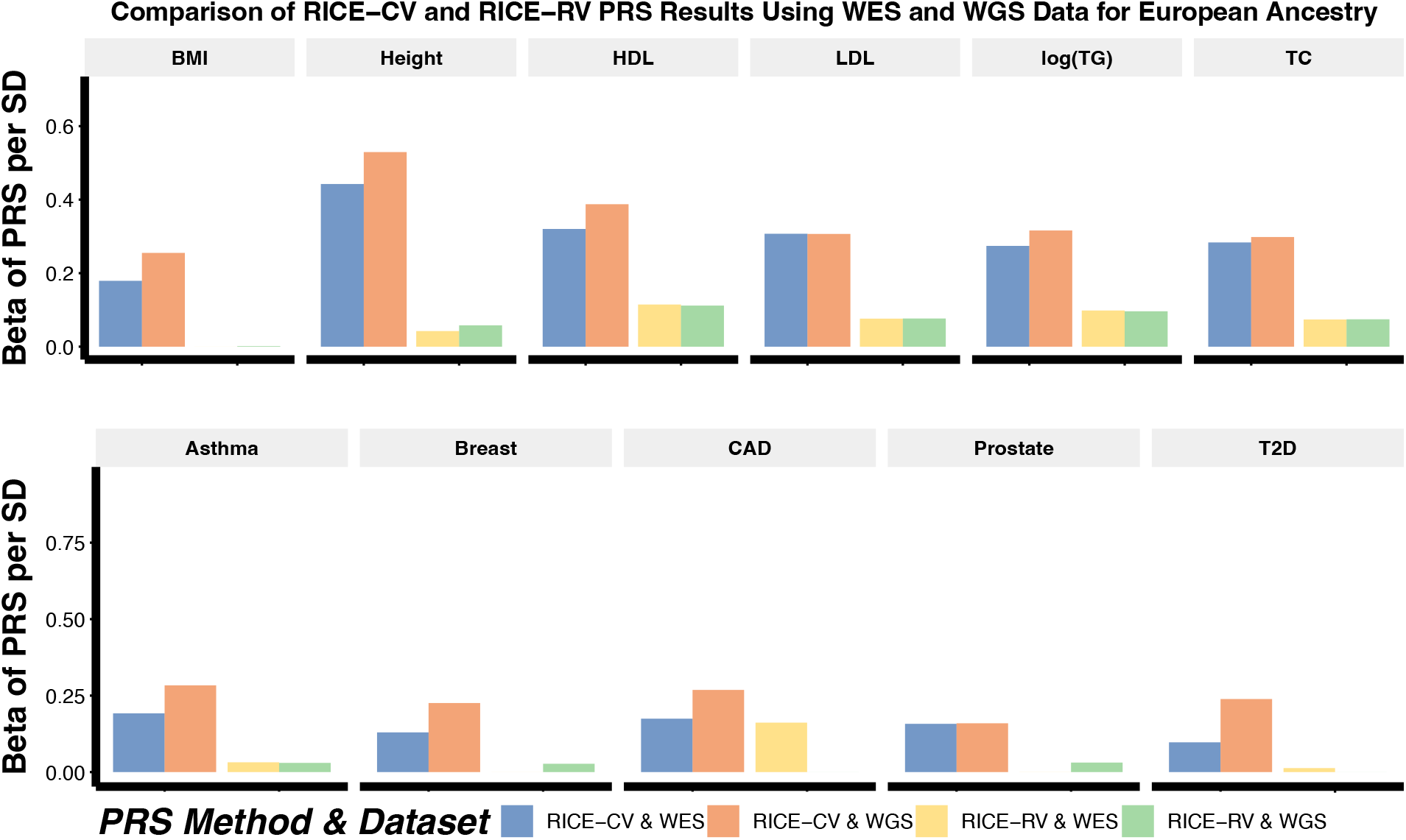
Comparison of ancestry-adjusted PRSs from RICE-CV and RICE-RV using UK Biobank (UKB) whole-exome sequencing (WES) and whole-genome sequencing (WGS) data for individuals of European ancestry. Results are shown for six continuous traits: body mass index (BMI), height, high-density lipoproteins (HDL), low-density lipoproteins (LDL), the natural logarithm of triglyceride (log(TG)), and total cholesterol (TC), and five binary traits: asthma, breast cancer, coronary artery disease (CAD), prostate cancer, and type 2 diabetes (T2D). The training data included only individuals of European ancestry, while the tuning and validation sets contained individuals from all four ancestries. Full sample size details for each ancestry are provided in **Supplementary Tables 2 and 3**.

Quality control assessments for UKB WES association analyses were performed using Manhattan and QQ plots, confirming well-calibrated p-values and controlled inflation factors (**Supplementary Figures 7–9)**

**Figure 7.**
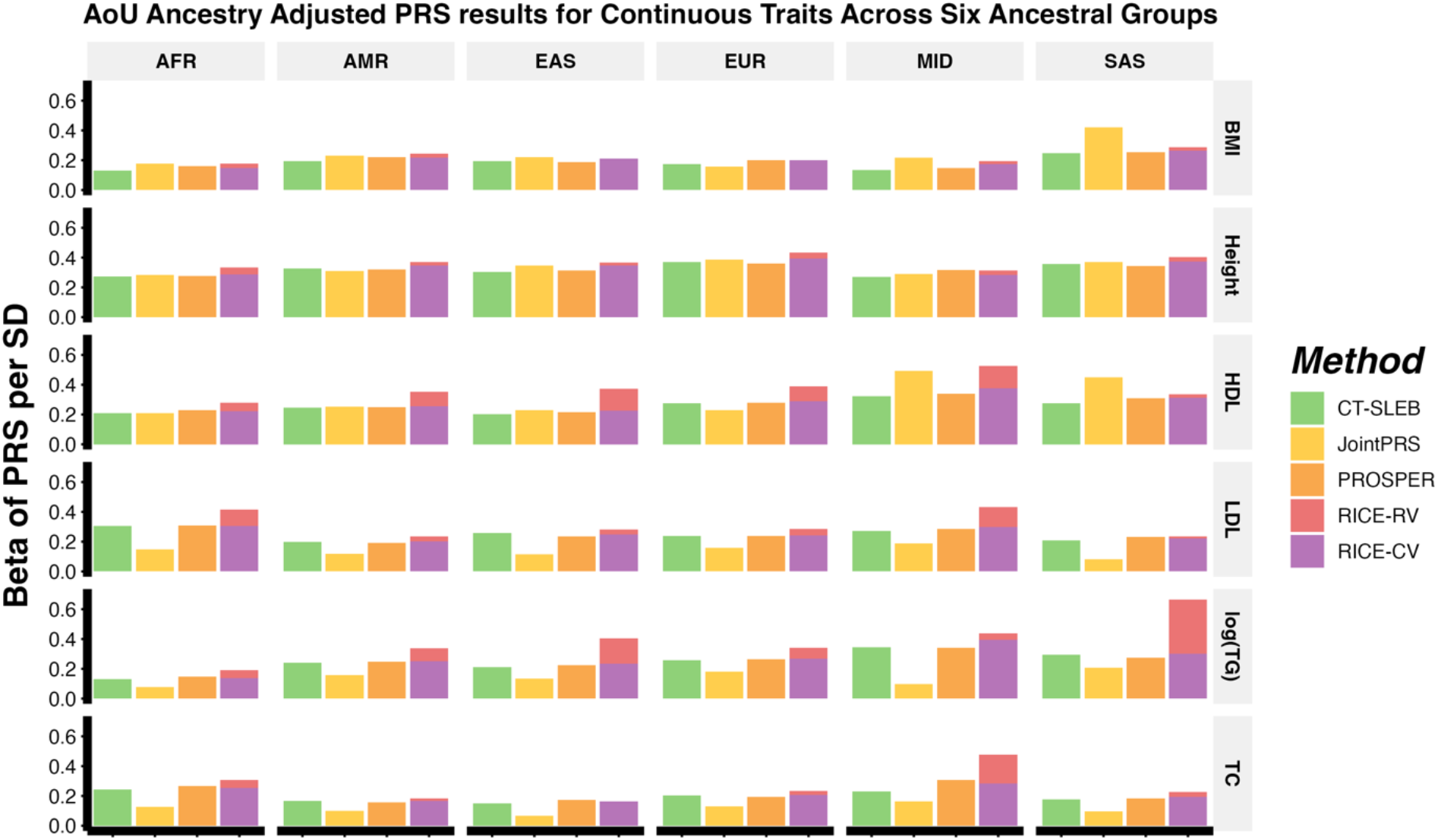
Predictive performance of ancestry-adjusted PRSs for continuous traits across six ancestral groups from the All of Us (AoU) whole-exome sequencing (WES) data. The traits analyzed include body mass index (BMI), height, high-density lipoproteins (HDL), low-density lipoproteins (LDL), the natural logarithm of triglyceride (log(TG)), and total cholesterol (TC). Results are shown for individuals of African (AFR), Admixed American or Latino (AMR), East Asian (EAS), European (EUR), Middle Eastern (MID), and South Asian (SAS) ancestries. Training data consisted of individuals of EUR, AFR and AMR ancestry, while tuning and validation sets included individuals from all six ancestries. Full sample size details for each ancestry and dataset are provided in **Supplementary Table 4**.

#### UKB WGS Results

Similar trends were observed in WGS data. RICE-CV consistently provided robust predictive accuracy, often slightly exceeding existing common variant PRS methods. RICE-RV added noticeable predictive value for lipid traits but showed little to no improvement for BMI (**Supplementary Figure 10**). Significant differences in lipids traits were revealed by stratification based on RICE-RV quantiles in European individuals (**Figure 5, Supplementary Figure 12c, 12d, and 12e**). For EUR individuals, stratifying lipid traits by RICE-RV quantiles revealed significant differences in standardized HDL values, with those in the lowest quantile showing an average decrease of 0.26 units, and those in the highest quantile showing an average increase of 0.40 units compared to the middle grouping (**Figure 5**). Incorporating RICE-RV helped to identify individuals whose trait values were significantly impacted by rare variants, which would have been missed by common variant PRSs alone. However, low sample sizes for non-European ancestries in the validation dataset made it challenging to discern general trends for these groups.

#### Comparison between WGS and WES Results

The substantially increased number of variants in the WGS data compared to WES data significantly enhanced the predictive performance of RICE-CV across all ancestries (**Figure 6, Supplementary Figure 13, Supplementary Table 9**). Specifically, increasing the number of common variants from 137,466 in WES to around 5.47 million in WGS improved RICE-CV’s performance, with average increases of 20.2%, 11.7%, 15.9%, and 20.1% for continuous traits in AFR, AMR, EUR, and SAS individuals, respectively and 0.11%, 18.6%, 27.8%, and 40.2% for all traits for AFR, AMR, EUR, and SAS individuals, respectively. For RICE-RV, however, increasing the number of rare variants from approximately 17 million in WES to 734 million in WGS did not lead to a noticeable improvement in predictive performance (**Figure 6, Supplementary Figure 13**). The lack of improvement may be due to rare variant signals being predominantly located in coding regions, with variants located in noncoding regions contributing less independent signal. A further breakdown of RICE-RV results by coding and noncoding regions revealed that variants in noncoding regions provided limited predictive signal compared to coding region variants (**Supplementary Figure 14**).

Quality control for the association analyses of UKB WGS data was assessed with Manhattan and QQ plots, showing appropriate p-value distributions and controlled inflation (**Supplementary Figures 15–18**).

### All of Us Results

We applied RICE and compared its performance against other existing multi-ancestry PRS methods across six continuous traits in the AoU WGS dataset (**Methods, Supplementary Table 5, Supplementary Figures 19**). Covariate adjustments followed similar procedures as UKB analyses. The training dataset (N = 155,611) consisted of individuals from AFR, AMR, and EUR ancestries, while the tuning (N = 33,342) and validation (N = 33,363) datasets included AFR, AMR, EAS, EUR, MID, and SAS populations (sample size details in **Supplementary Table 4**). Common variants were defined as those with MAF > 0.01 in any of the three ancestries in the training dataset. Rare variant analyses were constrained to the exome region for computational efficiency (**Methods**). The genomic inflation factor was well controlled, with λ_1000_ ranging from 1 to 1.002 (**Supplementary Table 4**).

RICE-CV and RICE-RV demonstrated similar performance patterns to those observed in the UKB analysis (**Figure 7**). RICE-CV consistently demonstrated robust performance across almost all traits and ancestries, often outperforming or matching existing methods. However, in MID individuals for HDL and SAS individuals for BMI, RICE-CV performed lower than JointPRS, likely due to smaller validation sample sizes in these populations. RICE-RV contributed significantly to lipid trait predictions, especially HDL and log(TG). For example, RICE-RV achieved 25.8% of RICE-CV’s prediction performance for HDL in EUR individuals and an average of 24.2% in non-EUR ancestries; Similarly, for log(TG), RICE-RV reached 22.0% of RICE-CV’s predictive power in EUR individuals and 32.1% across non-EUR ancestries. These results indicate that rare variants located in coding regions can substantially enhance predictive performance for individuals with significant rare variants affecting these traits across all ancestries.

With larger sample sizes for AFR and AMR individuals in AoU compared to UKB, clear distinctions were observed when comparing the bottom 5%, middle 30–70%, and top 95% quantiles of RICE-RV scores for height and lipid traits (**Supplementary Figure 19b–f**). Individuals in the top 95% quantile had standardized trait values that were on average 0.14 units higher than those in the middle group and 0.32 units higher than those in the bottom 5% quantile. However, separations in EAS, SAS, and MID individuals were less pronounced due to smaller sample sizes (**Supplementary Figure 19b–f**). Consistent with UKB results, BMI showed no clear predictive signal from rare variants (**Supplementary Figure 19a**).

Quality control for the association analyses of AoU data was assessed with Manhattan and QQ plots, confirming appropriate p-value distributions and controlled inflation (**Supplementary Figures 20–21**).

## Discussion

Understanding the role of rare genetic variants is crucial for unraveling the genetic architecture of complex traits. We introduced RICE, a new PRS framework that integrates both common and rare genetic variants to enhance genetic risk prediction across diverse ancestries. Using large-scale sequencing data from UKB and AoU studies, RICE significantly improves predictive accuracy, achieving an average increase of 25.7% compared to leading common variant PRS methods across traits, and enables more precise risk stratification across multiple traits and populations.

Our findings showed that RICE-CV, the common variant component, consistently delivered robust predictive accuracy, often slightly exceeding or matching existing common variant PRS methods across ancestries and traits (**Figure 4, Figure 7, Supplementary Figures 3 and 10**). By combining multiple PRS approaches, RICE-CV captured a broader spectrum of polygenic signals. This ensemble framework highlights that while no single method is best for all scenarios^24,27,53^, integrating diverse approaches can optimize predictions across populations.

Importantly, RICE-RV enhanced predictive power by incorporating rare variants, particularly improving risk prediction for lipid-related traits, while showing limited gains for BMI (**Figure 4, Figure 7, Supplementary Figures 3 and 10**). This aligns with previous heritability studies indicating that rare variants contribute more significantly to certain traits^38^. For instance, Weiner et al^38^. estimated that the ratio between exome burden heritability and common variant heritability was approximately 0.231 for LDL, but only 0.048 for BMI. Including rare variants enabled us to identify individuals with extreme trait values.

Traditional evaluation metrics like *R*^2^ may underestimate the impact of rare variants because they focus on average effects across the population. To better capture their contribution, we introduced a new evaluation metric: the regression coefficient of the standardized PRS on the standardized trait (**Figure 2**). This provides a direct measure of the PRS effect size, offering a nuanced assessment of predictive performance, especially for rare variants. Our analyses demonstrated that, despite affecting fewer individuals, rare variants can have significant effects on trait values, reinforcing the importance of including them in PRS models.

One unique contribution of our study is the comprehensive assessment of rare variants’ contributions across large and diverse datasets. Rare variants are often overlooked in risk prediction models due to complexity and challenges in modeling them^1,2^. Existing efforts have focused on combining high-penetrance genes with common variant PRSs^59–61^, as seen in breast cancer risk prediction models that include genes such as *BRCA1, BRCA2, PALB2, CHEK2*, and *ATM*^59^. While these approaches effectively integrate high-effect genes, they may miss modest-risk rare variant sets. By incorporating functional annotations using the STAARpipeline^49^, RICE-RV can identify novel rare variant sets beyond those described in existing literature, leveraging larger sample sizes and diverse ancestries to capture effects of rare variants across functionally distinct regions (e.g., predicted loss-of-function (pLoF) or deleterious missense variants). This enables us to capture the varying effect sizes of rare variants across functional regions, as shown in previous studies where loss-of-function variants often exhibit large effects^62–65^.

Our results further indicated that rare variant signals contributing to PRS were largely captured in coding regions, while the contribution of rare variants in noncoding regions was comparatively weaker (**Figure 6**). This observation aligns with findings from recent association testing studies^66,67^. For example, Gaynor et al.^66^ reported that the gene-based association signals from WGS data differed by only 1% from those derived from WES plus imputation, based on analyses of 100 complex traits in UKB data. Similar conclusions were reached^67,68^, which demonstrated that rare variants in coding regions, particularly pLoF variants, are significantly enriched for disease associations compared to synonymous variants. While coding regions appear to harbor most of the rare variant signal, rare variants in noncoding regions may still offer additional insights, albeit it remains challenging to quantify their effects^69–71^. These findings have important implications for optimizing the design of genetic studies and the development of risk prediction tools, particularly when prioritizing between WES and WGS data.

An important feature of RICE to provide a single PRS for common variants applicable across ancestries. Existing methods like CT-SLEB^24^ and PROSPER^27^ are often optimized for specific ancestries, requiring ancestry-specific tuning. While these methods may achieve high performance in a target ancestry, they complicate clinical translation by necessitating prior knowledge of a patient’s genetic ancestry. In contrast, RICE-CV uses a mixed-ancestry tuning dataset that combines results from multiple methods, yielding a single PRS applicable across ancestries. Our results showed that this approach achieved higher or comparable performance to the best alternative methods in nearly all traits and ancestries (**Figure 7**), simplifying its potential clinical utility.

Our analyses demonstrated that incorporating rare variants in the coding region enhances the prediction of lipid traits, with genes like *APOC3, APOB*, and *LDLR* emerged as key contributors in our models (**Supplementary Data**), consistent with existing findings in lipid metabolism^72–74^. For example, rare loss-of-function variants in *APOC3* have been associated with lower triglyceride levels and reduced coronary artery disease risk^74^. This alignment with known biological mechanisms reinforces the validity of our results, highlighting the relevance of these genes in clinical risk assessment and therapeutic strategies, while also offering new opportunities for biological insights.

Despite the promising results, our study has several limitations. First, collapsing rare variant sets assumes equal weighting of all variants, which may overlook variant-specific differences in effect sizes and direction. Incorporating variable weighting based on functional annotations or predicted pathogenicity could enhance RICE-RV’s performance^47–49,75–78^. Second, while RICE-RV demonstrated consistent results in simulations, its performance in real data analyses was more variable, likely reflecting the complex nature of rare variant effects and interactions with environmental factors. Third, smaller sample sizes for certain ancestries may impact model stability and generalizability. Increasing representation in genetic studies is crucial for enhancing PRS accuracy and supporting equitable healthcare applications^79^.

In conclusion, we developed RICE, a PRS framework that integrates both common and rare variants within a single model to improve genetic risk prediction across diverse ancestries. Our study demonstrates that incorporating rare coding variants enhances the predictive power of PRSs for lipid-related traits, particularly for individuals carrying these variants who may experience significant impacts on trait values. By providing open-source software for RICE (https://github.com/jwilliams10/RICE), we offer a practical tool to advance polygenic risk prediction and contribute to precision medicine.

## Data Availability

All data produced in the present work are contained in the manuscript.

## Acknowledgements

The analysis utilized the high-performance computation Biowulf cluster at National Institutes of Health, USA. This research has been conducted using the UK Biobank Resource under Application Number 52008. We gratefully acknowledge All of Us participants for their contributions, without whom this research would not have been possible. We also thank the National Institutes of Health’s All of Us Research Program for making available the participant level data examined in this study. We want to thank Jin Jin, Jingning Zhang, and Xiaoyu Wang for sharing their code for PRSs approaches.

## Author Contributions Statement

J.W., X.L., and H.Z. conceived the project. J.W. carried out all data analyses with supervision from X.L. and H.Z.; T.C. provided code and performed phenotype querying for the All of Us analysis; X.H. processed and cleaned rare variants in the All of Us analysis; and J.W., X.L., and H.Z. drafted the manuscript, and T.C., X.H., W.W., P.K., and K.Y. provided comments. All co-authors reviewed and approved the final version of the manuscript.

## Methods

We developed RICE, a framework that integrates both common and rare genetic variants to enhance polygenic risk prediction across diverse populations. RICE has three primary components: (1) RICE-CV, which constructs a robust PRS for common variants using ensemble regression; (2) RICE-RV, which identifies and incorporates rare variant signals into the PRS; and (3) a final evaluation step that combines the PRSs from RICE-CV and RICE-RV within a regression model to produce an integrated risk prediction (**Figure 1**).

RICE employs a three-way data split into training, tuning, and validation datasets. The training dataset is used to compute GWAS summary statistics, train common variant PRS models, perform rare variant association testing, compute burden scores, and train rare variant PRS models using penalized regression (LASSO and ridge regression). The tuning dataset is used to train the ensemble learning model, combining PRSs generated by different methods and tuning parameters, using LASSO and ridge regression as base learners. The validation dataset is then used to evaluate the performance of the final PRSs.

### Phenotype and PRS Standardization

To ensure consistency and comparability across ancestries during the tuning and validation stages, we implemented specific standardization procedures for phenotypes and PRSs. For the initial association testing of common variants in GWAS and rare variants in STAARpipeline within the training dataset, we used the original phenotype values. These analyses adjusted for covariates including the top 10 PCs, sex, age, and age squared. Linear regression was applied for continuous traits, and logistic regression was used for binary traits, aligning with standard association testing practices.

During the ensemble learning for RICE-CV and RICE-RV in the tuning dataset, and fitting LASSO and ridge regression models for burden scores in the training dataset (detailed in the later sections), continuous phenotypes were adjusted by regressing out the same set of covariates to obtain residuals. These residuals were used as inputted outcomes in the mentioned procedures. Binary traits remained on their original scale throughout all stages.

In the validation stage, we standardized the residualized continuous phenotypes within each ancestry group to have a mean of 0 and a variance of 1. PRSs generated from RICE-CV and RICE-RV were also standardized within each ancestry to have a mean of 0 and a variance of 1, following methods outlined in prior works^4,15^ and described in the **Supplementary Note**. This standardization ensured that both phenotypes and PRSs were on a common scale across ancestries, facilitating fair comparisons during performance evaluation.

### RICE-CV: Common Variant PRS Modeling

The RICE-CV component has three main steps: (1) PRS training, (2) ensemble learning, and (3) selection of the final PRS.

#### PRS Training

In RICE-CV, common variants are defined as variants with MAF ≥ 0.01 in any of the genetically inferred ancestry groups. Let *û*_*kl*_ denote the estimated effect size of the *k*th genetic variant in the *l*th ancestry group, with *s*_*kl*_ as its standard error. We construct the common variant PRS using several established PRS methods.

For single-ancestry analyses (e.g., in a primarily European population), we employ methods such as CT^11–13^, LDpred2^52^, and Lassosum2^16^, which represent clumping-based, Bayesian, and penalization-based approaches, respectively. For multi-ancestry analyses, we apply CT-SLEB^24^, JointPRS^53^, and PROSPER^27^, covering the same methodological categories. Each method generates multiple PRSs for each individual in the tuning dataset by varying its tuning parameters. Detailed implementations of each method are provided in the **Existing PRS Methods** section.

Let *PRS*_*ij*_, denote the PRS for individual *i* in the tuning dataset, computed using a specific method and tuning parameter set *j*. As detailed in the **Phenotype and PRS Standardization section**, all PRSs are pre-standardized within each ancestry group before they are input into the ensemble model.

#### Ensemble Learning

The ensemble learning step combines the candidate PRSs to optimize predictive performance. Specifically, we use LASSO and ridge regression as base learners within the SuperLearner package^54,80^, which selects the optimal combination of PRSs to maximize prediction accuracy.

For continuous traits, the ensemble model is trained to minimize the cross-validated mean squared error (MSE). The process is as follows: for LASSO regression, we train a model on the tuning dataset using the PRS features as predictors. The model minimizes the objective function:

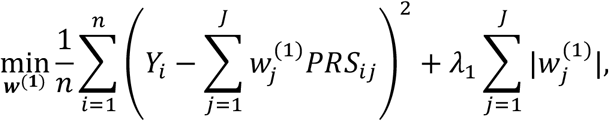

where *Y*_*i*_ is the revisualized phenotypes for indivudal *i* regressing out for covariates, 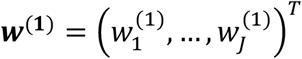 are the weights estimated for each PRS, λ_1_ is the LASSO regularization parameter, and *n* is the number of individuals in the tuning dataset.

For ridge regression, we minimize the objective function:

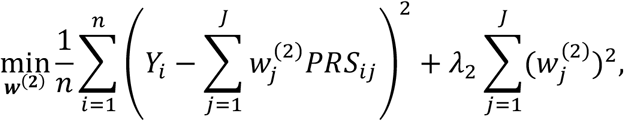

Where 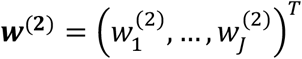 are the weights estimated for each PRS, λ_2_ is the ridge regularization parameter.

The optimal regularization parameters λ_1_ and λ_2_ are selected by minimizing the cross-validated MSE (default fold = 20). With optimized weights for each base learner, we generate predictions for each individual in the tuning dataset as: 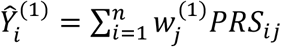, for LASSO and 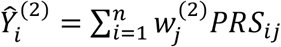. The ensemble learning algorithm then estimates the optimal weights **α** = (α_1_, α_2_)^T^ to combine these predictions from the base learners. This is done by solving:

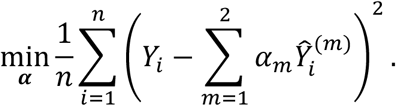

The optimal weights α_1_ and α_2_ are chosen using cross-validation (default fold = 20) to minimize the cross-validated MSE. The final ensemble prediction for individual *i* is a weighted combination of base learner predictions: 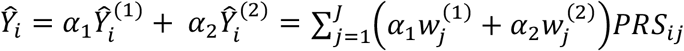.

For binary traits, the ensemble model is trained to minimize the negative log-likelihood function, as ridge regression is not supported for binary outcomes in the SuperLearner package. We use LASSO logistic regression:

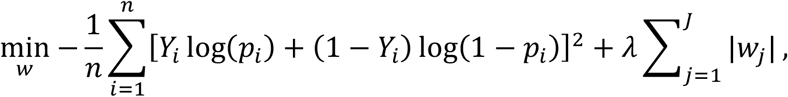

where 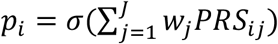 is the predicted probability for individual *i*, with σ(*x*) = *exp*(*x*)/{1 + exp(*x*)} being the logistic function. *Y*_*i*_ is the binary phenotype (0 or 1) for individual *i*. The optimal weight ***w*** by maximizing AUC for binary traits.

#### Selection of the Final PRS

After training, the ensemble prediction is compared to each individual PRS from the input methods on the tuning dataset. For continuous traits, we select the PRS with the lowest MSE; for binary traits, we select based on the highest AUC, capturing the discriminative ability of the PRS. The PRS that achieves the best performance, whether it is the ensemble PRS or an individual PRS, becomes the final common variant PRS for RICE-CV. The final PRS for RICE-CV is denoted as: ***Ŷ***_*CV*_ = ***PRS***_*CV*_ ***w***_*CV*_, where ***Ŷ***_*CV*_ is the predicted response from the common variants. ***PRS***_*CV*_ = [***PRS***_1_, ***PRS***_2_, …, ***PRS***_*j*_] represents the matrix of PRSs generated by the individual methods (with each column ***PRS***_*j*_ = (*PRS*_1*j*_, .., *PRS*_*nj*_)^*T*^ for *n* individuals, and ***w***_*CV*_ is the vector of optimal weights derived from the ensemble learning model. If an individual PRS outperformed the ensemble PRS in the evaluation, ***w***_*CV*_ corresponds to that specific PRS.

### RICE-RV: Rare Variant PRS Modeling

The RICE-RV component focuses on identifying and integrating signals from rare variants into the PRS to enhance genetic risk prediction. It has four main steps: (1) fitting the null model of STAARpipeline^49^ to adjust for covariates, (2) performing rare variant association testing by functional grouping, (3) modeling significant rare variant sets using penalized regression, and (4) combining PRSs through ensemble learning. Steps 1-2 are based on STAARpipeline focusing on association testing. Steps 3-4 focuses on building the PRS model using the select rare variants sets.

#### Fitting the Null Model

We first fit a null model using STAARpipeline to adjust for covariates and the predicted common variant PRS from RICE-CV. This step ensures that rare variant signals are independent of common variant effects. For each individual *i*, let *Y*_*i*_ denote the phenotype of interest, which can be continuous (e.g. height) or binary (e.g., disease status). We model the conditional mean of *Y*_*i*_ using:

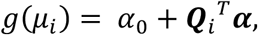

where *g*(*μ*_*i*_) is the link function (identify for continuous traits, logit for binary traits). α_0_ is the intercept term, ***Q***_*i*_ = (*Q*_*i*1_, …, *Q*_*iq*_)^*T*^ represent *q* covariates, such as age, sex, PCs and the common variant PRS from RICE-CV (Ŷ_*CV,i*_).

#### Rare Variant Association Testing

After fitting the null model, we perform rare variant association testing using STAARpipeline, which is specifically designed for large-scale sequencing studies. STAARpipeline adjusts for population structure, corrects for case-control imbalances, and incorporates functional annotations to increase the power of rare variant tests.

Rare variants are grouped into biologically meaningful sets based on functional categories, increasing the power to detect associations by aggregating variants likely to affect gene function. To capture orthogonal signals, RICE-RV uses STAARpipeline to perform association tests on rare variant sets, conditional on the RICE-CV PRS. STAARpipeline defines gene-centric rare variant sets by functional categories. For variants in coding regions, it analyzes five categories per gene: (1) putative loss of function (stop gain, stop loss and splice); (2) missense, (3) disruptive missense, (4) putative loss of function and disruptive missense; and (5) synonymous. For variants in noncoding regions, it tests eight categories: (1) promoter overlaid with CAGE sites, (2) promoter overlaid with DHS sites, (3) enhancer overlaid with CAGE sites, (4) enhancer overlaid with DHS sites, (5) UTR, (6) upstream region, (7) downstream region, and (8) ncRNA. In our study, we use variants in coding regions for WES data and variants in coding and noncoding regions for WGS data.

Within each set, STAARpipeline performs annotation-weighted tests, aggregating information across variants and weighting each variant based on its functional annotation. Multiple test statistics, including burden, SKAT, and ACAT-V, are computed, and an omnibus p-value is generated using ACAT to combine correlated p-values. Within RICE-RV, we focus on the p-values derived from the annotation-weighted burden tests, referred to as STAAR-B (1,1) p-values in STAARpipeline. We identify significant rare variant sets as those with STAAR-B (1,1) p-value less than *p* < 1 × 10^−3^.

#### Modeling Significant Rare Variant Sets Using Penalization Regression

For each significant rare variant set, we compute a burden score for each individual to summarize the collective effect of the rare variants within that set. Let *K* denote the total number of significant rare variant sets identified. For the *k*-th rare variant (*k* = 1, 2, …, *K*), let *m*_)_ be the number of rare variants in the *k*th set. For individual *i*, let 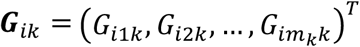 denote the vector of genotypes for the rare variants within the *k*-th set. Here, *G*_*ijk*_ represents the genotype of individual *i* for the *j*-th rare variant in set *k*, coded as the number of minor alleles (e.g., 0, 1, or 2). The burden score for individual *i* for the *k*th rare variant set is defined as:

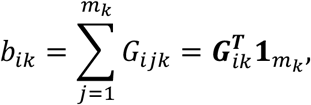

where 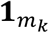 is a vector of ones of length *m*_*k*_. This burden score *b*_*ik*_ represents the total number of minor alleles carried by individual *i* across all rare variants in the *k*-th rare variant set.

We then model the relationship between phenotype *Y*_+_ and burden scores using penalized regression methods, specifically LASSO and ridge regression. For continuous traits, we estimate the coefficients ***γ*** = (*γ*_1_, *γ*_2_, …, *γ*_*k*_)^*T*^ by minimizing the following objective function:

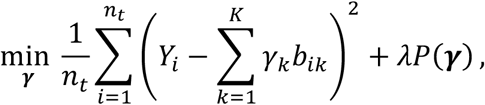

where *Y*_*i*_ is the revisualized phenotypes regressing out for covariates, *n*_*t*_ is the total of number individuals in the training dataset, and λ is the regularization parameter controlling the penalty strength. *P*(***γ***) is the penalty function (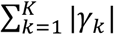 for LASSO or 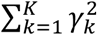 for ridge regression).

For binary traits, we use a logistic regression framework with penalization, estimating ***γ*** by minimizing:

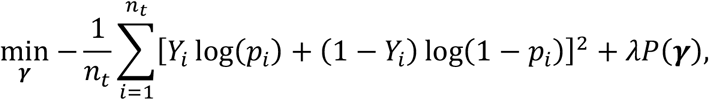

where 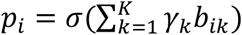 is the predicted probability for individual *i*, with σ(*x*) = *exp*(*x*)/{1 + exp(*x*)} being the logistic function. For fitting the penalized regression models on burden scores, we used the glmnet^81^ package in R (version 4.1.8). The PRS for specific individual *i* given the estimated 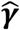 under a specific penalization λ, can be calculated as 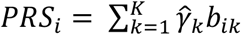.

#### Ensemble learning to combine PRSs

After computing the burden score PRSs, we follow a similar ensemble learning step on the tuning dataset as in RICE-CV to compute the final PRS for RICE-RV. Using the SuperLearner package, we combine predictions from different penalized regression models, optimizing predictive performance. The final PRS for RICE-RV is denoted as: Ŷ = ***PRS***_*RV*_***w***_*RV*_ where Ŷ_*RV*_ is the predicted response from the rare variants. ***PRS***_*RV*_ = [***PRS***_1_, ***PRS***_2_, …, ***PRS***_*j*_] represents the matrix of PRSs generated by based on the burden score, with each column ***PRS***_*j*_ = (*PRS*_1*j*_, .., *PRS*_*nj*_)^*T*^ corresponds to a PRS derived from a specific penalized regression model for *n* individuals. ***w***_*RV*_ is the vector of optimal weights derived from the ensemble learning model. If an individual PRS outperformed the ensemble PRS in the evaluation, ***w***_*RV*_ corresponds to that specific PRS.

### RICE Validation

The final RICE prediction combines the PRSs generated from RICE-CV and RICE-RV using a regression model, with weights estimated from the validation dataset. For continuous traits, the standardized residuals from the phenotype regression are used as the outcome variable in a linear model: 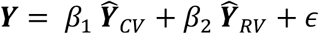, where ***Ŷ***_*CV*_ and ***Ŷ***_*RV*_ are the standardized PRSs from RICE-CV and RICE-RV, respectively, and β_1_ and β_2_ are their estimated weights. For binary trait, we use logistic regression: 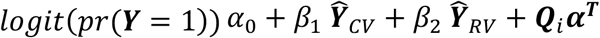, where ***Q***_*i*_ represent other covariates, such as age, sex, PCs.

### Genetically-Inferred Ancestry Labels

For the analysis of UKB WES and WGS data, genetically inferred ancestry groups were defined using the 1000 Genomes Project Phase 3 (1000G) dataset^55^, which includes 3,202 individuals across five ancestry groups: 893 AFR, 490 AMR, 585 EAS, 633 EUR, and 601 SAS. We first performed principal component analysis on all 3,202 samples using plink v2.0^82^, using a set of selected variants provided from gnomAD to capture population structure. Using the first five PCs, we trained a random forest classifier to accurately assign each individual in the 1000G dataset to their respective ancestry group. Next, we projected the UKB WES and WGS datasets onto this PC space and applied the pre-trained random forest classifier to assign genetically-inferred ancestry groups to each UKB sample. For the analyses of AoU data, genetically inferred ancestry groups (EUR, EAS, SAS, AMR, MID, AFR) were directly provided by the All of Us Research Program^42^.

### Simulation Study

We conducted large-scale simulations using simulated traits generated from chromosome 22 of the UKB WES dataset. The training dataset included only individuals with EUR ancestry, with sample sizes of 49,173 or 98,343. The remaining individuals, comprising those with EUR, AFR, AMR, EUR and SAS ancestry, were evenly divided into tuning and validation sets.

Causal variants were randomly selected and included both common variants (MAF > 0.01 in the UKB WES dataset) and rare variant sets. The proportion of causality varied among three levels: 0.01, 0.05, and 0.2. For each level, we randomly selected causal common variants and causal rare variant burdens with these proportions. The heritability was kept constant across simulations, with 0.05 assigned to causal common variants and 0.0125 to causal rare variant burdens. Additionally, we considered two scenarios of negative selection: strong negative selection and no negative selection.

Let *u*_*CV,q*_ denote the standardized effect size of the *q*th causal common variant, and *u*_*RV,k*_ denote the standardized effect size for the *k*th causal rare variant burden. Under strong negative selection scenario, the standardized effect sizes were drawn from normal distributions: 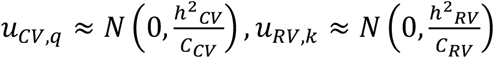, where *C*_*CV*_ is the number of causal common variants, *h*^2^_*CV*_ = 0.05 is the heritability attributed to the common variants, *C*_*RV*_ is the number of causal rare variant sets, and *h*^2^_*RV*_ = 0.0125 is the heritability attributed to the rare variant sets.

Phenotypes were simulated using the following linear model:

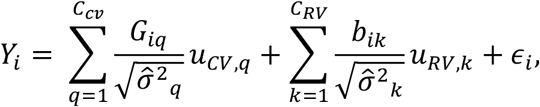

where *Y*_*i*_ is the phenotype for individual *i, G*_*iq*_ is the genotype of individual *i* at the *q*th causal common variant, *b*_*ik*_ is the burden score of individual *i* for the *k*th causal rare variant set, and *ϵ*_*i*_ is the residual error term. The variance 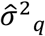 and 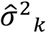 are defined as follows: 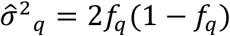, where *f*_*q*_ is the effect allele frequency for the *q*th causal common variant, and 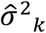 is calculated similarly for the *k*th rare variant sets. Under the no negative selection scenario, we set 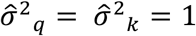. Each simulation scenario had 100 simulated traits, and the final presented results is the average of the 100 validation dataset results.

### Existing PRS Methods

CT^11–13^ is a method that first removes variants in high LD with an index variant that has the lowest p-value within a specified base-pair window. From the remaining set of index SNPs, multiple p-value thresholds are applied to generate PRSs. The optimal PRS and corresponding p-value threshold are then chosen based on performance in the tuning dataset. We implemented CT using plink1.9^82^ with an LD threshold *r*^2^ of 0.5 (--clump-r2), a window size of 500 kb, and nine p-value thresholds ranging from 5 × 10^−8^ to 1.

Lassosum2^16^ implements LASSO regression using GWAS summary statistics and LD information. Lassosum2 penalizes effect sizes according to LD by minimizing the objective function: ***β***^*T*^((1 − *s*)***R*** + *s****I***)***β*** – ***β***^*T*^ ***r*** + λ_1_‖***β***‖_1_, where ***β*** is the vector of effect sizes, ***R*** is the LD matrix, ***I*** is the identify matrix, ***r*** is the vector of marginal correlations from summary statistics, and *s* is a tuning parameter to ensure convexity. The parameter λ_1_ controls *L*_1_ penalty, with‖***β***‖_1_ denote the *L*_1_ norm of ***β***. We implemented Lassosum2 with 10 different values of *s* (ranging from 0.5 to 1000) and 30 values of λ_1_.

LDpred2^52^ utilizes a Bayesian framework with a spike-and-slab prior for variant effects β. The prior density is given by 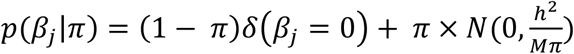, where *δ* is the Dirac delta function, *π* is the proportion of casual variants, *h*^2^ is the total heritability, *M* is the total number of variants, and *π* is the proportion of nonzero variants. Posterior effect sizes are estimated using Markov Chain Monte Carlo (MCMC) sampling. We conducted a grid search over 15 values of *h*^2^ (from 0.1 to 1.5) times the heritability estimated by LD score regression, 17 values of *π* ranging from 1 × 10^−4^ to 1, and set the sparse setting to “False”. Both LDpred2 and Lassosum2 were implemented in R using the bigsnpr package (version 1.12.6).

CT-SLEB^24^ is a recent multi-ancestry extension of CT method that consists of three steps: two-dimensional CT, calibration of regression coefficients using Empirical Bayes, and ensemble learning using SuperLearner. Two dimensional CT and Empirical Bayes are implemented between pairs of ancestries, designating one as the target population and the other as the reference population. For our analysis of the AoU data, which included three ancestries in the training dataset (AFR, AMR, and EUR), CT-SLEB was implemented twice, treating AFR and AMR as target populations with EUR as the reference population. We further implemented the SuperLearner step to include both sets of PRSs, producing a single prediction for all ancestries. The hyperparameters required for CT-SLEB are identical to those used in CT. We implemented CT-SLEB with eight different p-value thresholds ranging from 5 × 10^−8^ to 1, two window sizes (50kb and 100 kb), and six LD threshold (*r*^2^ = 0.01, 0.05, 0.1, 0.2, 0.5, 0.8).

JointPRS^53^ is a multi-ancestry PRS method that extends the PRS-CSx^26^ method. JointPRS assumes variant effect size ***β***, across multiple populations using a correlated Gaussian prior: ***β***_*j*_ ∼ *N*(0, *ψ*_*j*_***MΣM***), where ***MΣM*** accounts for correlations across ancestries and differences in the sample size, and *ψ*_*j*_ is a variant-specific hyper-parameter with a hyper-prior *ψ*_*j*_ ∼ *Gamma*(1, *δ*_*j*_) and 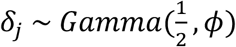. We implemented JointPRS Auto version, which assumes *ϕ* ∼ *Cauchy*(0,1). JointPRS Auto requires only GWAS summary statistics from the training data and does not need tuning data for parameter estimation. LD information was provided using the 1000G reference panel included with the PRS-CSx^26^ software.

PROSPER^27^ is an ensemble penalized regression multi-ancestry PRS method. It involves three steps: (1) performing single-ancestry Lassosum2 to estimate optimal parameters, second, (2) constructing a joint analysis across ancestries using penalized regression, and (3) combining all PRSs from the second step using ensemble learning with SuperLearner. We implemented single ancestry Lassosum2 with the default five values for *s* and five values for λ_1_, as described in the Lassosum2 section. The joint multi-ancestry analysis was performed similarly, using default values for the penalty parameters for λ and *c* following PROSPER GitHub guidance. Ensemble regression was performed using SuperLearner.

### UKB WES and WGS Analysis

We analyzed the UKB WES (Field #24304) and WGS (Field #24305) datasets, focusing on participants from four ancestries: AFR, AMR, EUR, and SAS. EAS ancestry was excluded due to limited sample size in the UKB data, which led to unstable performance in the evaluation. Our analyses included six continuous traits: BMI, height, HDL, LDL, log(TG), and TC, and five binary traits: asthma, breast cancer, CAD, prostate cancer, and T2D.

Participants were randomly divided into training, tuning, and validation sets using stratified sampling within each ancestry, allocating 70%, 15%, and 15% of the total sample size, respectively. Since EUR is the majority ancestry in UKB, we restricted the training dataset to EUR participants, while both EUR and non-EUR participants were included in the tuning and validation sets. Detailed sample sizes for each group are provided in **Supplementary Table 1**. Standard quality control procedures were applied to both common and rare variants (**Supplementary Note**). All analyses, including GWAS, rare variant association testing, and ensemble learning within RICE-CV and RICE-RV, were adjusted for control covariates: the first 10 PCs, sex, age, and age squared.

We evaluated PRS prediction performance on the validation set across five methods: CT, LDpred2, Lassosum2, RICE-CV, and RICE-RV. Both phenotypes and PRSs were standardized to have mean 0 and variance 1 within each ancestry, as described in the **Phenotype and PRS Standardization** section. Performance of the standardized PRSs was assessed by fitting a linear regression model between the standardized phenotype and standardized PRS. For binary traits, PRS standardization followed the same procedure as for continuous traits, and prediction performance was evaluated using logistic regression with control covariates.

For the CT, LDpred2, and Lassosum2 methods, LD reference panels were estimated from 3,000 individuals randomly sampled from the training dataset for both WES and WGS analyses. In the WGS analysis, due to computational constraints, LDpred2 and Lassosum2 analyses were restricted to a subset of variants included in the HapMap3 (HM3) array. The STAARpipeline was implemented differently for the WES and WGS analyses. In the WES analysis, we used rare variants located in the coding region of protein-coding genes. For the WGS analysis, we included rare variants located in both coding and noncoding regions of protein-coding genes.

### AoU Analysis

We analyzed the AoU dataset using WGS data version 7.1. For common variants, we used the [Allele Count/Allele Frequency (ACAF) threshold data](https://support.researchallofus.org/hc/en-us/articles/14929793660948-Smaller-Callsets-for-Analyzing-Short-Read-WGS-SNP-Indel-Data-with-Hail-MT-VCF-and-PLINK) provided in the AoU Researcher Workbench, which include variants with MAF > 0.01 or allele count > 100 in any of the computed ancestry groups. For rare variants, we focused on exonic regions using the [“Exome” dataset](https://support.researchallofus.org/hc/en-us/articles/14929793660948-Smaller-Callsets-for-Analyzing-Short-Read-WGS-SNP-Indel-Data-with-Hail-MT-VCF-and-PLINK) from the AoU Researcher Workbench. We employed the genetically inferred ancestry groups provided by AoU, based on variants from gnomAD^67^, the Human Genome Diversity Project^83^, and 1000G^55^. The ancestries included in our analysis were AFR, AMR, EAS, MID, EUR, and SAS. We examined six continuous traits: BMI, height, HDL, LDL, log(TG), and TC.

Data were randomly split using stratified sampling within ancestries into training, tuning, and validation, comprising 70%, 15%, and 15% of total sample size, respectively. Since AFR, AMR, and EUR are the majority ancestries in the AoU database, we included only these three ancestries in the training dataset. For the tuning and validation datasets, we included all available ancestries in AoU. The detailed sample size breakdown is provided in **Supplementary Table 3**. Both common and rare variants underwent standard quality control procedures (**Supplementary Note**). All analyses, including GWAS, rare variant association testing, and ensemble learning within RICE-CV and RICE-RV, were adjusted for control covariates: the first 10 PCs, sex, age, and age squared.

We compared PRS prediction performance on the validation set across five methods: CT-SLEB, JointPRS, PROSPER, RICE-CV, and RICE-RV. Following a similar procedure as in the UKB analyses, we standardized the phenotypes and PRSs to have a mean of 0 and variance of 1 within each ancestry. The detailed standardization procedure is provided in the **Phenotype and PRS Standardization** section. The performance of the standardized PRSs was assessed by fitting a linear regression model between the standardized phenotype and the standardized PRS.

LD reference data for JointPRS and PROSPER was publicly provided and built using the 1000G reference data. LD reference data for CT-SLEB was derived from a random subset of 3,000 individuals from the training dataset. The provided LD for JointPRS and PROSPER were constrained to HM3 or HM3 + Multi-Ethnic Genotyping Array. Therefore, these methods were analyzed on this subset of SNPs. CT-SLEB was implemented on the full set of SNPs. RICE-RV was implemented using rare variants located in coding regions of protein-coding genes utilizing STAARpipeline.

## Data Availability

UK Biobank phenotype data, WES data, and WGS data can be accessed through the UK Biobank research analysis platform (https://ukbiobank.dnanexus.com/landing). All data used in this research are publicly available to registered researchers through the UKB data-access protocol and who are listed as collaborators on UKB-approved access applications. All of Us phenotype data, WES data, and WGS data can be accessed through the All of Us research workbench (https://workbench.researchallofus.org/). All data used in this research are publicly available to registered researchers with controlled tier access through the All of Us data-access protocol.

## Code Availability

Simulation and data analyses code is available at: GitHub (https://github.com/jwilliams10/RareVariantPRS)

Tutorial to implement RICE is available at: GitHub (https://github.com/jwilliams10/RICE) Software implementing CT-SLEB is available at: GitHub (https://github.com/andrewhaoyu/CTSLEB)

Software implementing PROSPER is available at: GitHub (https://github.com/Jingning-Zhang/PROSPER)

Software implementing JointPRS is available at: GitHub (https://github.com/LeqiXu/JointPRS)

Lassosum2 and LDpred2: https://github.com/privefl/bigsnpr

PLINK: https://www.cog-genomics.org/plink/1.9/

PLINK2: https://www.cog-genomics.org/plink/2.0/

The majority of our statistical analyses are performed using the following R packages: ggplot2 version 3.5.1, dplyr version 1.1.4, bigsnpr version 1.12.15, SuperLearner version 2.0.29, caret version 6.0.94, glmnet version 4.1.8, STAARpipeline version 0.9.7, TxDb.Hsapiens.UCSC.hg38.knownGene 3.18.0, SeqVarTools 1.42.0, SeqArray 1.44.0

