## Supplementary Figures and Notes for "Integrating Common and Rare Variants Improves Polygenic Risk Prediction Across Diverse Populations"

### Supplementary Figures and Supplementary Note

**Supplementary Figure 1. Simulation results comparing the predictive performance of PRSs for four ancestral groups from the UK Biobank (UKB).** The training data had a sample size of N = 49,173 (Supp. Fig. 1a and 1c) or N = 98,343 (Supp. Fig. 1b) with only individuals of European ancestry (EUR), while the tuning (N = 20,869) and validation datasets (N = 20,868) contained individuals of African (AFR), Admixed American or Latino (AMR), European (EUR), and South Asian (SAS) ancestries (**Supplementary Table 1**). Simulations assumed a common variant heritability of 0.05 and a rare variant set heritability of 0.0125, under the assumption of either strong negative selection (Supp. Fig. 1a and 1b) or no negative selection effect size distribution (Supp. Fig. 1c) (**Methods**). Causal proportions for both common variants and rare variant sets varied across three levels: 0.01 (top), 0.05 (middle), and 0.2 (bottom). Data were generated using unrelated individuals from UK Biobank whole-exome sequencing data (WES), with simulation based on chromosome 22. PRS performance is reported as the “Beta of PRS per standard deviation (SD)”, derived from the regression model $Y \sim PRS \times \beta$, with $\beta$ representing the effect of standardized PRS on the standardized outcome (**Methods**). For RICE, the model used was $Y \sim PRS_{CV} \times \beta_{CV} + PRS_{RV} \times \beta_{RV}$. Beta values can be interpreted as the square root of the heritability ($\sqrt{h^2}$) of the outcome explained by the PRS (**Supplementary Note**).

**a)** Training dataset consisting of 49,173 individuals of European Ancestry and data simulated under a strong negative selection model.

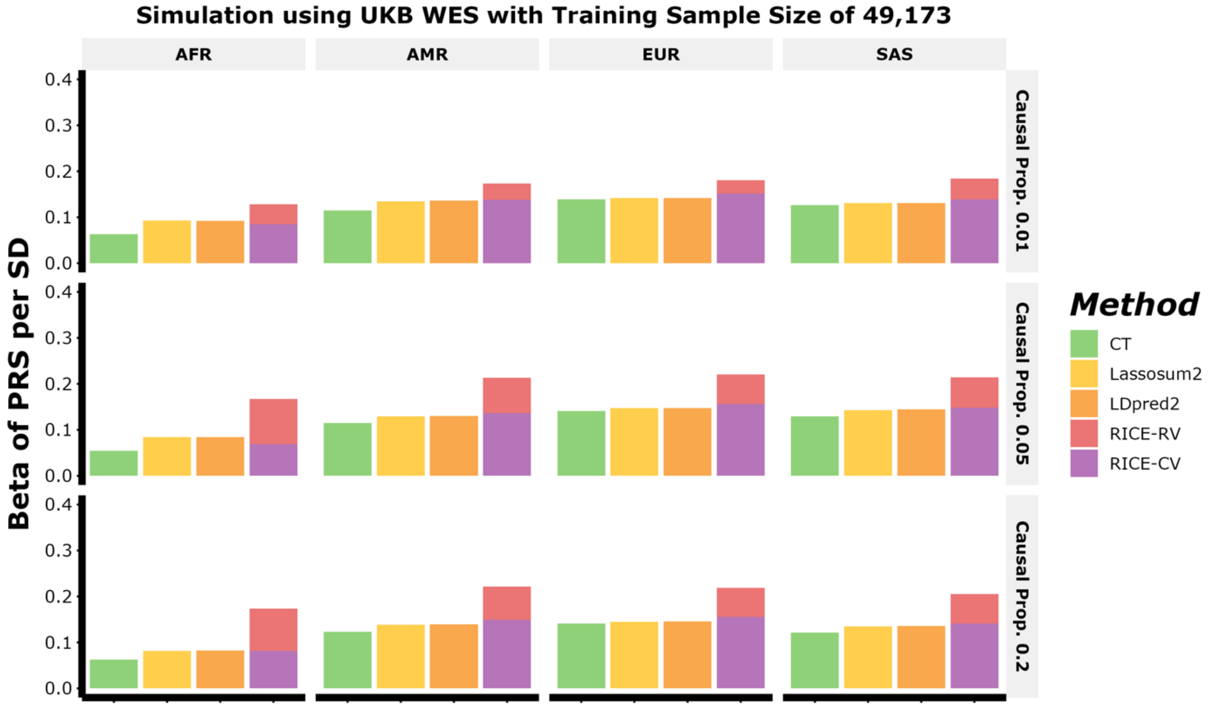

**Supplementary Figure 1 continued**

***b)*** *Training dataset consisting of 98,343 individuals of European Ancestry and data simulated under a strong negative selection model.*

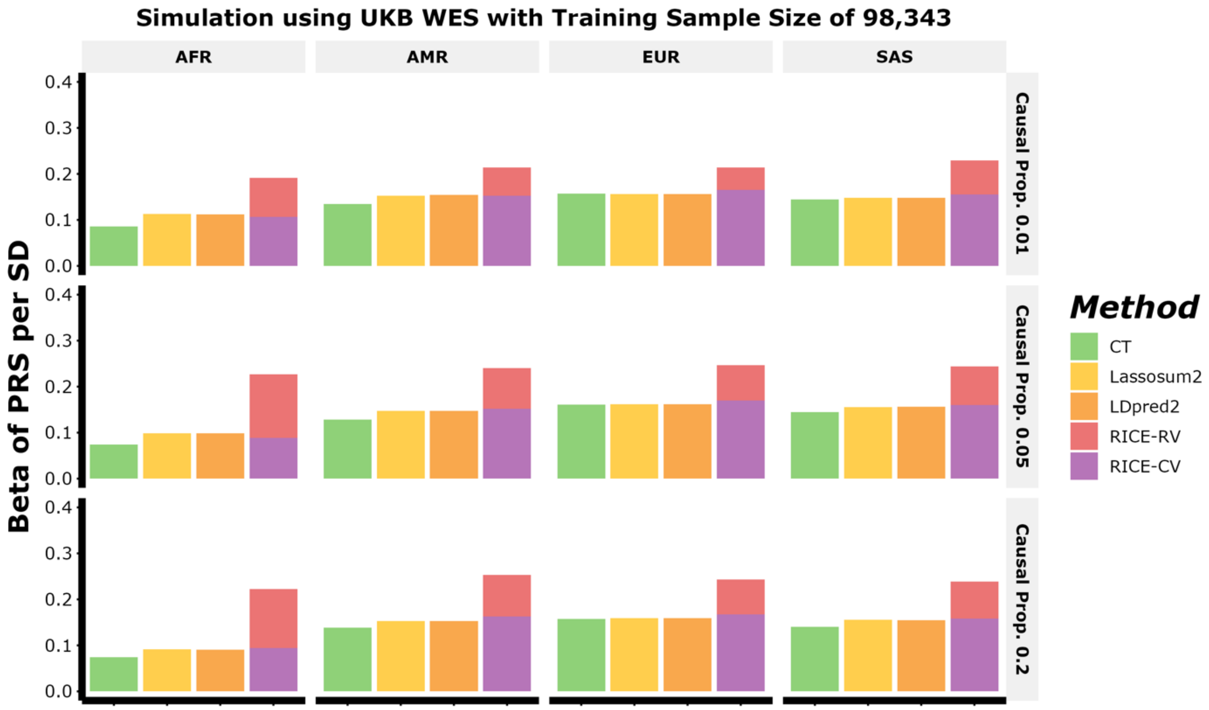

**Supplementary Figure 1 continued**

**c)** Training dataset consisting of 49,173 individuals of European Ancestry and data simulated under no negative selection.

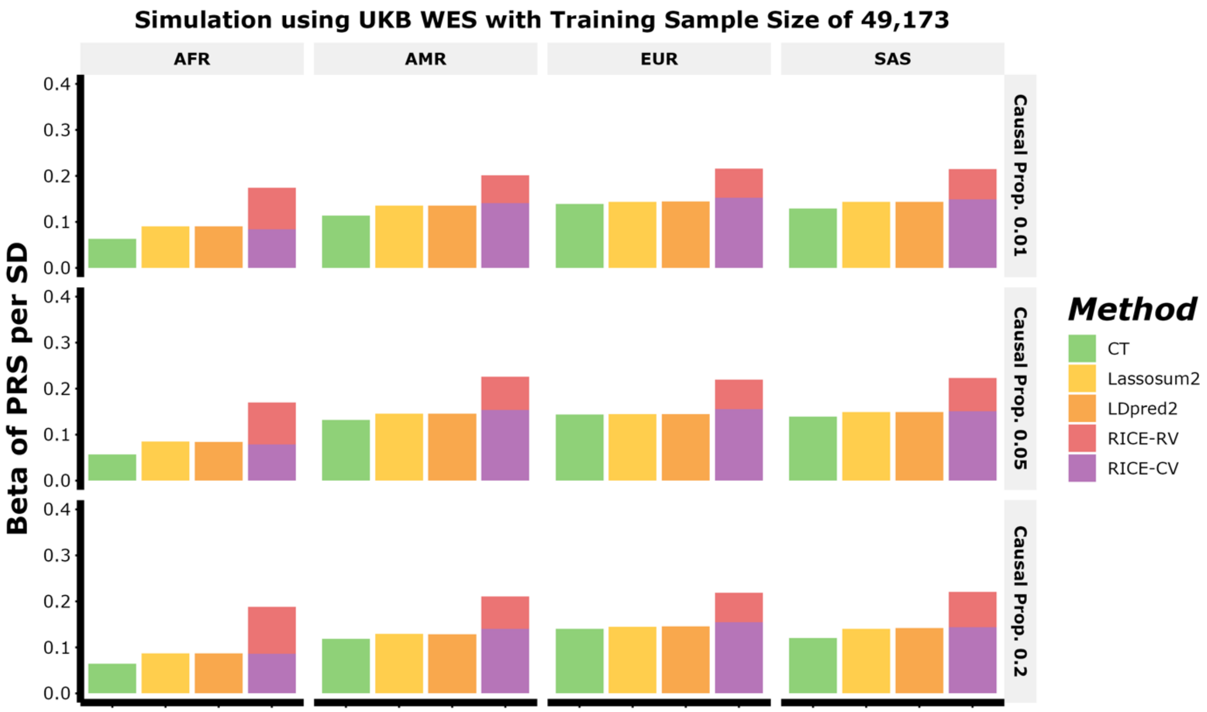

**Supplementary Figure 2. Average heritability of rare variant burden scores for each ancestry by simulation design.** Simulations assumed a common variant heritability of 0.05 and a rare variant set heritability of 0.0125. Causal proportions for both common variants and rare variant sets varied across three levels: 0.01 (top), 0.05 (middle), and 0.2 (bottom). Data was simulated under both strong negative selection (Scaled: Yes) and no negative selection (Scaled: No). Data were generated using unrelated individuals from UK Biobank whole-exome sequencing data (WES), with simulation based on chromosome 22.

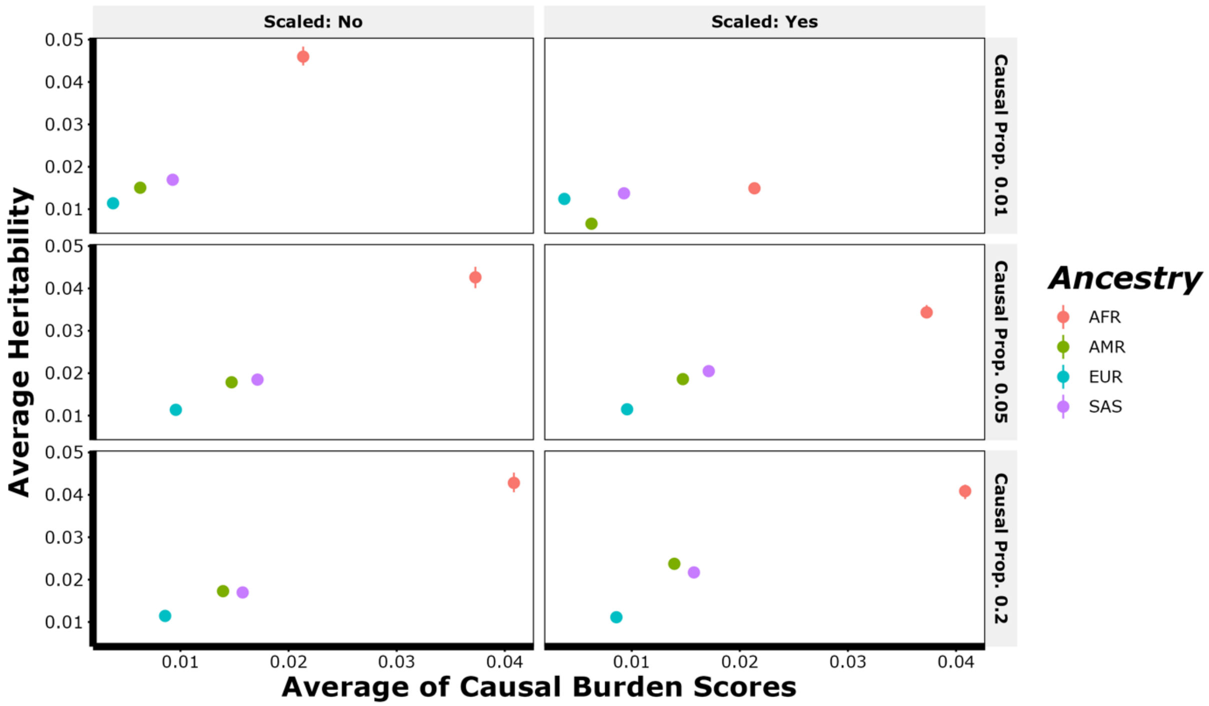

**Supplementary Figure 3. Predictive performance of PRSs for five binary traits across four ancestral groups from UK Biobank (UKB) whole-exome sequencing (WES) data.** The binary traits analyzed include asthma, breast cancer, coronary artery disease (CAD), prostate cancer and type 2 diabetes (T2D). Results are shown for individuals of African (AFR), Admixed American or Latino (AMR), European (EUR), and South Asian (SAS) ancestries. The training data consisted solely of individuals of European ancestry, while tuning and validation sets included all four ancestries. Full sample sizes details for each ancestry are provided in **Supplementary Table 2**.

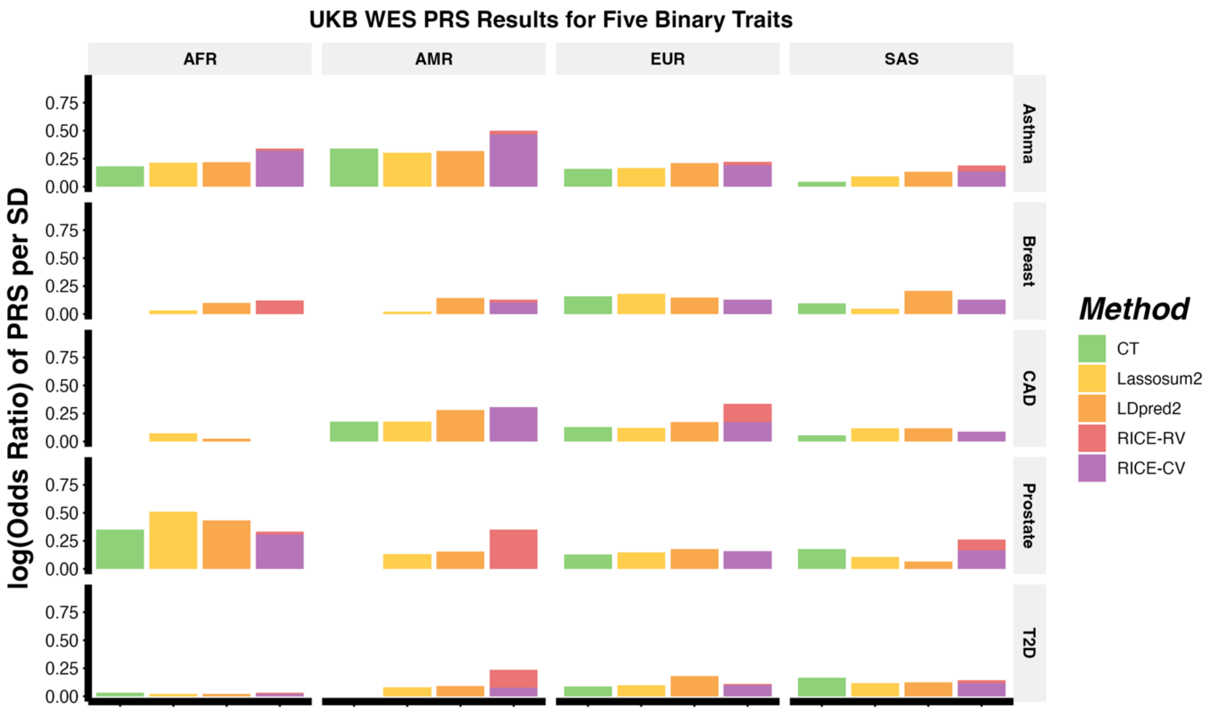

**Supplementary Figure 4. Predictive performance of PRSs standardized within genetically-inferred ancestries or using the first five principal components (Methods) for six continuous traits and five binary traits across four ancestral groups from UK Biobank (UKB) whole-exome sequencing (WES) data.** The continuous traits analyzed include body mass index (BMI), high-density lipoprotein cholesterol (HDL), height, low-density lipoprotein cholesterol (LDL), natural logarithm of triglyceride cholesterol (log(TG)), and total cholesterol (TC). The binary traits analyzed include asthma, breast cancer, coronary artery disease (CAD), prostate cancer and type 2 diabetes (T2D). Results are shown for individuals of African (AFR), Admixed American or Latino (AMR), European (EUR), and South Asian (SAS) ancestries. Note that for BMI, the estimated Beta of PRS per SD for African and Admixed American have much larger standard errors than for European and South Asian. This is because there are a few individuals of European or South Asian ancestry that had a predicted PRS significantly higher than the average individual. This affected the standardization using regression with the first five principal components as these values were influential outliers adjusting the standardized PRSs distribution.

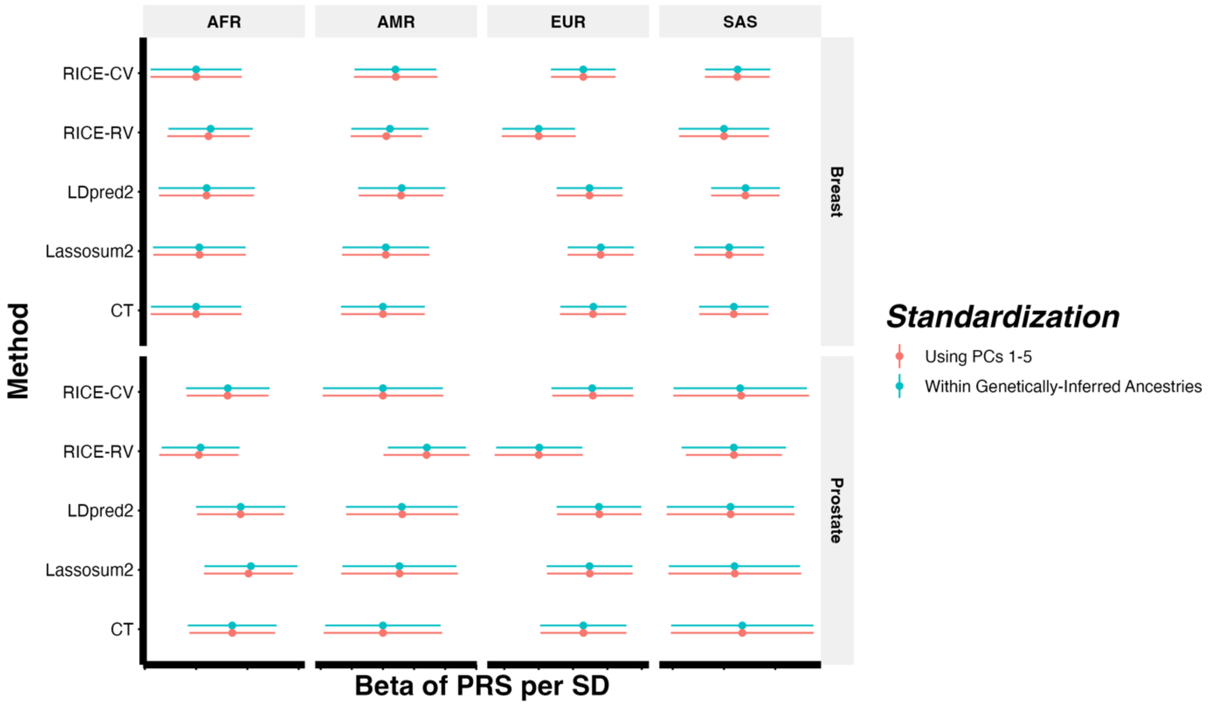

**Supplementary Figure 4 continued.** Figures for CAD, T2D and asthma are included in this page.

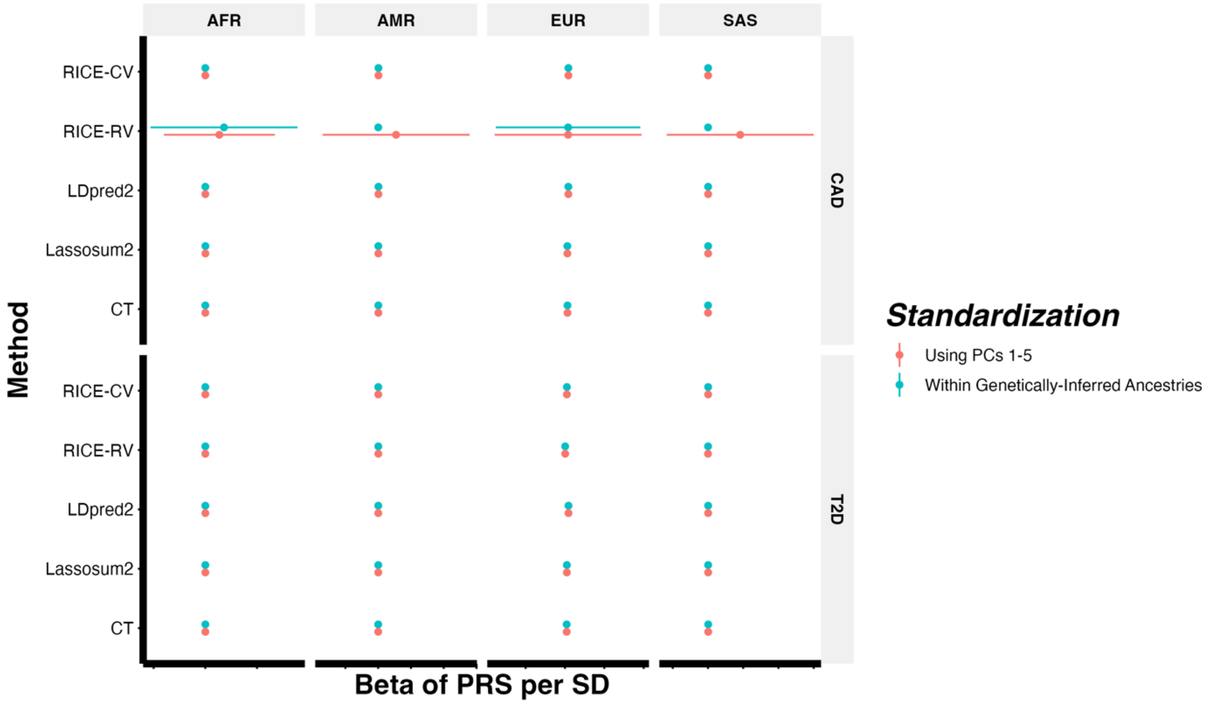


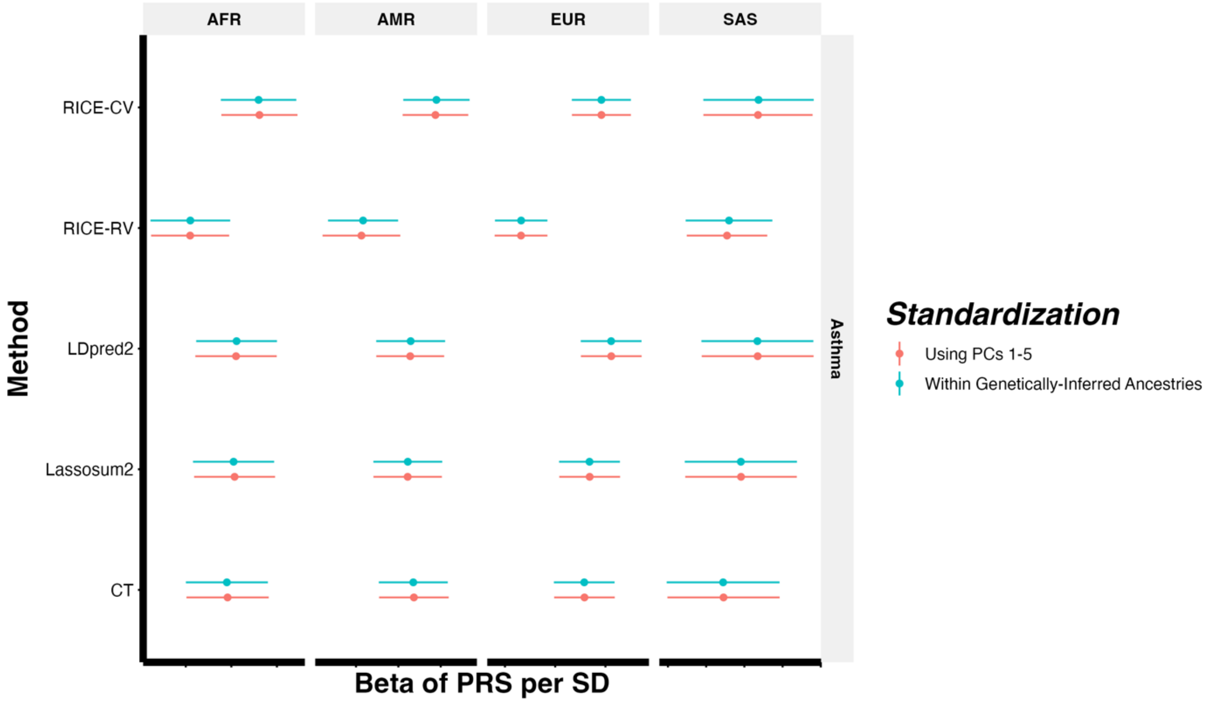

**Supplementary Figure 4 continued.** Figures for BMI, height, HDL and LDL are included in this page.

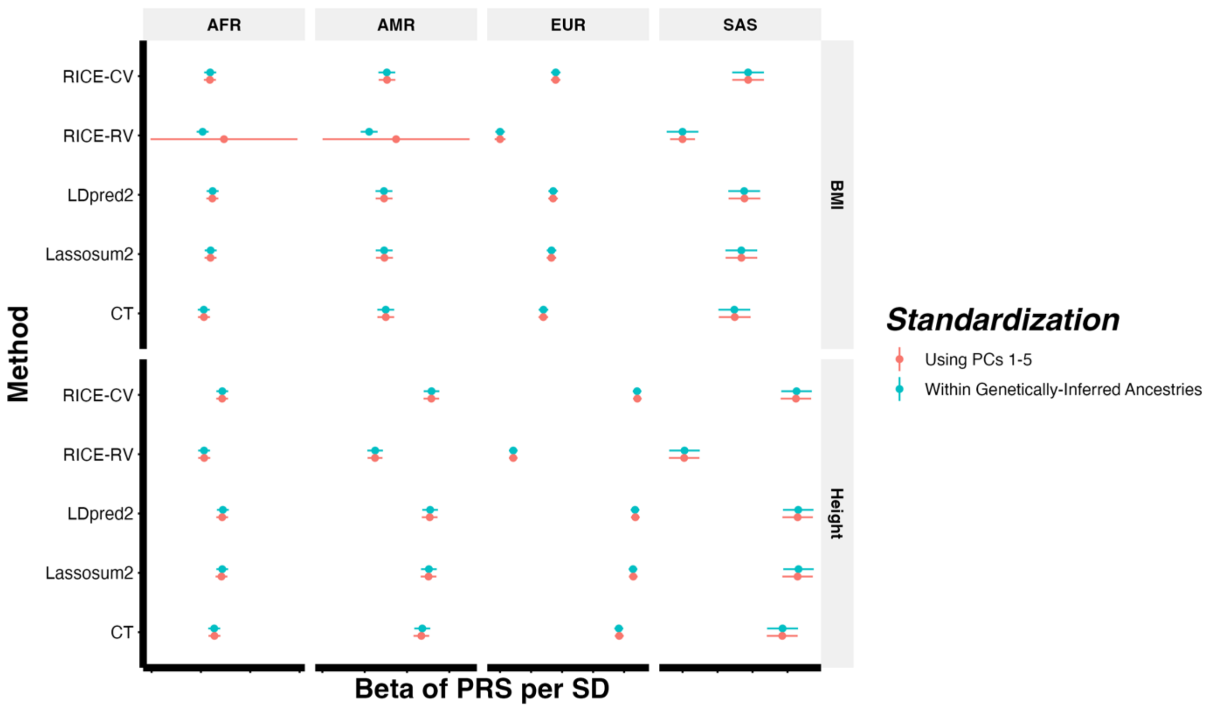


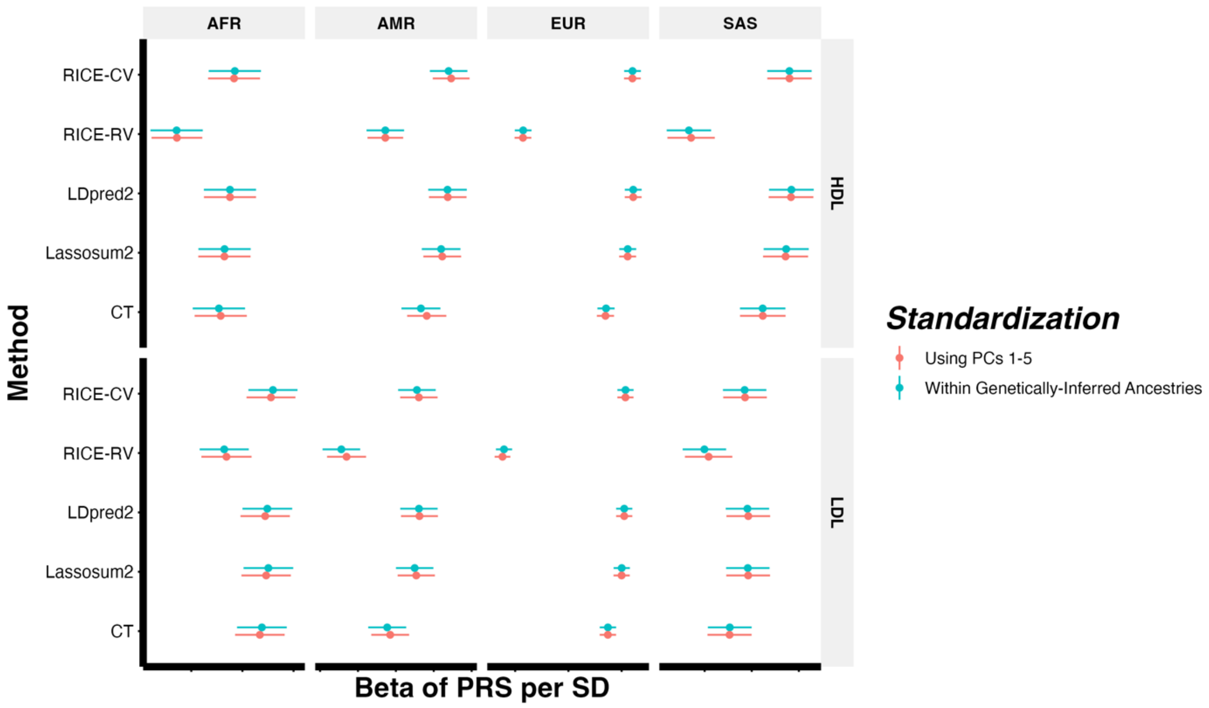

**Supplementary Figure 4 continued.** Figures for log(TG) and TC are included in this page.

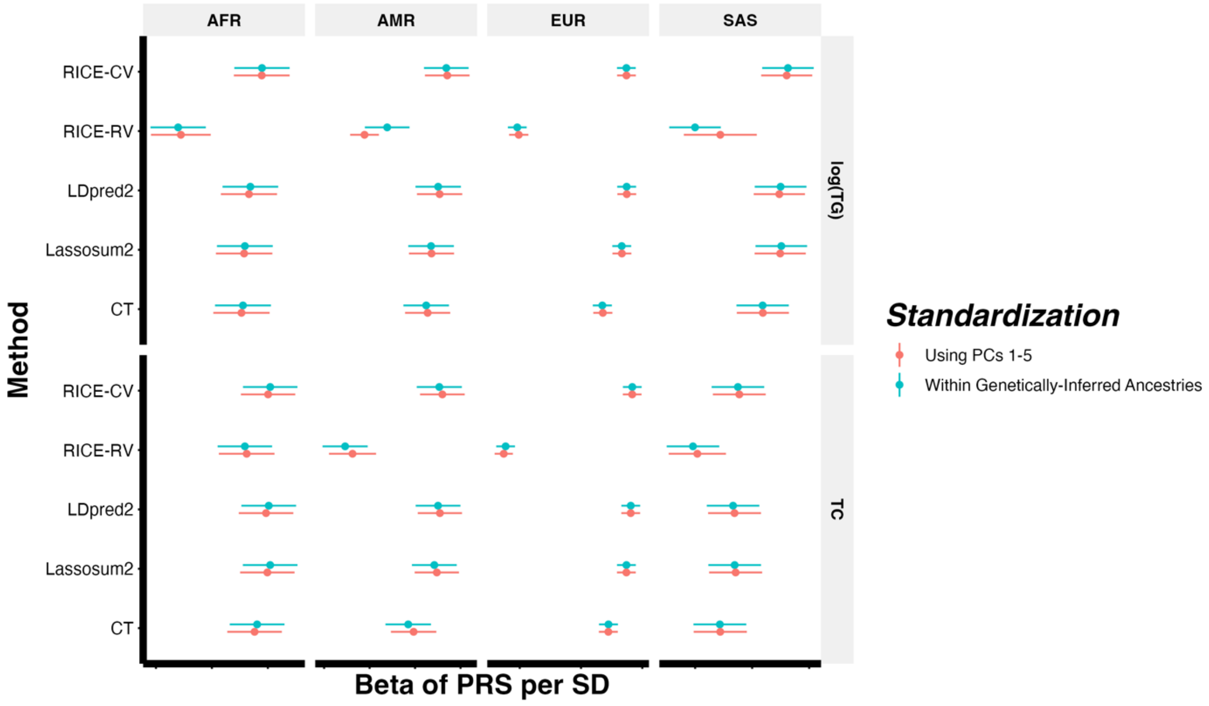

**Supplementary Figure 5. Relationship between common and rare variant PRSs and the estimated odds ratio for Europeans from UK Biobank (UKB) whole-exome sequencing (WES) data.** Figures are shown for the five binary traits: asthma (Supp. Fig. 5a), breast cancer (Supp. Fig. 5b), coronary artery disease (CAD) (Supp. Fig. 5c), prostate cancer (Supp. Fig. 5d), and type 2 diabetes (Supp. Fig. 5e). PRS quantiles for RICE-CV (common variants) are plotted on the x-axis, and estimated odds ratio is on the y-axis. Data are stratified by rare variant PRS quantiles from RICE-RV (red: below 5%, green: 20–70%, blue: above 95%). Results are shown for individuals of European (EUR) ancestries. The training data consisted solely of individuals of European ancestry, while the tuning and validation sets included all four ancestries. Full sample sizes details for each ancestry are provided in **Supplementary Table 2**.

**a)** Relationship between ancestry-adjusted common and rare variant PRSs and the estimated odds ratio of asthma for Europeans from UKB WES data.

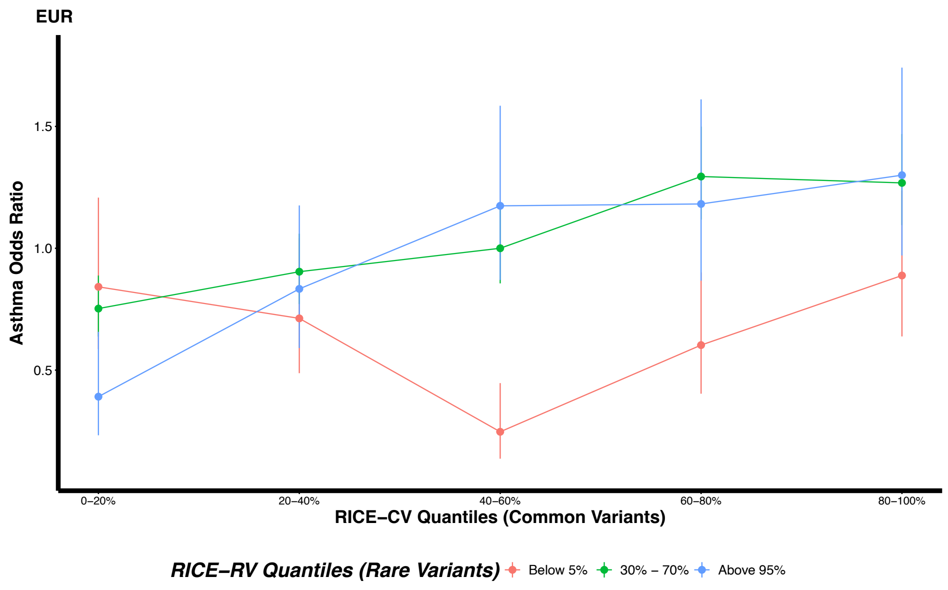

**Supplementary Figure 5 continued. b)** Relationship between ancestry-adjusted common and rare variant PRSs and the estimated odds ratio of breast cancer for Europeans from UKB WES data.

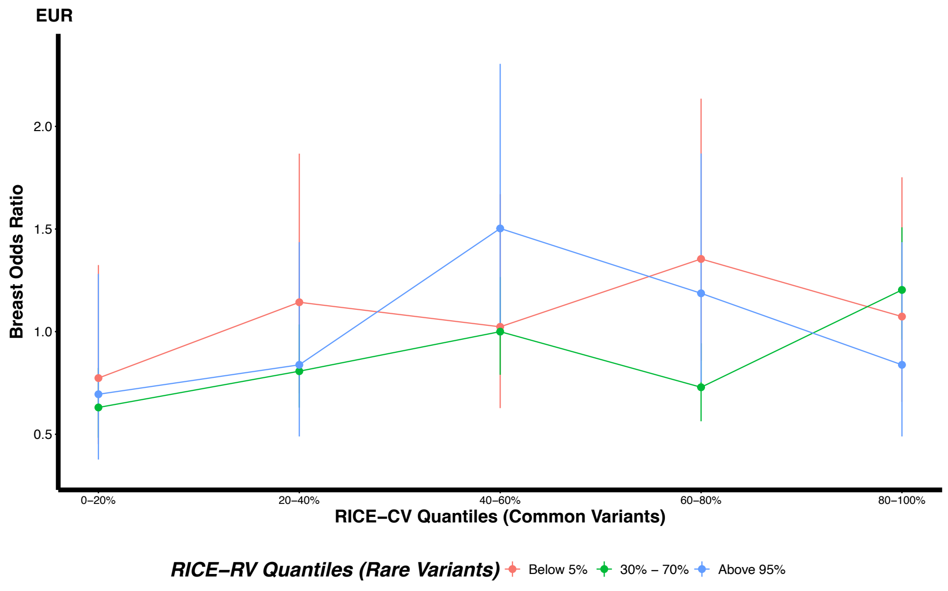


**c)** Relationship between ancestry-adjusted common and rare variant PRSs and the estimated odds ratio of coronary artery disease (CAD) for Europeans from UKB WES data.

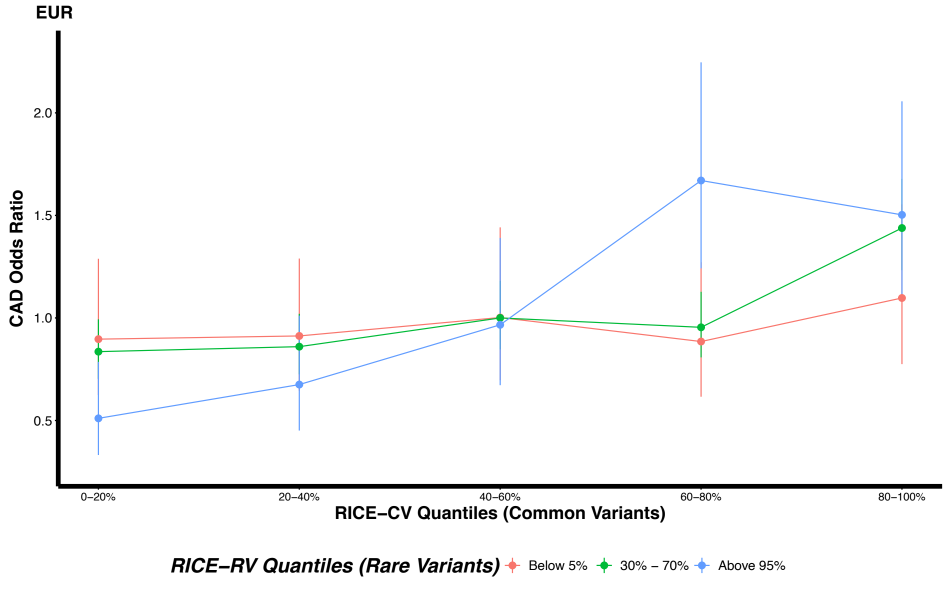

**Supplementary Figure 5 continued. d)** Relationship between ancestry-adjusted common and rare variant PRSs and the estimated odds ratio of prostate cancer for Europeans from UKB WES data.

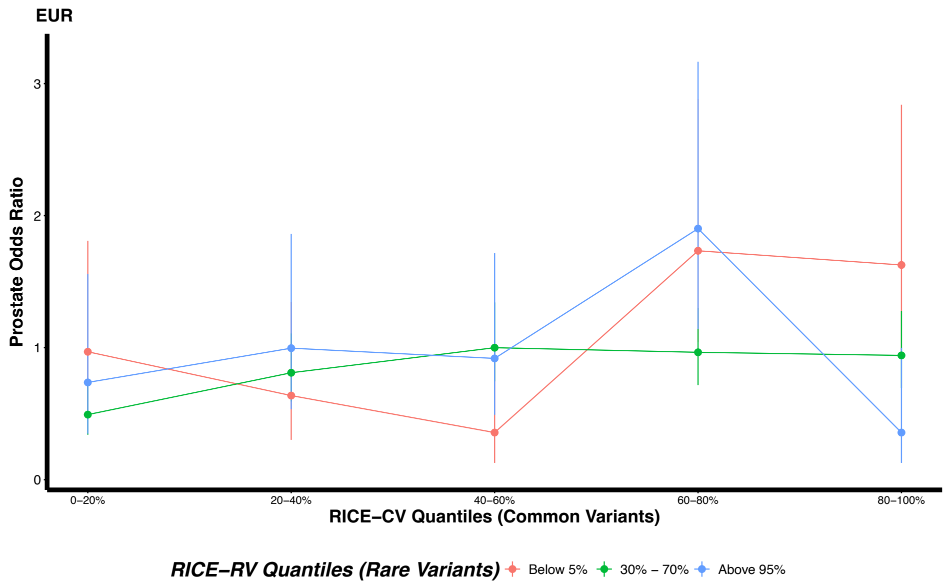


**e)** Relationship between ancestry-adjusted common and rare variant PRSs and the estimated odds ratio of type 2 diabetes (T2D) for Europeans from UKB WES data.

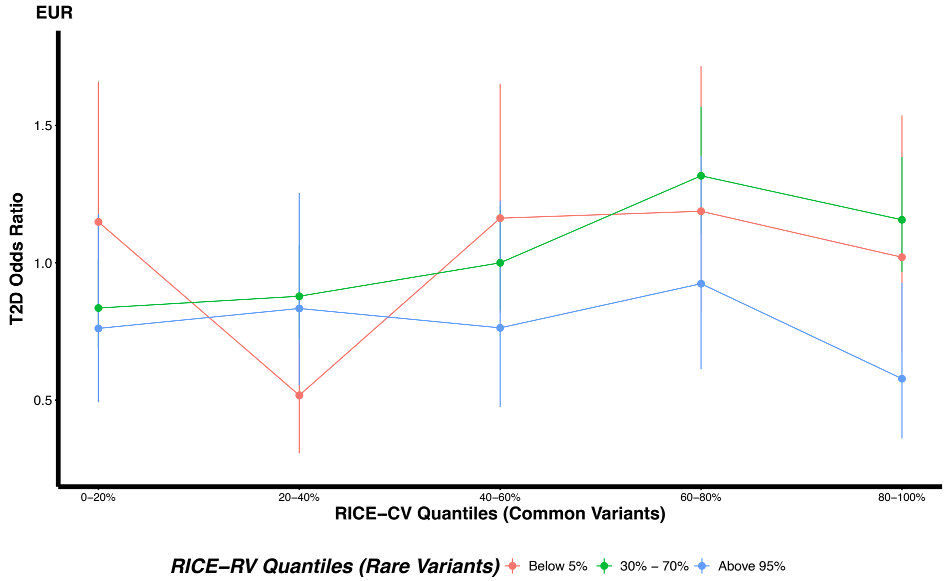

**Supplementary Figure 6. Relationship between common and rare variant PRSs and standardized traits across four ancestral groups from UK Biobank (UKB) whole-exome sequencing (WES) data.** The six continuous traits analyzed and shown: body mass index (BMI) (Supp. Fig. 6a), high-density lipoprotein cholesterol (HDL) (Supp. Fig. 6b), height (Supp. Fig. 6c), low-density lipoprotein cholesterol (LDL) (Supp. Fig. 6d), natural logarithm of triglyceride cholesterol (log(TG)) (Supp. Fig. 6e), and total cholesterol (TC) (Supp. Fig. 6f). PRS quantiles for RICE-CV (common variants) are plotted on the x-axis, and standardized trait on the y-axis. Data are stratified by rare variant PRS quantiles from RICE-RV (red: below 5%, green: 20–70%, blue: above 95%). Results are shown for individuals of African (AFR), Admixed American/Latino (AMR), European (EUR), and South Asian (SAS) ancestries. The training data consisted solely of individuals of European ancestry, while the tuning and validation sets included all four ancestries. Full sample sizes details for each ancestry are provided in **Supplementary Table 2**.

**a)** Relationship between ancestry-adjusted common and rare variant PRSs and standardized body mass index (BMI) levels across four ancestral groups from UKB WES data.

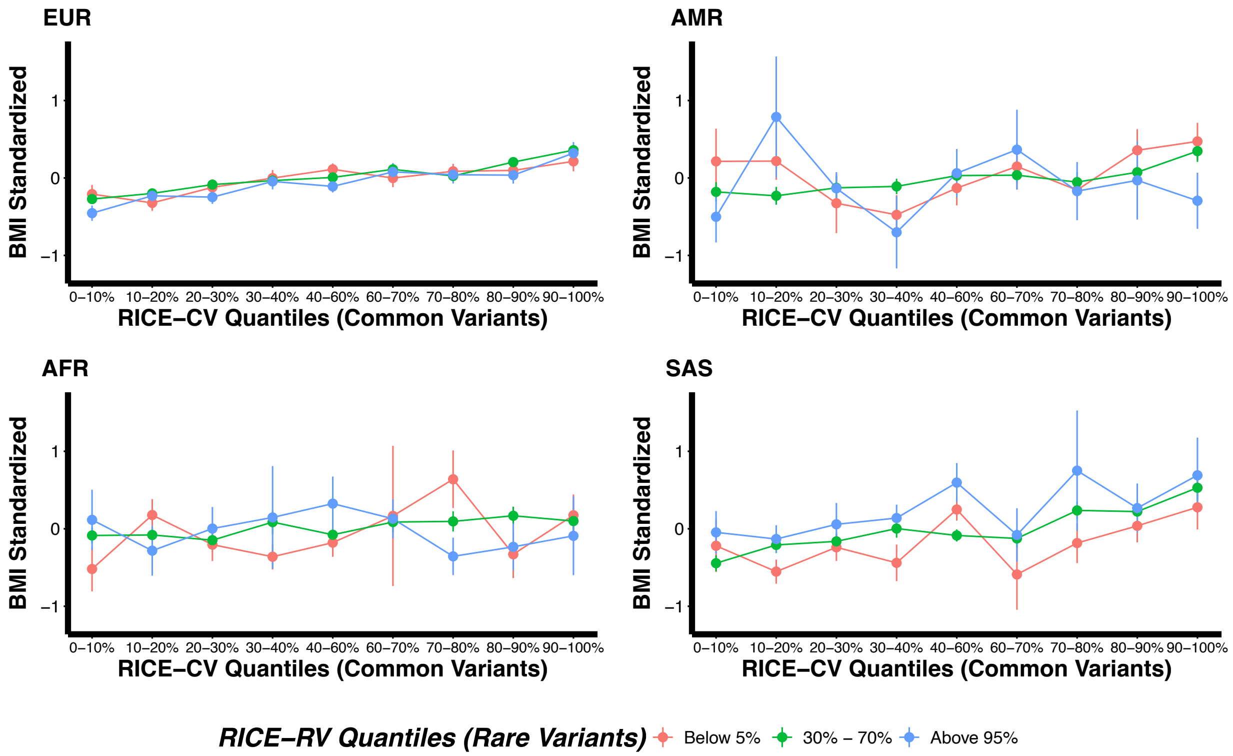

**Supplementary Figure 6 continued. b)** Relationship between ancestry-adjusted common and rare variant PRSs and standardized high-density lipoprotein cholesterol (HDL) levels across four ancestral groups from UKB WES data.

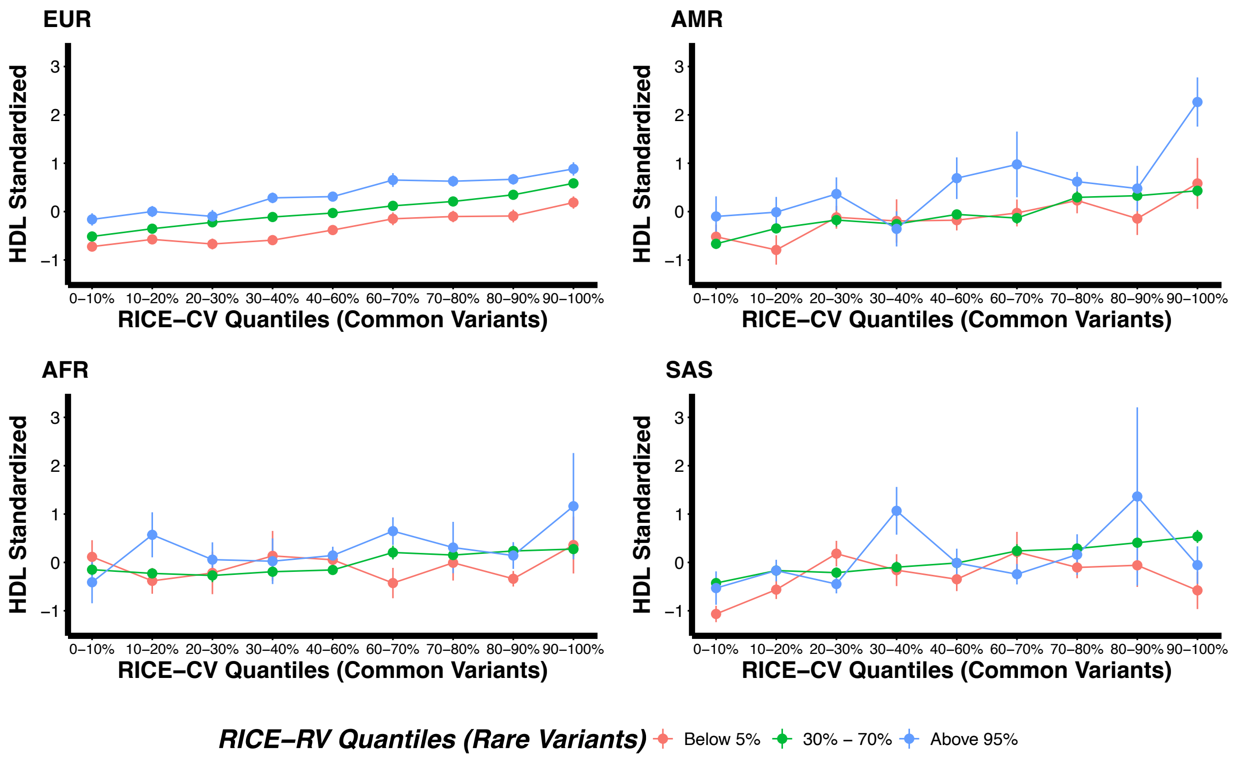

**Supplementary Figure 6 continued. c)** Relationship between ancestry-adjusted common and rare variant PRSs and standardized height across four ancestral groups from UKB WES data.

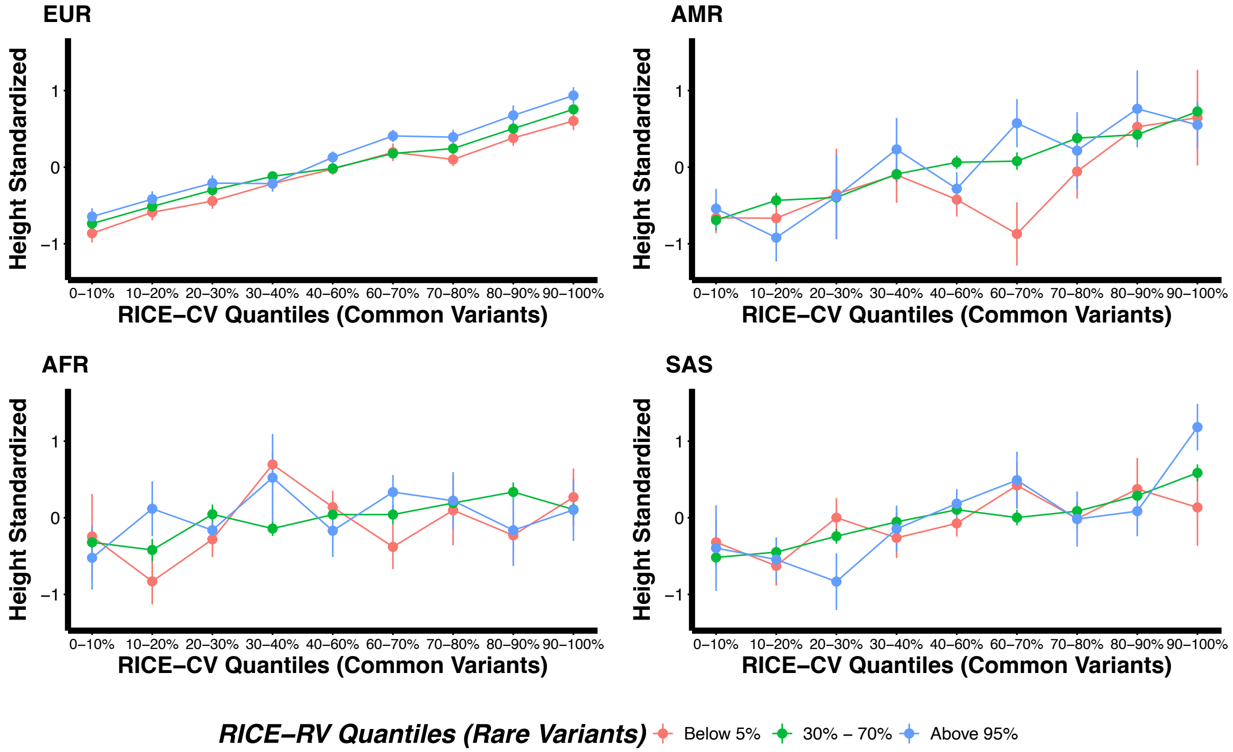

**Supplementary Figure 6 continued. d)** Relationship between ancestry-adjusted common and rare variant PRSs and standardized low-density lipoprotein cholesterol (LDL) levels across four ancestral groups from UKB WES data.

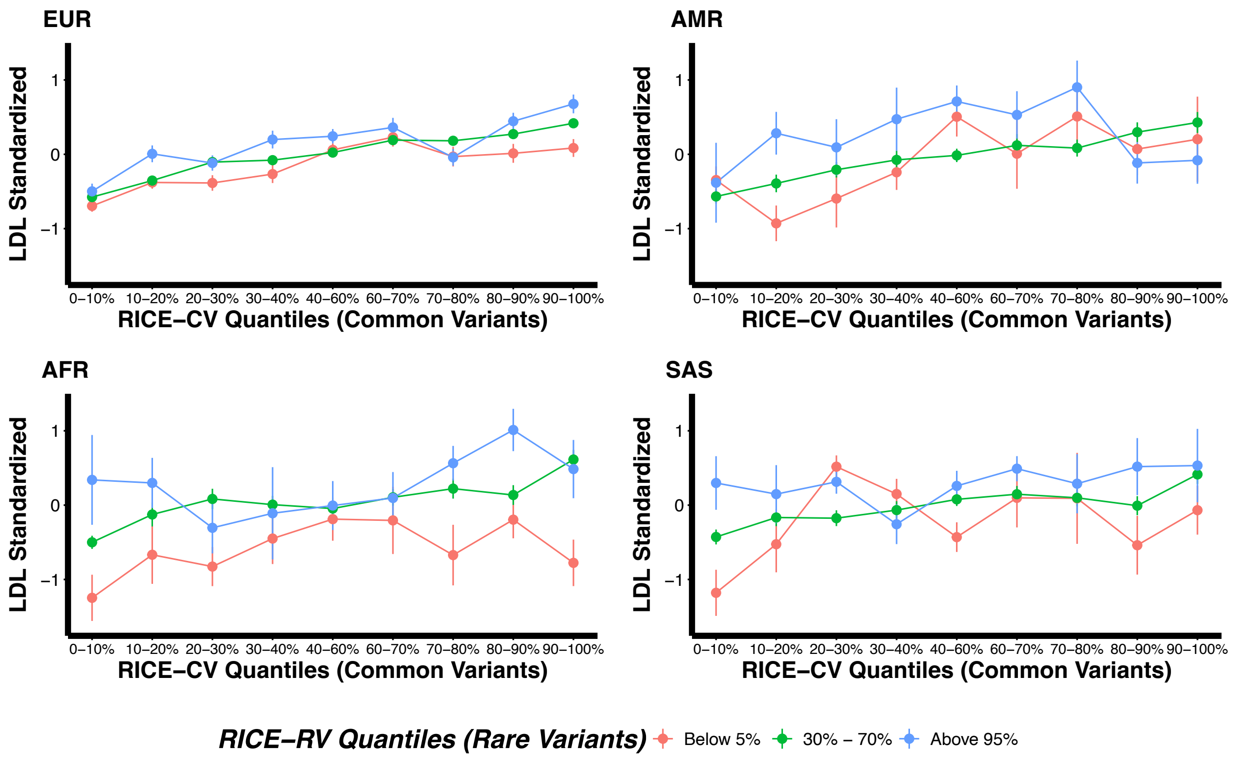

**Supplementary Figure 6 continued. e)** Relationship between ancestry-adjusted common and rare variant PRSs and standardized natural logarithm of triglycerides (log(TG)) levels across four ancestral groups from UKB WES data.

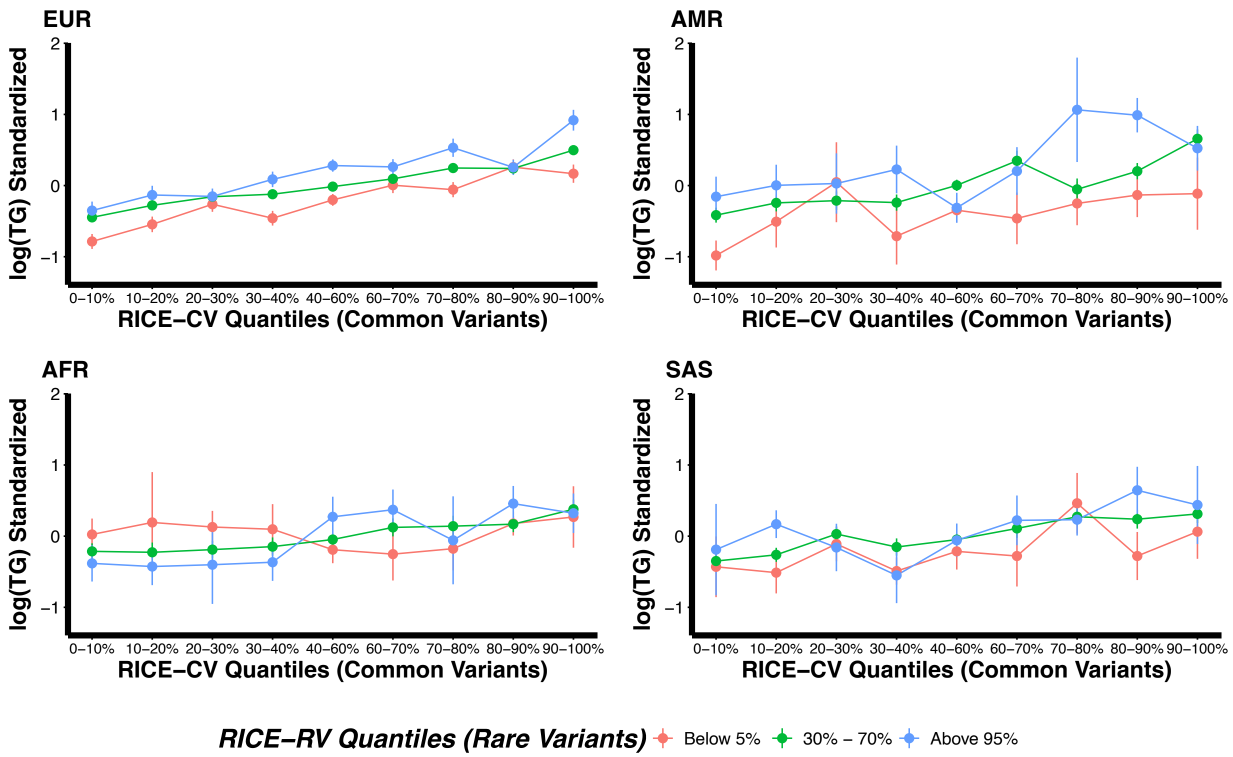

**Supplementary Figure 6 continued. f)** Relationship between ancestry-adjusted common and rare variant PRSs and standardized total cholesterol (TC) levels across four ancestral groups from UKB WES data.

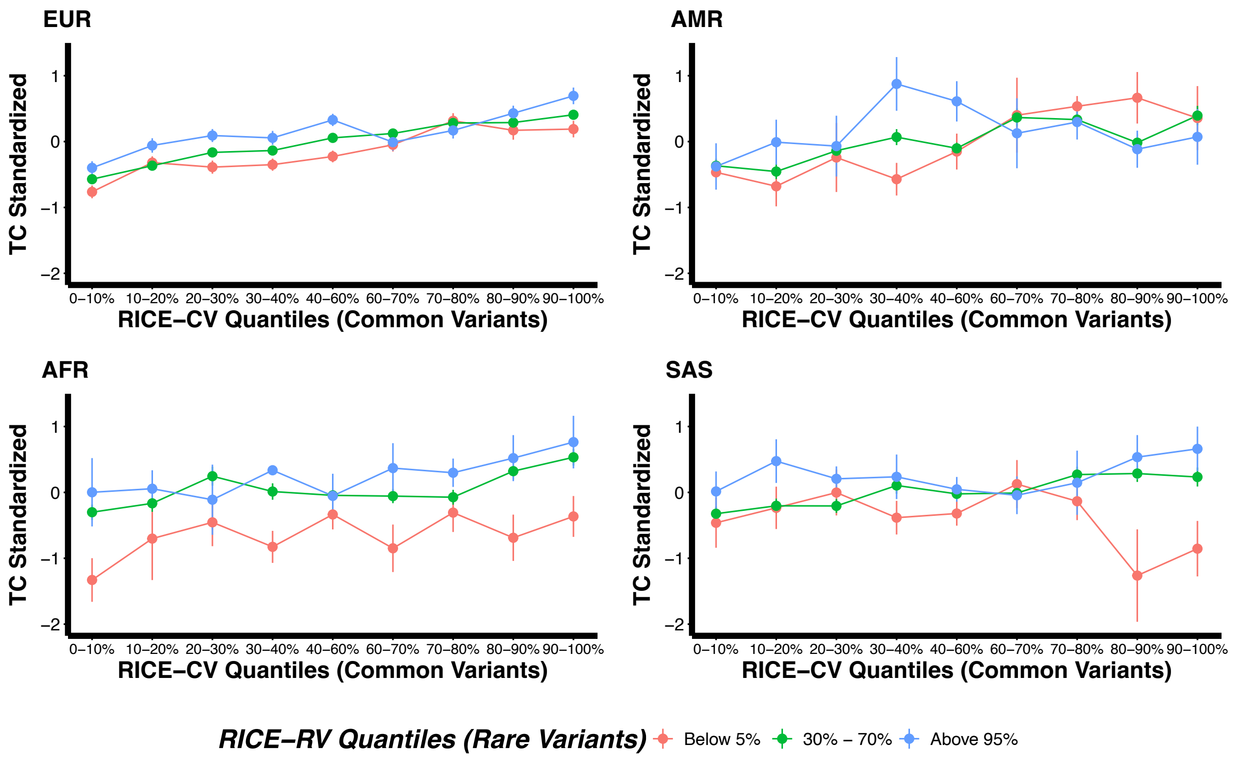

**Supplementary Figure 7.** Manhattan plot and QQ plots based on the UK Biobank whole exome sequencing (WES) GWAS summary statistics computed using the training set consisting of only individuals of European ancestry (EUR) for five binary traits: asthma, breast cancer, coronary artery disease (CAD), prostate cancer, and type 2 diabetes (T2D). The red and blue shaded regions around the diagonal line in the QQ plots indicate the 95% confidence intervals expected under the null hypothesis of no association between genetic variants and the trait of interest, for minor allele frequencies (MAF) within the ranges (0.05, 0.5] and [0.01, 0.05], respectively. Under the null hypothesis, the p-value follows a uniform (0,1) distribution. The jth order statistic follows a Beta (j, N-j+1) distribution, where N is the total number of variants given a specific MAF cutoff. For binary traits, $\lambda_{1000}$ scales $\lambda$ to a study with 1000 cases and 1000 controls using $\lambda_{1000} = 1 + 1000 \times (\lambda - 1) \times \left(\frac{1}{N_{case}} + \frac{1}{N_{control}}\right)$. Genomic control factors are shown in **Supplementary Table 5**.

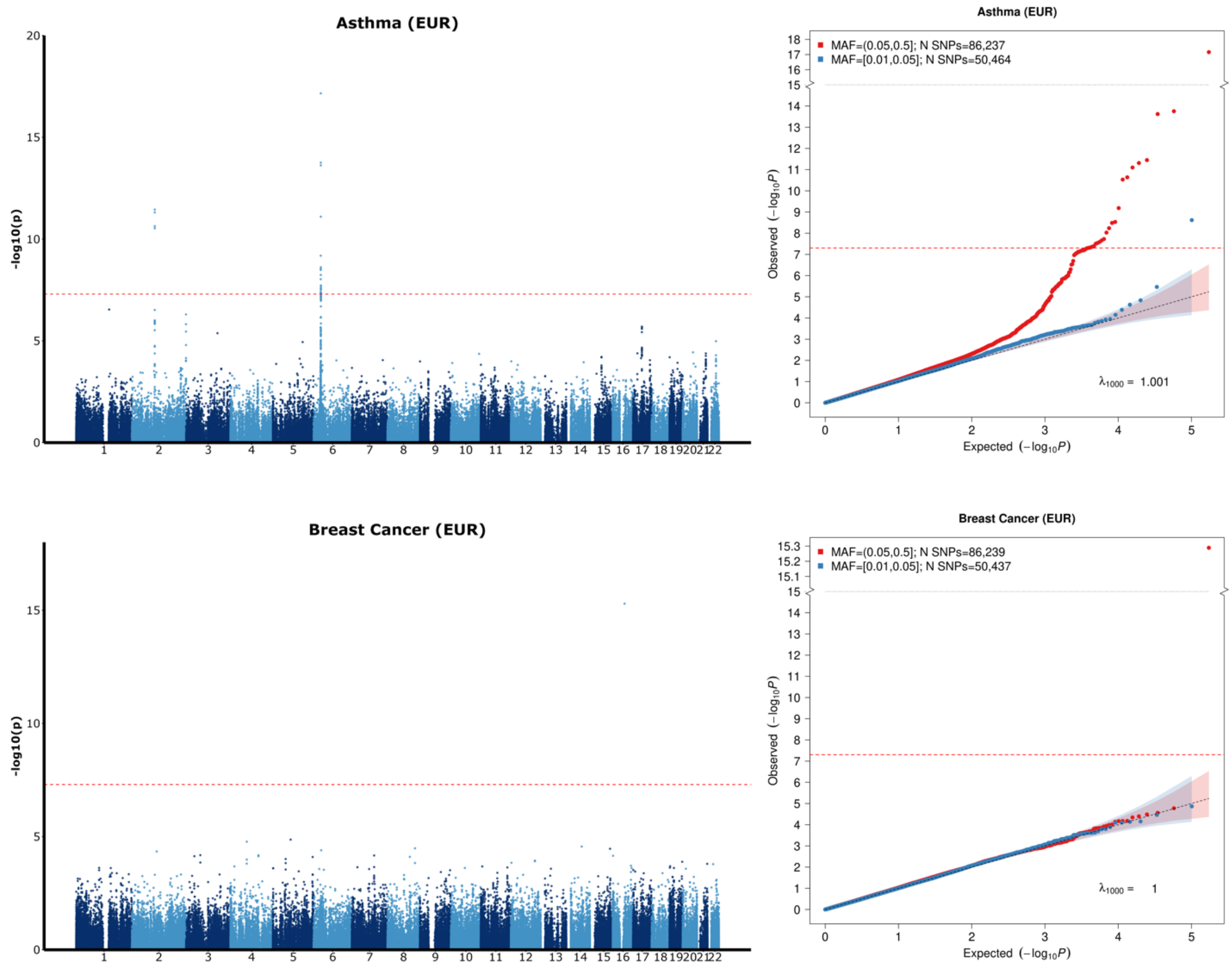

**Supplementary Figure 7 continued:** Manhattan and QQ Plots for CVD, prostate cancer and T2D based on UK Biobank WES GWAS summary statistics in European populations.

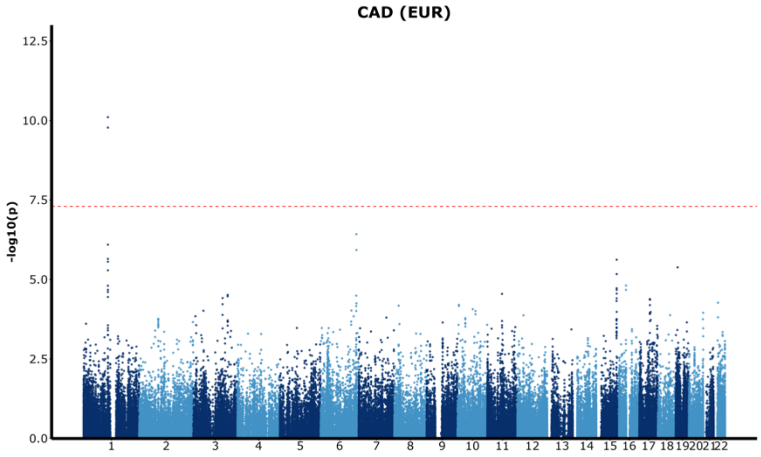


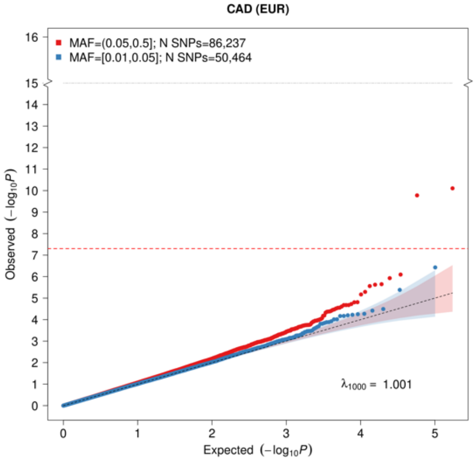


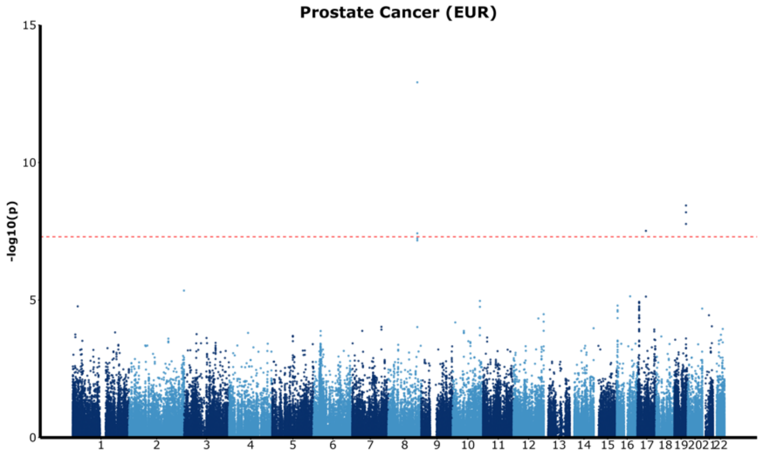


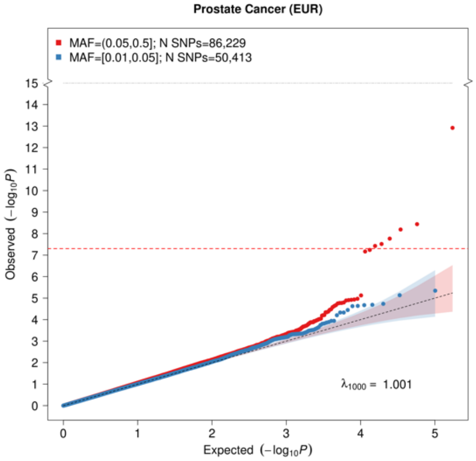


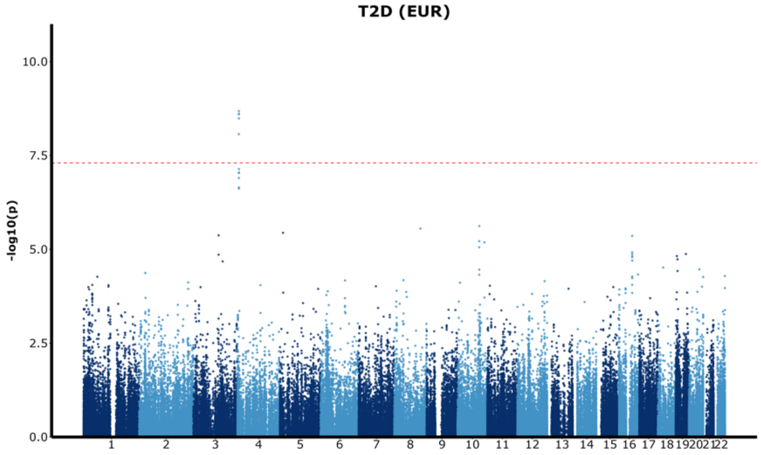


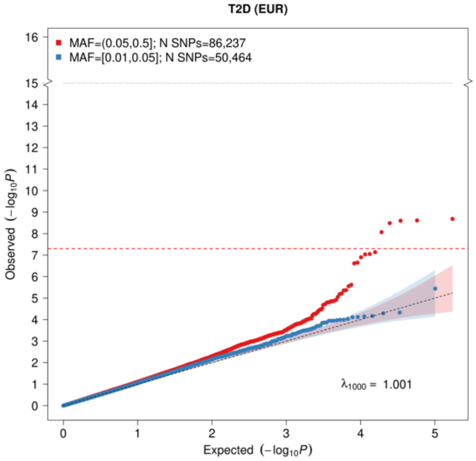

**Supplementary Figure 8.** Manhattan plot and QQ plots based on the UK Biobank whole exome sequencing (WES) GWAS summary statistics computed using the training set consisting of only individuals of European ancestry (EUR) for six continuous traits: body mass index (BMI), high-density lipoprotein cholesterol (HDL), height, low-density lipoprotein cholesterol (LDL), natural logarithm of triglycerides (log(TG)), and total cholesterol (TC). The red and blue shaded regions around the diagonal line in the QQ plots indicate the 95% confidence intervals expected under the null hypothesis of no association between genetic markers and the trait of interest, for minor allele frequencies (MAF) within the ranges (0.05, 0.5] and [0.01, 0.05], respectively. Under the null hypothesis, the p-value follows a uniform (0,1) distribution. The jth order statistic follows a Beta (j, N-j+1) distribution, where N is the total number of variants given a specific MAF cutoff. For continuous traits, $\lambda_{1000}$ scales $\lambda$ to a study with 1000 cases and 1000 controls using $\lambda_{1000} = 1 + 1000 \times (\lambda - 1)/\mathrm{N}$. Genomic control factors are shown in **Supplementary Table 5**.

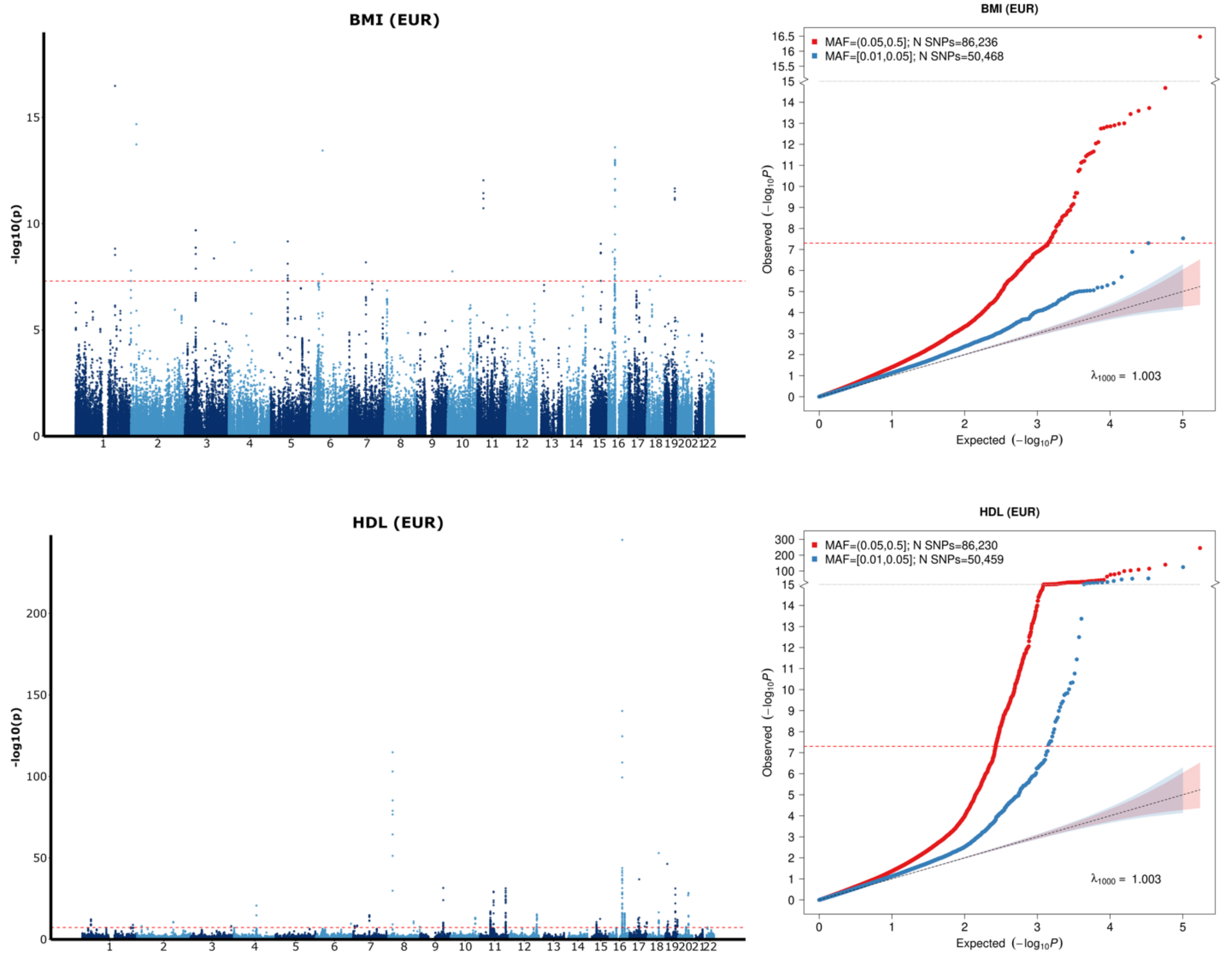

**Supplementary Figure 8 (continued):** Manhattan and QQ Plots for Height, LDL, and log(TG) based on UK Biobank WES GWAS summary statistics in European populations.

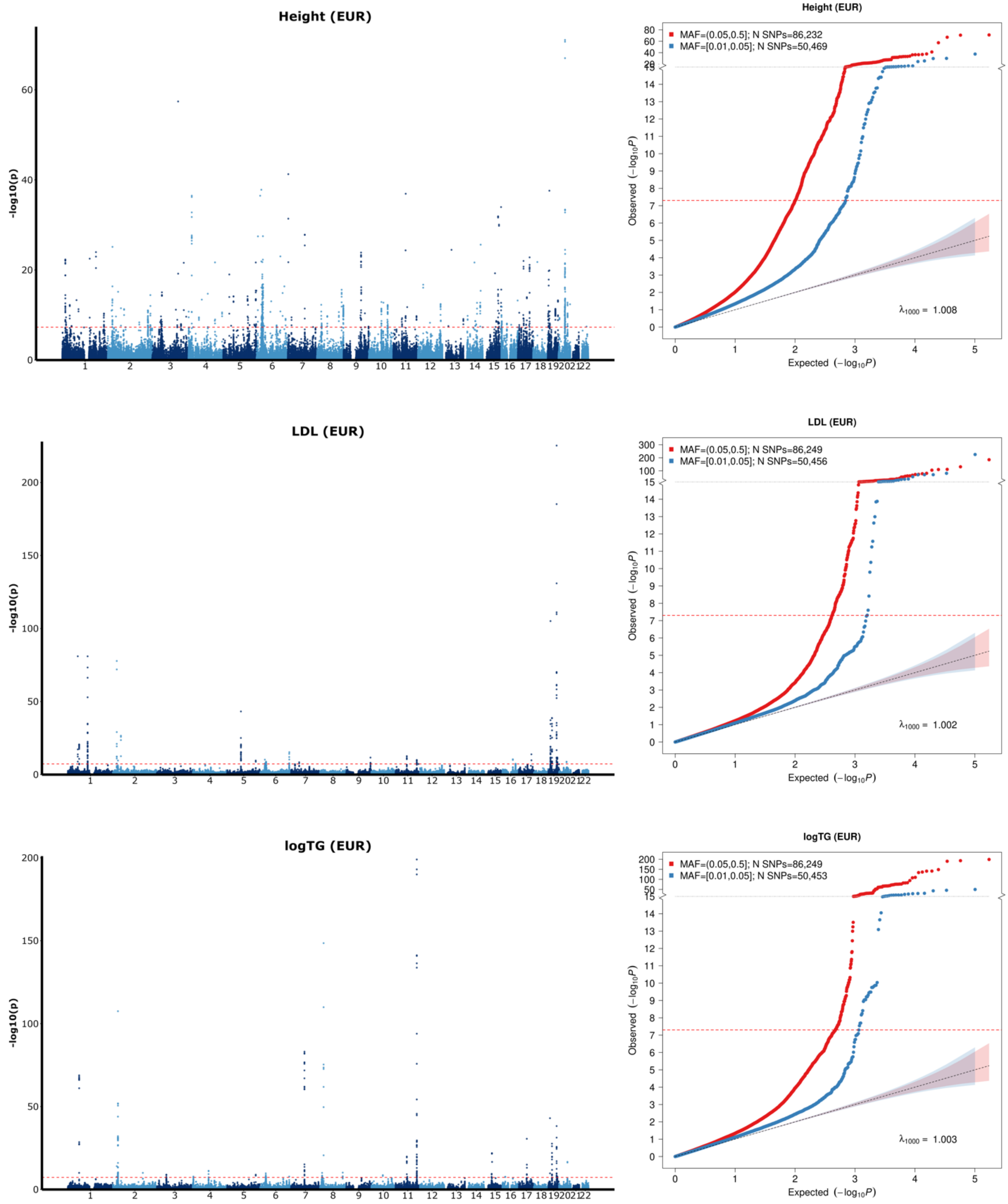

**Supplementary Figure 8 continued:** Manhattan and QQ Plots for TC based on UK Biobank WES GWAS summary statistics in European populations.

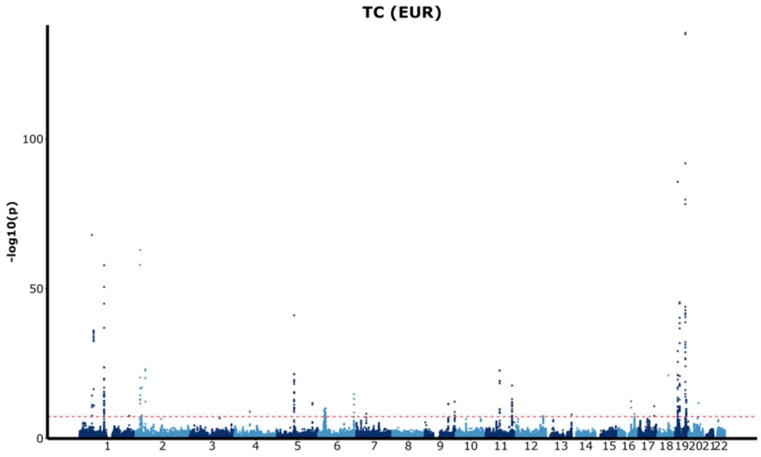


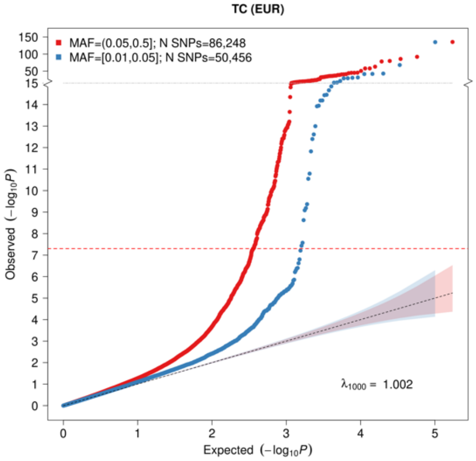

**Supplementary Figure 9.** QQ plots based on the UK Biobank whole exome sequencing rare variant association analysis for coding genes conducted with STAARpipeline using the training set consisting of only individuals of European ancestry (EUR) for 11 traits (sample sizes provided in **Supplementary Table 2**). The five binary traits analyzed and displayed in Supplementary Figure 9a: asthma, breast cancer, coronary artery disease (CAD), prostate cancer, and type 2 diabetes (T2D) and the six continuous traits analyzed and displayed in Supplementary Figure 9b: body mass index (BMI), high-density lipoprotein cholesterol (HDL), height, low-density lipoprotein cholesterol (LDL), natural logarithm of triglycerides (log(TG)), and total cholesterol (TC). P-values are split into the five functional categories investigated in the gene-centric coding analysis in the STAARpipeline: putative loss of function (pLoF), putative loss of function and disruptive (pLoF+D), missense, disruptive missense, and synonymous. Under the null hypothesis, the p-value follows a uniform (0,1) distribution.

**a)** QQ plots for five binary traits from the UKB WES rare variant association analysis using coding genes with STAARpipeline.

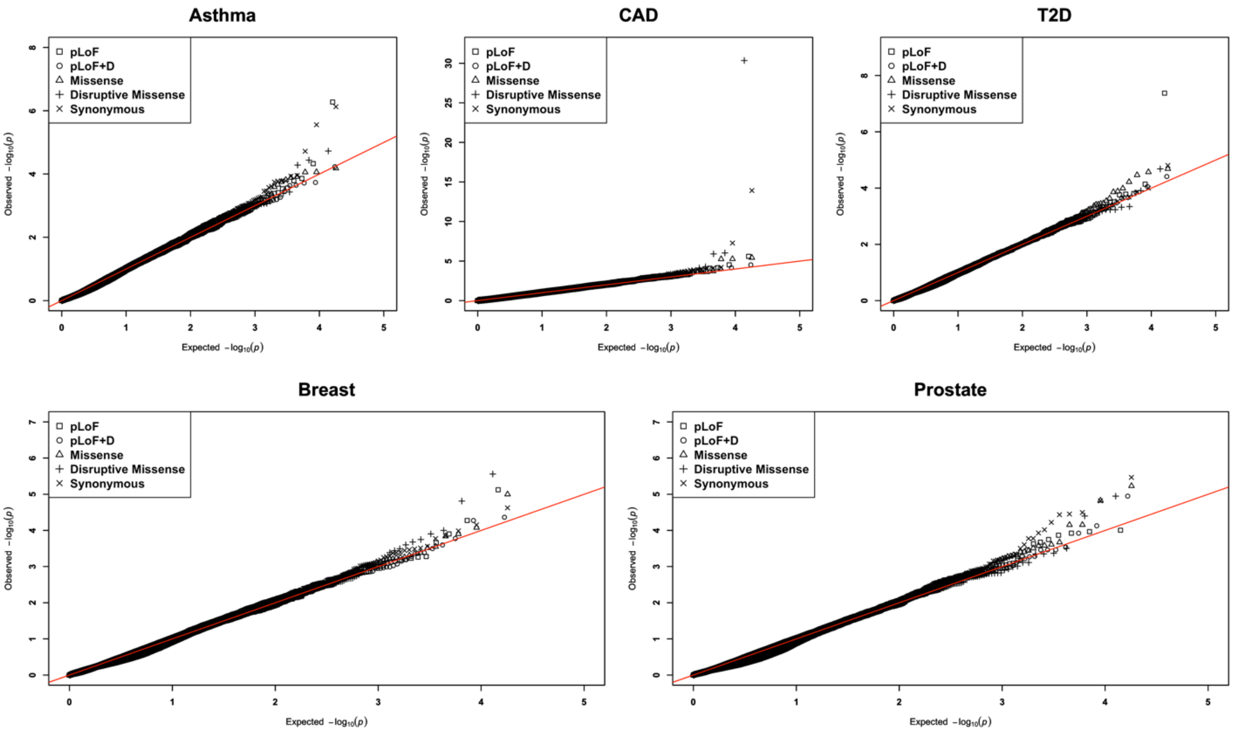

**Supplementary Figure 9 continued. b)** QQ plots for six continuous traits from the UKB WES rare variant association analysis using coding genes with STAARpipeline.

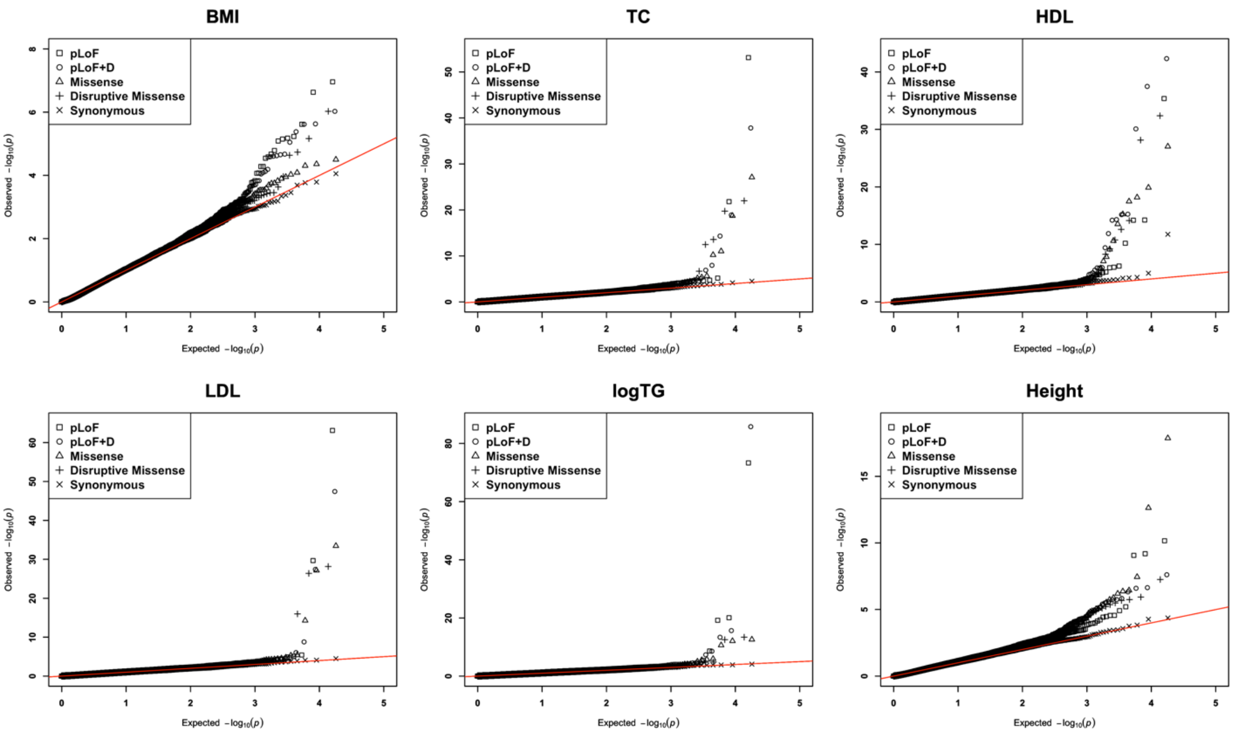

**Supplementary Figure 10. Predictive performance of ancestry-adjusted PRSs for 11 traits across four ancestral groups from UK Biobank (UKB) whole-genome sequencing (WGS) data.** The five binary traits analyzed and displayed in Supplementary Figure 10a: asthma, breast cancer, coronary artery disease (CAD), prostate cancer, and type 2 diabetes (T2D). The six continuous traits analyzed and displayed in Supplementary Figure 10b include body mass index (BMI), height, high-density lipoproteins cholesterol (HDL), low-density lipoproteins cholesterol (LDL), the natural logarithm of triglyceride cholesterol (log(TG)), and total cholesterol (TC). Results are shown for individuals of African (AFR), Admixed American or Latino (AMR), European (EUR), and South Asian (SAS) ancestries. The training data consisted solely of individuals of European ancestry, while tuning and validation sets included all four ancestries. Full sample sizes details for each ancestry are provided in **Supplementary Table 3**.

**a)** Predictive performance of ancestry-adjusted PRSs for five binary traits across four ancestral groups from UK Biobank (UKB) whole-genome sequencing (WGS) data

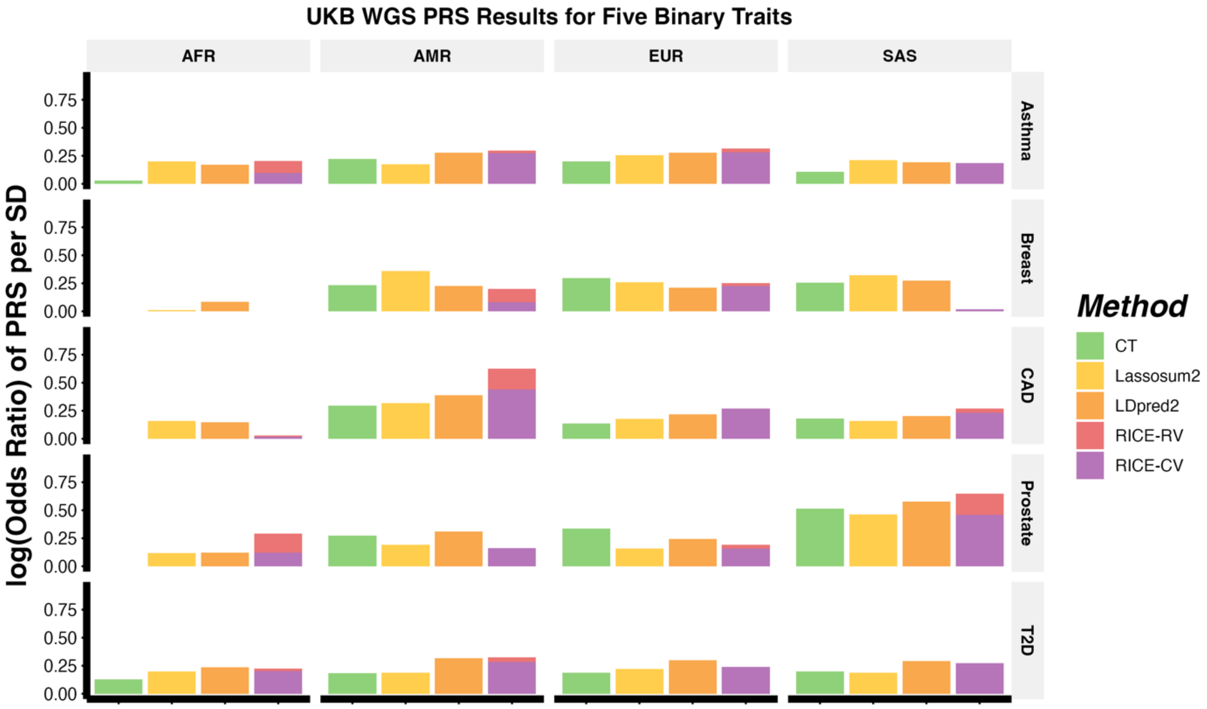

**Supplementary Figure 10 continued. b)** Predictive performance of ancestry-adjusted PRSs for six continuous traits across four ancestral groups from UK Biobank (UKB) whole-genome sequencing (WGS) data.

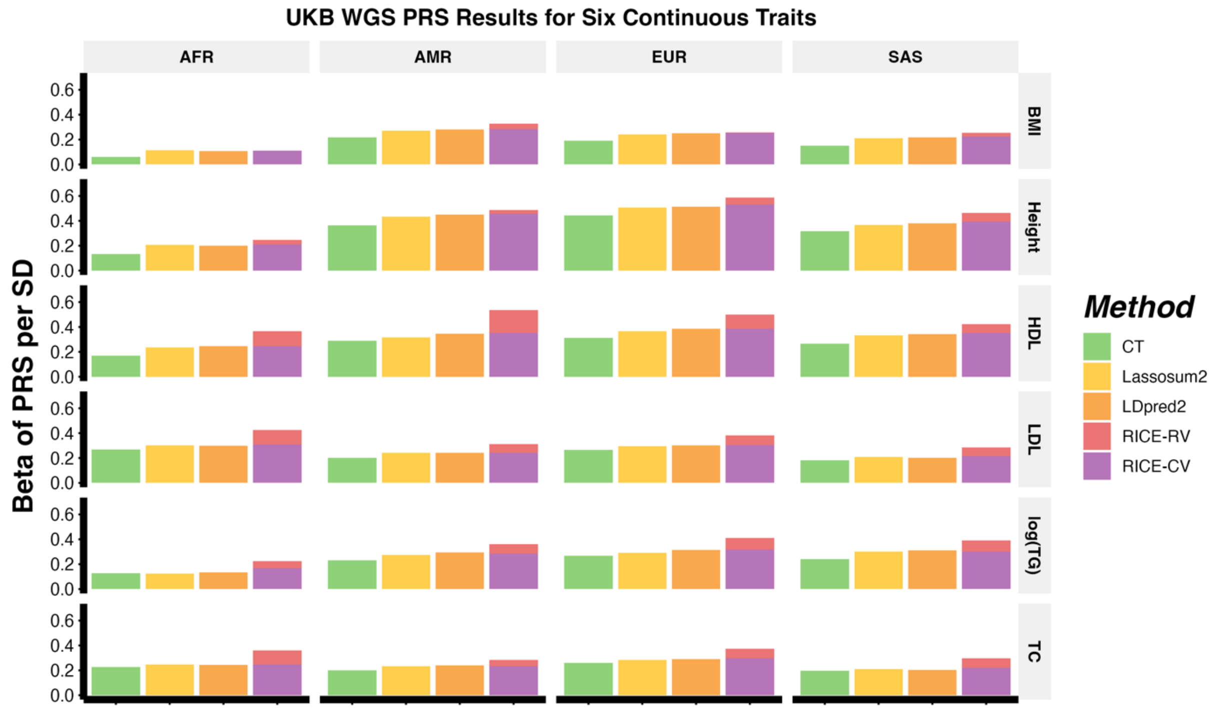

**Supplementary Figure 11. Relationship between common and rare variant PRSs and the estimated odds ratio for Europeans from UK Biobank (UKB) whole-genome sequencing (WGS) data.** Figures are shown for the five binary traits: asthma (Supp. Fig. 11a), breast cancer (Supp. Fig. 11b), coronary artery disease (CAD) (Supp. Fig. 11c), prostate cancer (Supp. Fig. 11d), and type 2 diabetes (Supp. Fig. 11e). PRS quantiles for RICE-CV (common variants) are plotted on the x-axis, and estimated odds ratio is on the y-axis. Data are stratified by rare variant PRS quantiles from RICE-RV (red: below 5%, green: 20–70%, blue: above 95%). Results are shown for individuals of European (EUR) ancestries. The training data consisted solely of individuals of European ancestry, while the tuning and validation sets included all four ancestries. Full sample sizes details for each ancestry are provided in **Supplementary Table 3**.

**a)** Relationship between ancestry-adjusted common and rare variant PRSs and the estimated odds ratio of asthma for Europeans from UKB WGS data.

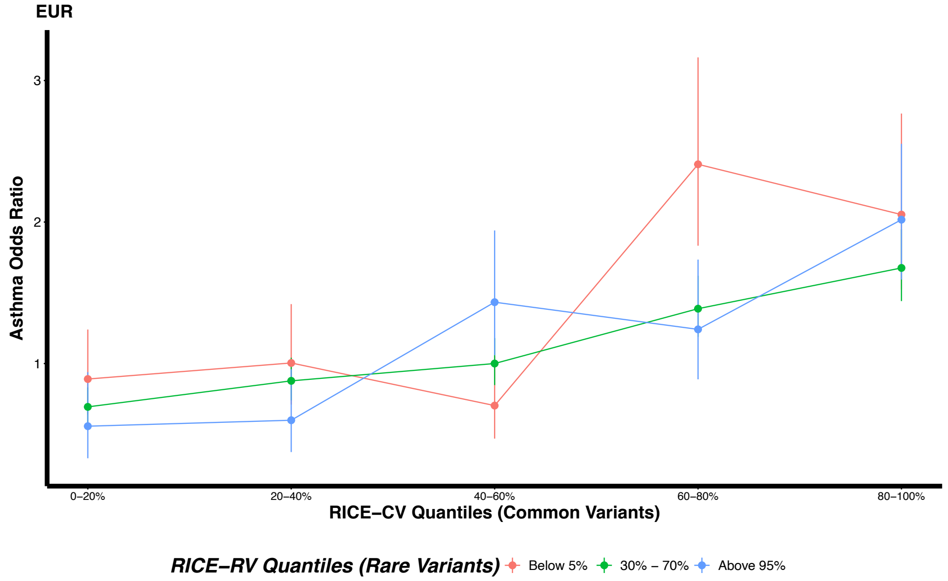

**Supplementary Figure 11 continued. b)** Relationship between ancestry-adjusted common and rare variant PRSs and the estimated odds ratio of breast cancer for Europeans from UKB WGS data.

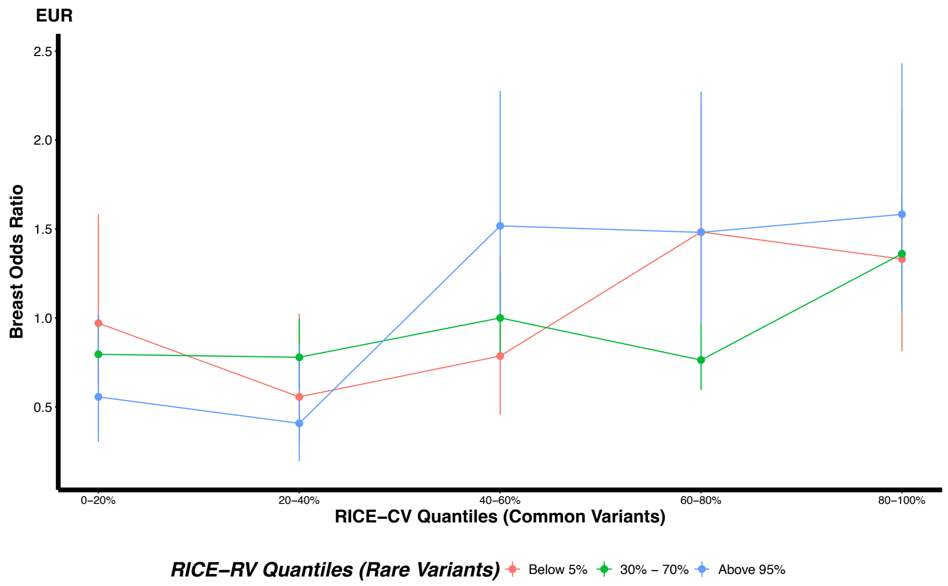


**c)** Relationship between ancestry-adjusted common and rare variant PRSs and the estimated odds ratio of coronary artery disease (CAD) for Europeans from UKB WGS data.

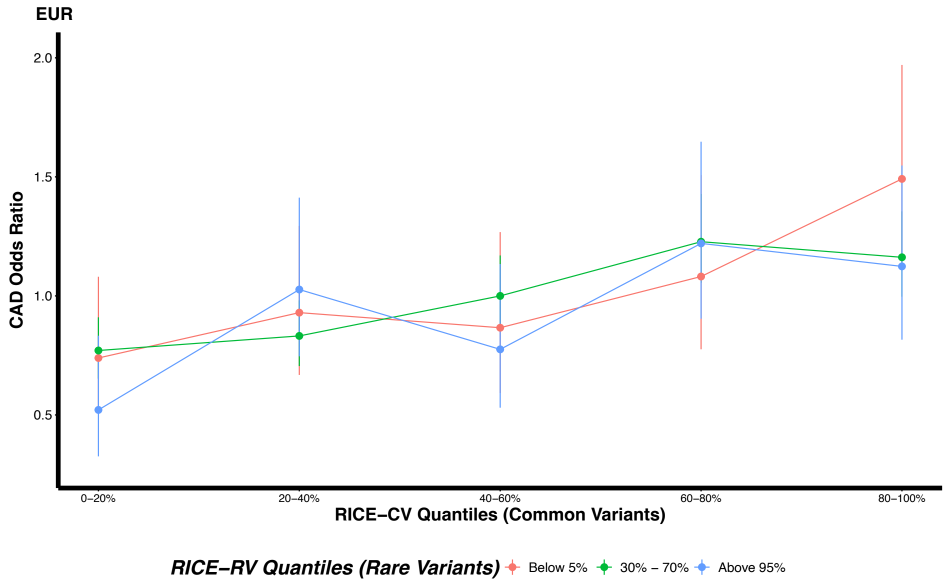

**Supplementary Figure 11 continued. d)** Relationship between ancestry-adjusted common and rare variant PRSs and the estimated odds ratio of prostate cancer for Europeans from UKB WGS data.

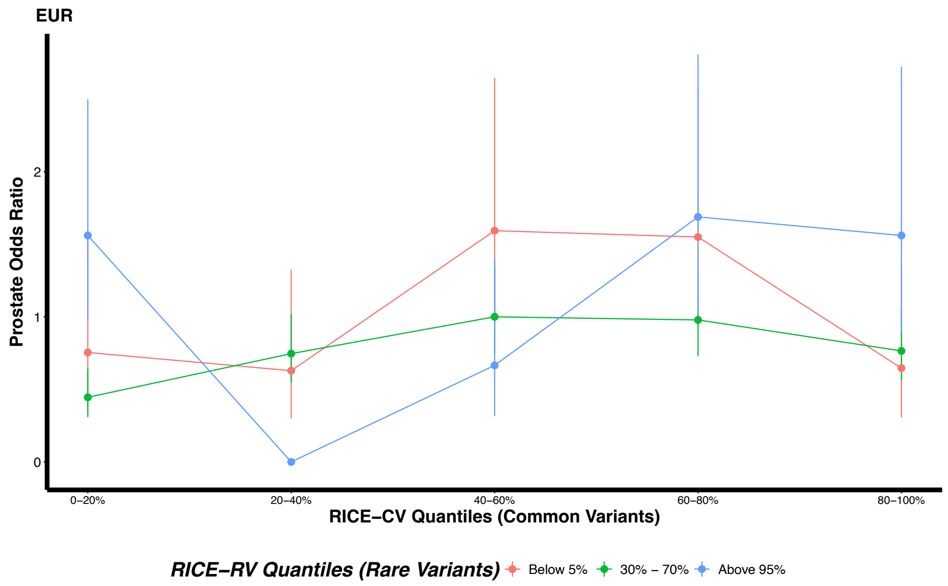


**e)** Relationship between ancestry-adjusted common and rare variant PRSs and the estimated odds ratio of type 2 diabetes (T2D) for Europeans from UKB WGS data.

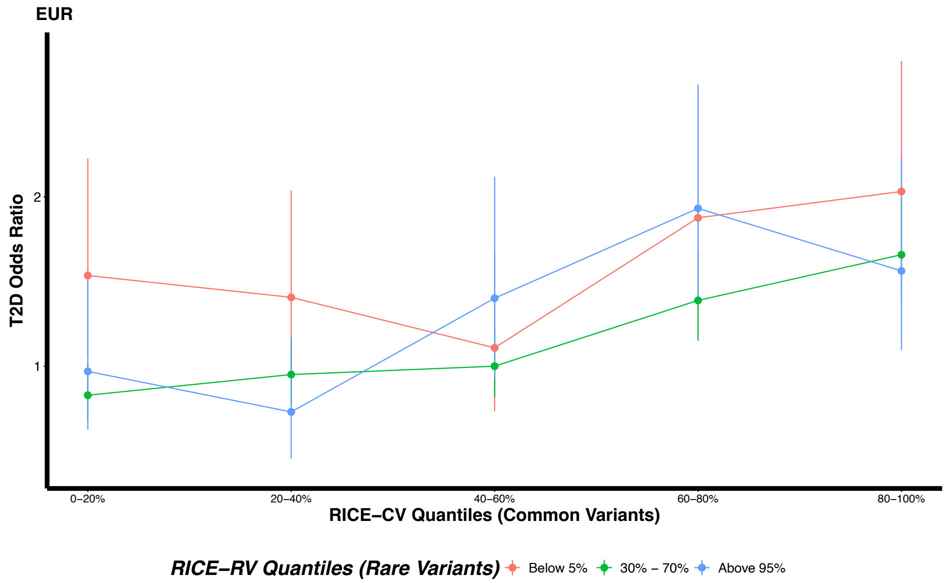

**Supplementary Figure 12. Relationship between common and rare variant PRSs and standardized traits across four ancestral groups from UK Biobank (UKB) whole-genome sequencing (WGS) data.** The six continuous traits analyzed and shown: body mass index (BMI) (Supp. Fig. 12a), height (Supp. Fig. 12b), low-density lipoprotein cholesterol (LDL) (Supp. Fig. 12c), natural logarithm of triglyceride cholesterol (log(TG)) (Supp. Fig. 12d), and total cholesterol (TC) (Supp. Fig. 12e). PRS quantiles for RICE-CV (common variants) are plotted on the x-axis, and standardized trait on the y-axis. Data are stratified by rare variant PRS quantiles from RICE-RV (red: below 5%, green: 20–70%, blue: above 95%). Results are shown for individuals of African (AFR), Admixed American/Latino (AMR), European (EUR), and South Asian (SAS) ancestries. The training data consisted solely of individuals of European ancestry, while the tuning and validation sets included all four ancestries. Full sample sizes details for each ancestry are provided in **Supplementary Table 3**.

**a)** Relationship between ancestry-adjusted common and rare variant PRSs and standardized body mass index (BMI) levels across four ancestral groups from UKB WGS data.

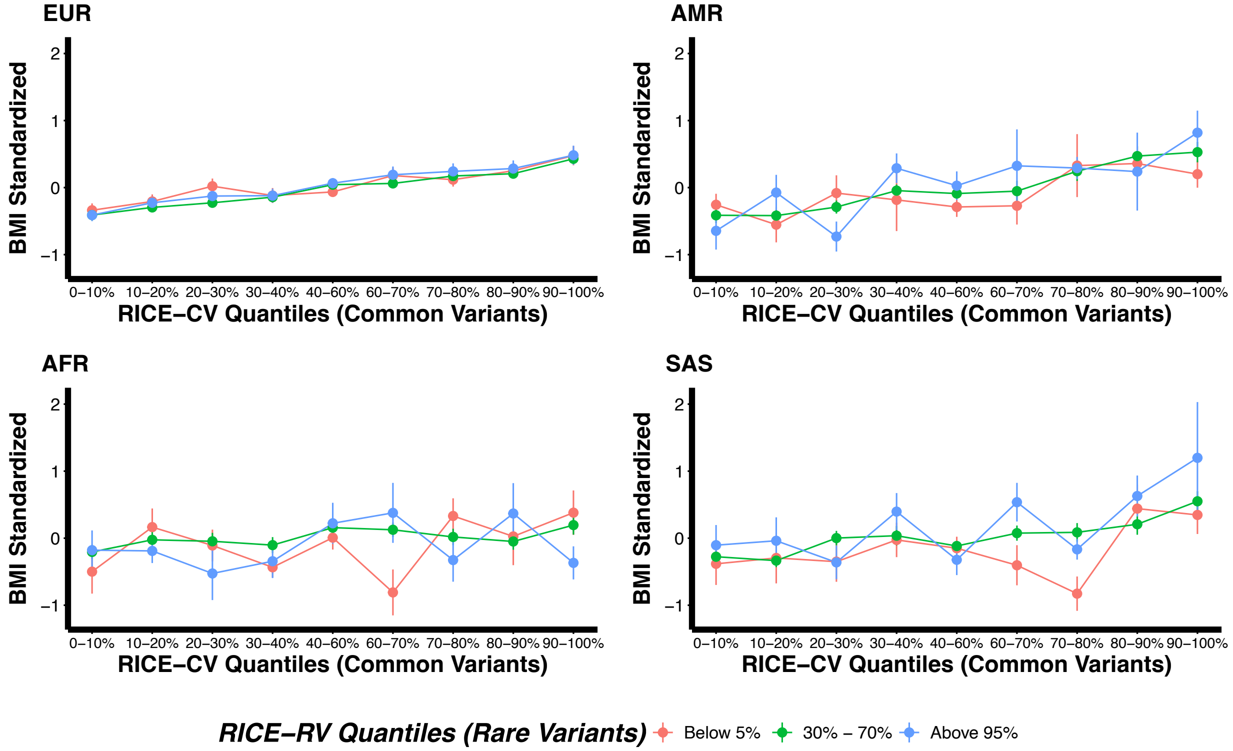

**Supplementary Figure 12 continued. b)** Relationship between ancestry-adjusted common and rare variant PRSs and standardized height across four ancestral groups from UKB WGS data.

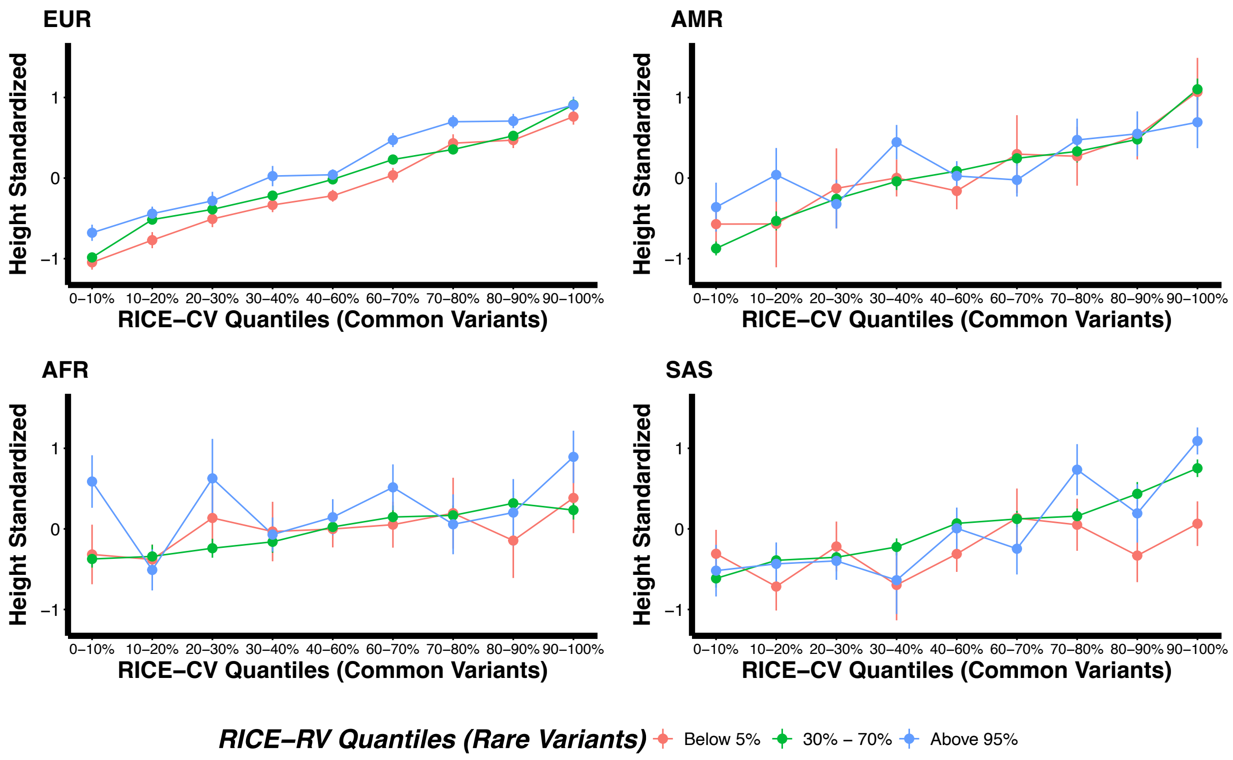

**Supplementary Figure 12 continued. c)** Relationship between ancestry-adjusted common and rare variant PRSs and standardized low-density lipoprotein cholesterol (LDL) levels across four ancestral groups from UKB WGS data.

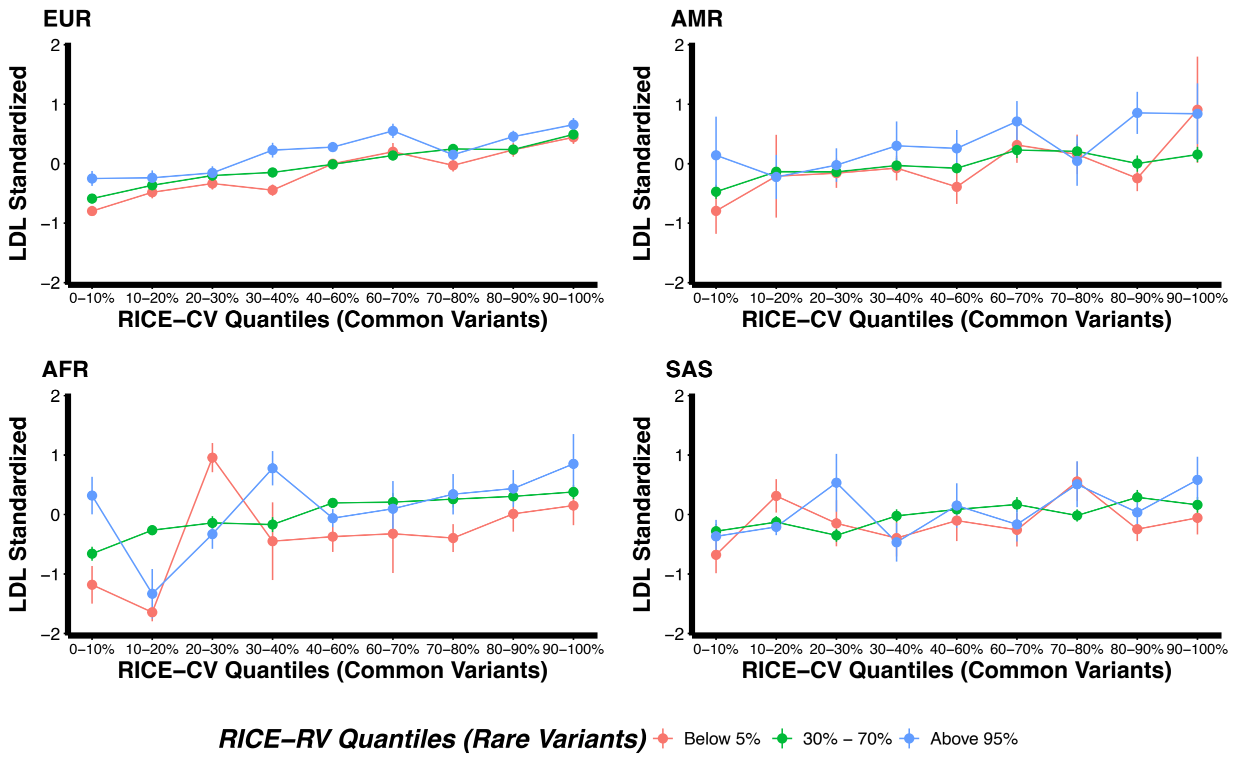

**Supplementary Figure 12 continued. d)** Relationship between ancestry-adjusted common and rare variant PRSs and standardized natural logarithm of triglycerides (log(TG)) levels across four ancestral groups from UKB WGS data.

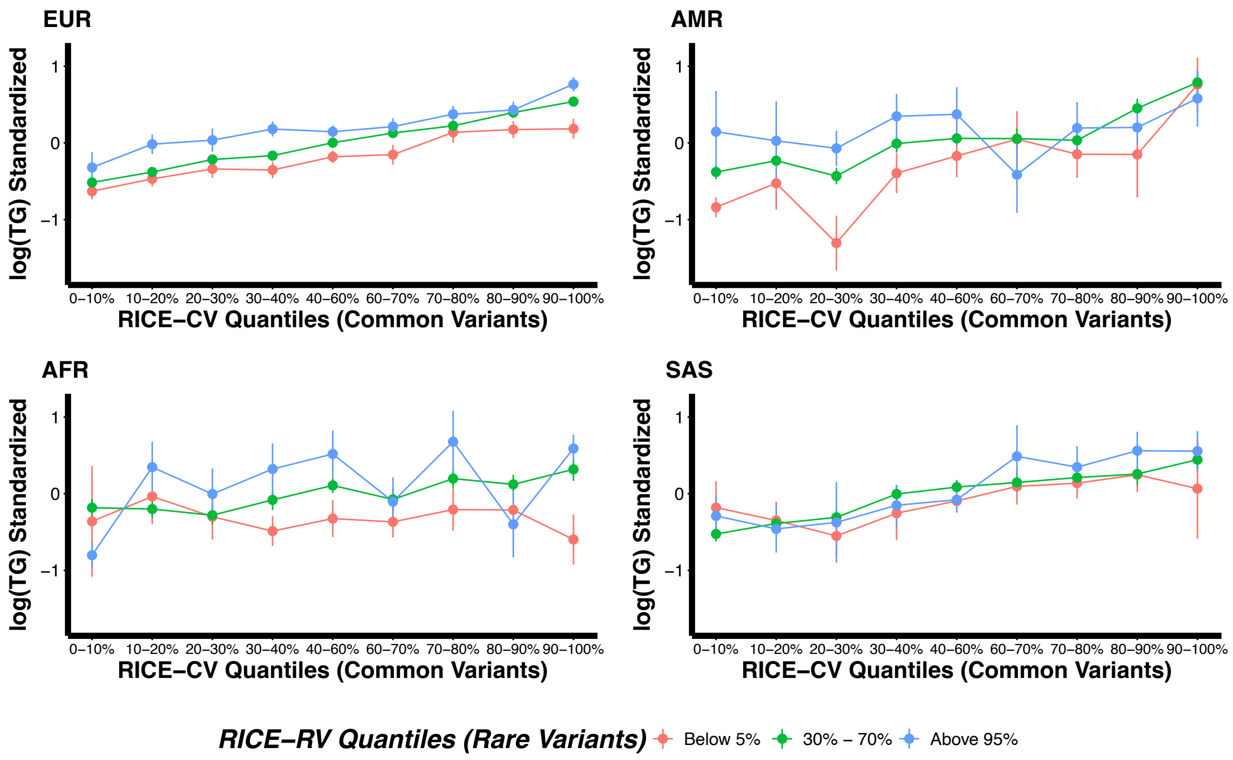

**Supplementary Figure 12 continued. e)** Relationship between ancestry-adjusted common and rare variant PRSs and standardized total cholesterol (TC) levels across four ancestral groups from UKB WGS data.

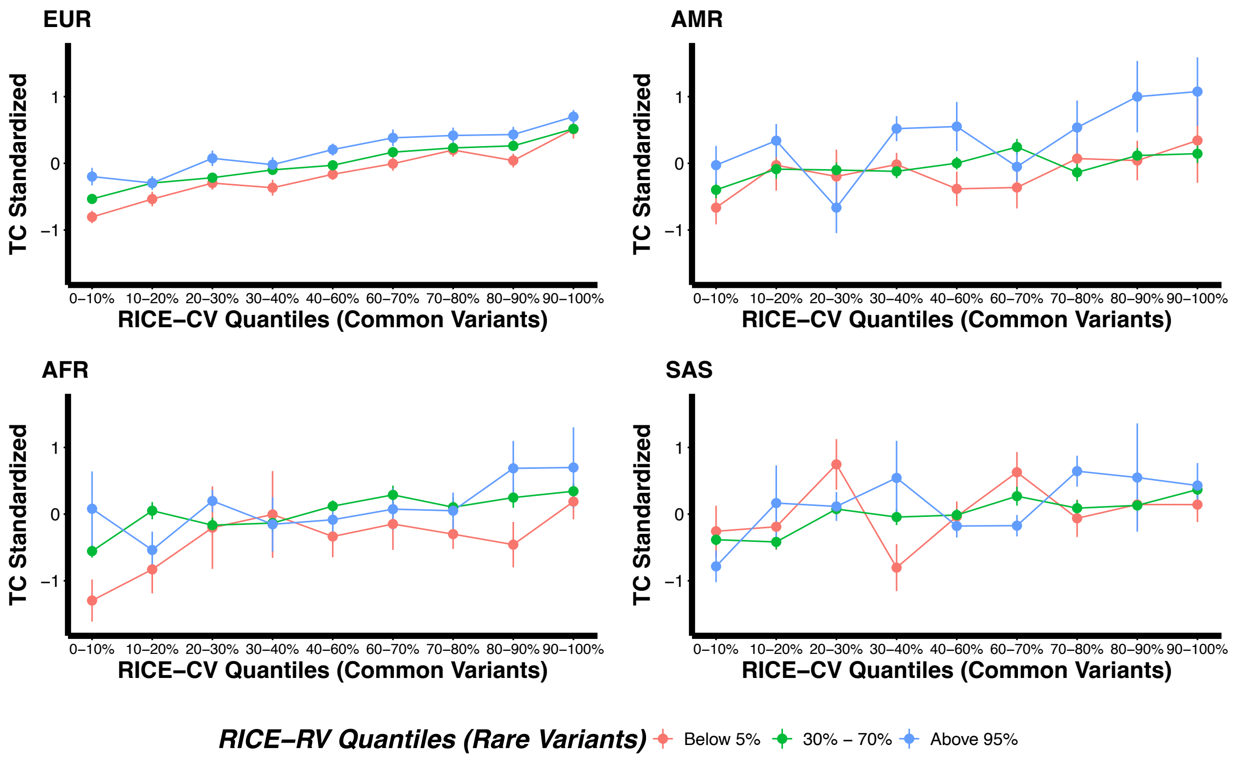

**Supplementary Figure 13. Comparison of ancestry-adjusted PRSs from RICE-CV and RICE-RV using UK Biobank (UKB) whole-exome sequencing (WES) and whole-genome sequencing (WGS) data for individuals of either African, Admixed American, or South Asian ancestry.** Results are shown for six continuous traits: body mass index (BMI), height, high-density lipoproteins (HDL), low-density lipoproteins (LDL), the natural logarithm of triglyceride (log(TG)), total cholesterol (TC), and five binary traits: asthma, breast cancer, coronary artery disease (CAD), prostate cancer, and type 2 diabetes (T2D). The training data included only individuals of European ancestry, while the tuning and validation sets contained individuals from all four ancestries. Full sample size details for each ancestry are provided in **Supplementary Tables 2 and 3**. Separate figures for individuals of African (Supp. Fig. 13a), Admixed American (Supp. Fig. 13b), and South Asian (Supp. Fig. 13c) ancestry are shown below.

**a)** Comparison of ancestry-adjusted PRSs from RICE-CV and RICE-RV using UKB WES and WGS data for individuals of African ancestry.

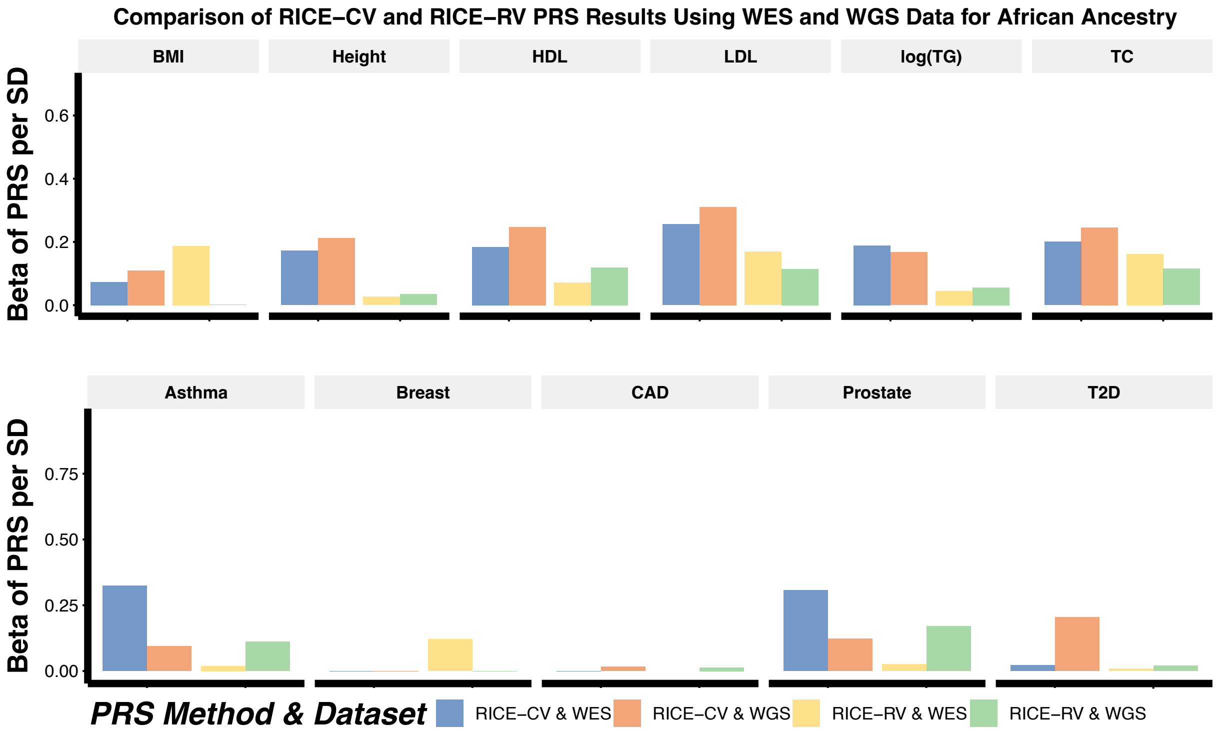

**Supplementary Figure 13 continued. b)** Comparison of ancestry-adjusted PRSs from RICE-CV and RICE-RV using UKB WES and WGS data for individuals of Admixed American ancestry.

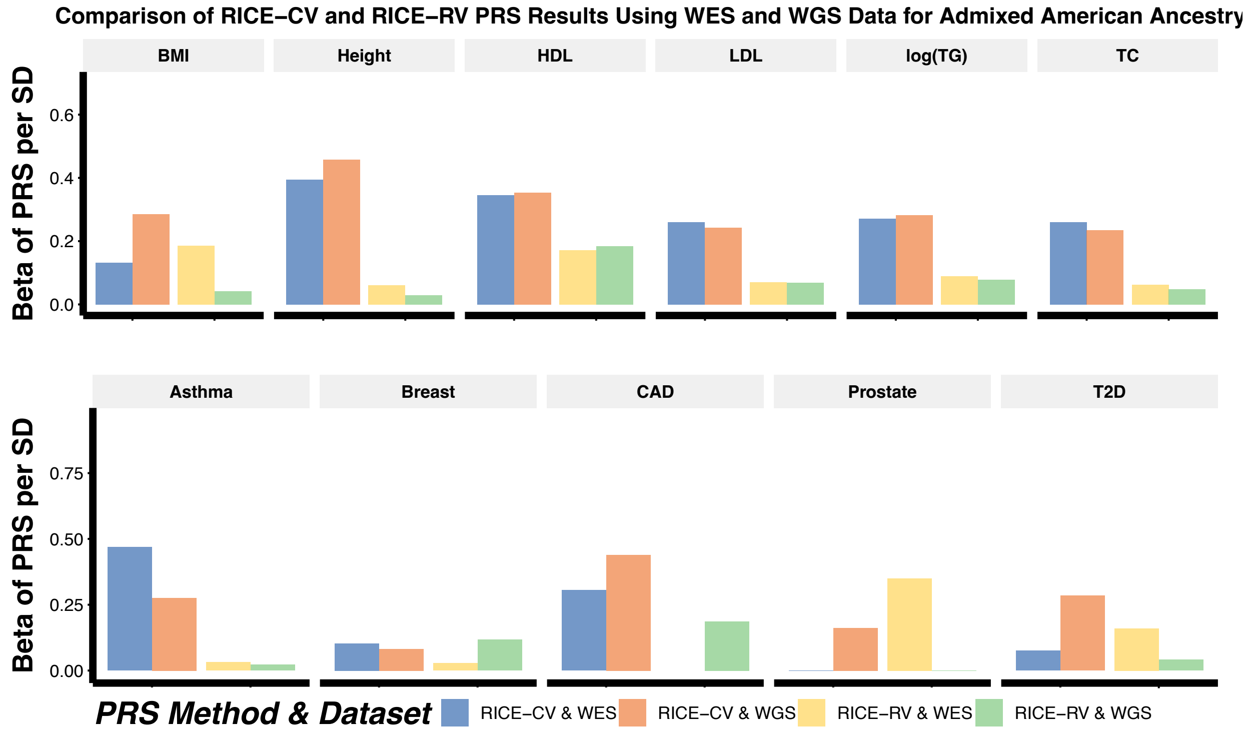


**c)** Comparison of ancestry-adjusted PRSs from RICE-CV and RICE-RV using UKB WES and WGS data for individuals of South Asian ancestry.

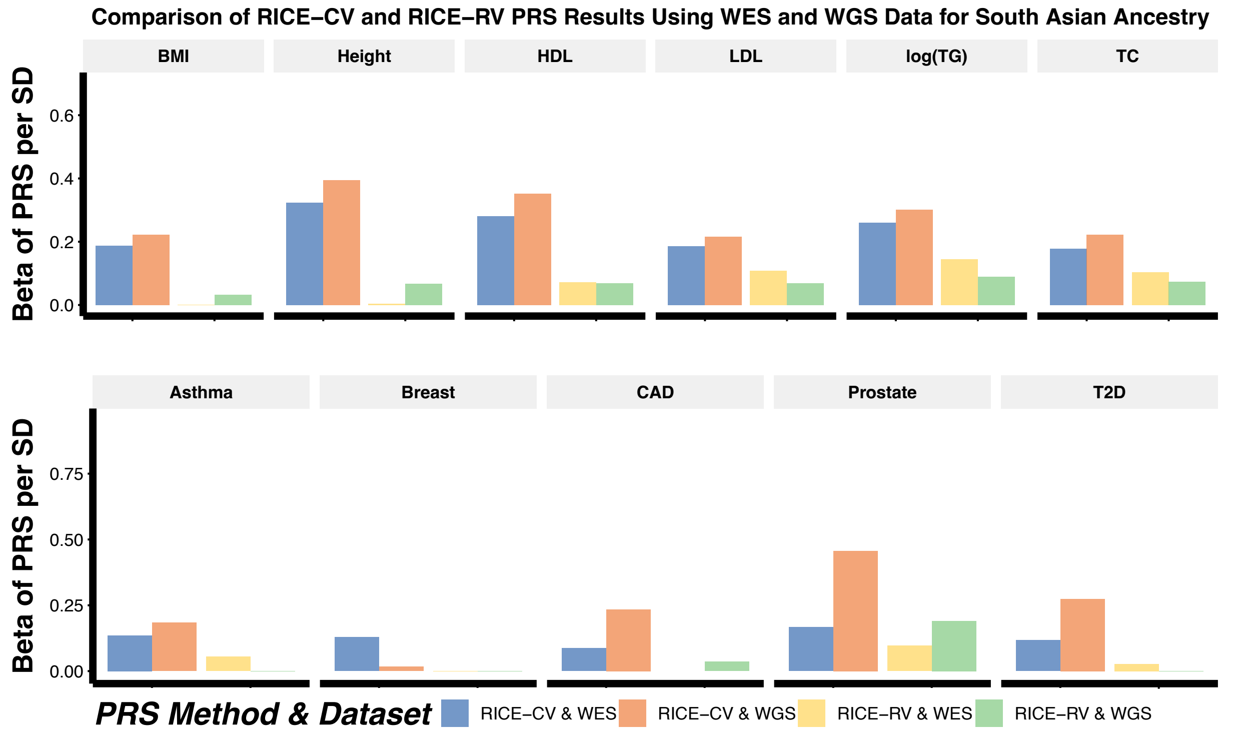

**Supplementary Figure 14. Comparison of ancestry-adjusted PRSs from RICE-CV, RICE-RV, RICE-RV constructed with only coding genes (Coding), and RICE-RV constructed using only noncoding genes (Noncoding) using UK Biobank (UKB) whole-genome sequencing (WGS) data for individuals of either African, Admixed American, or South Asian ancestry.** Results are shown for five binary traits (Supp. Fig. 14a): asthma, breast cancer, coronary artery disease (CAD), prostate cancer, and type 2 diabetes (T2D) and six continuous traits (Supp. Fig. 14b): body mass index (BMI), height, high-density lipoproteins (HDL), low-density lipoproteins (LDL), the natural logarithm of triglyceride (log(TG)), total cholesterol (TC). The training data included only individuals of European ancestry, while the tuning and validation sets contained individuals from all four ancestries. Full sample size details for each ancestry are provided in **Supplementary Tables 2 and 3**.

**a)** Comparison of ancestry-adjusted PRSs from RICE-CV, RICE-RV, RICE-RV constructed with only coding genes (Coding), and RICE-RV constructed using only noncoding genes (Noncoding) using UKB WGS data for five binary traits.

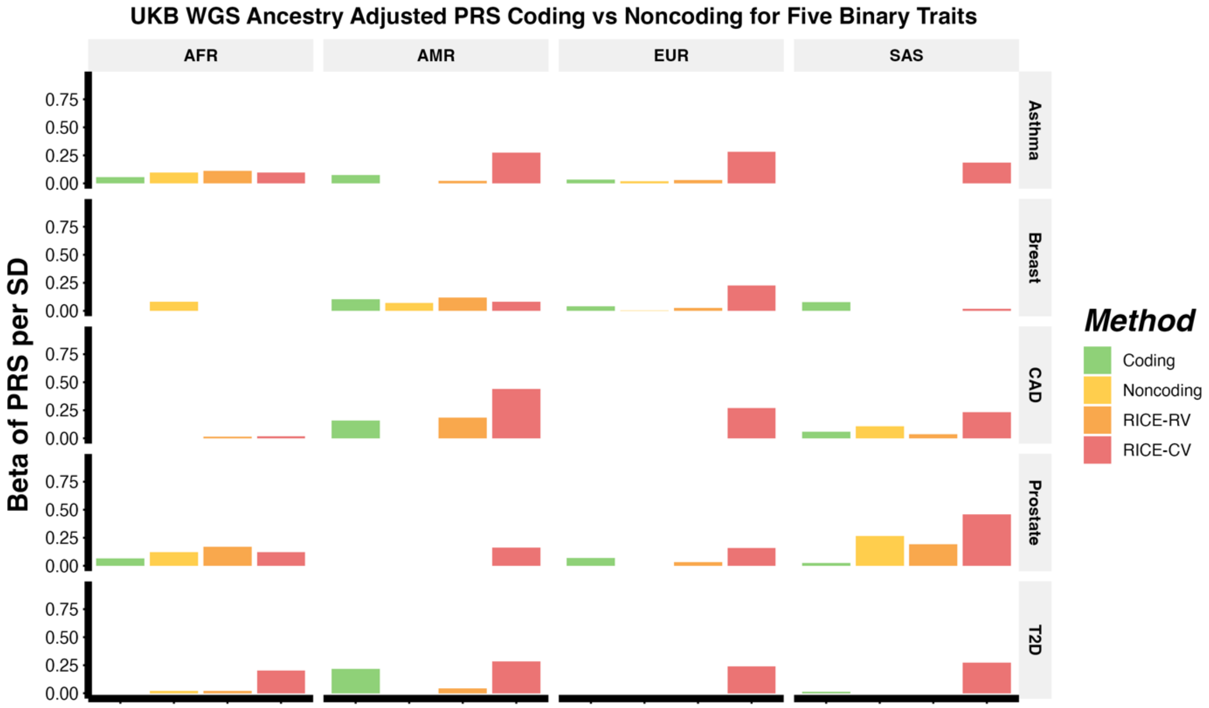

**Supplementary Figure 14 continued. b)** Comparison of ancestry-adjusted PRSs from RICE-CV, RICE-RV, RICE-RV constructed with only coding genes (Coding), and RICE-RV constructed using only noncoding genes (Noncoding) using UKB WGS data for six continuous traits.

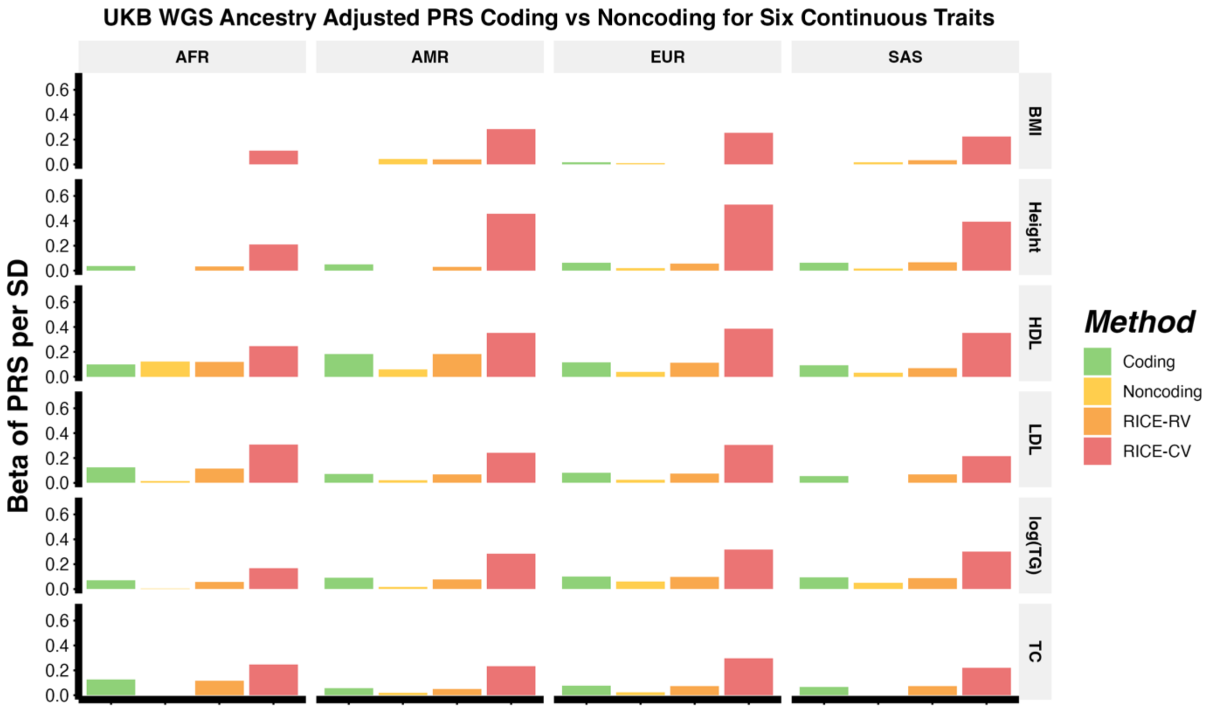

**Supplementary Figure 15.** Manhattan plot and QQ plots based on the UK Biobank whole genome sequencing (WGS) GWAS summary statistics computed using the training set consisting of only individuals of European ancestry (EUR) for five binary traits: asthma, breast cancer, coronary artery disease (CAD), prostate cancer, and type 2 diabetes (T2D). The red and blue shaded regions around the diagonal line in the QQ plots indicate the 95% confidence intervals expected under the null hypothesis of no association between genetic variants and the trait of interest, for minor allele frequencies (MAF) within the ranges (0.05, 0.5] and [0.01, 0.05], respectively. Under the null hypothesis, the p-value follows a uniform (0,1) distribution. The jth order statistic follows a Beta (j, N-j+1) distribution, where N is the total number of variants given a specific MAF cutoff. For binary traits, $\lambda_{1000}$ scales $\lambda$ to a study with 1000 cases and 1000 controls using $\lambda_{1000} = 1 + 1000 \times (\lambda - 1) \times \left(\frac{1}{N_{case}} + \frac{1}{N_{control}}\right)$. Genomic control factors are shown in **Supplementary Table 5**.

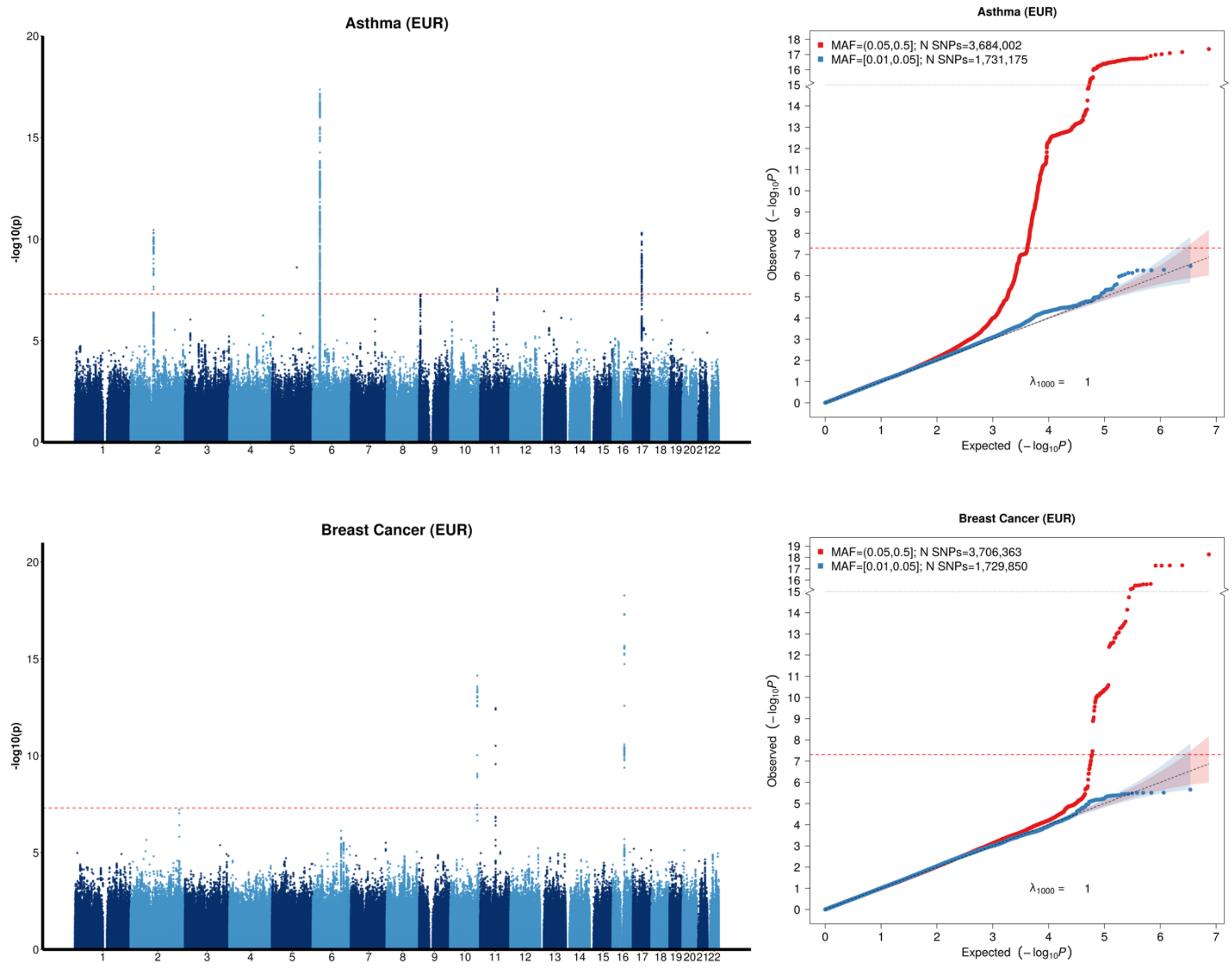

**Supplementary Figure 15 continued.** Manhattan and QQ Plots for CAD, prostate cancer and T2D based on UK Biobank WGS GWAS summary statistics in European populations.

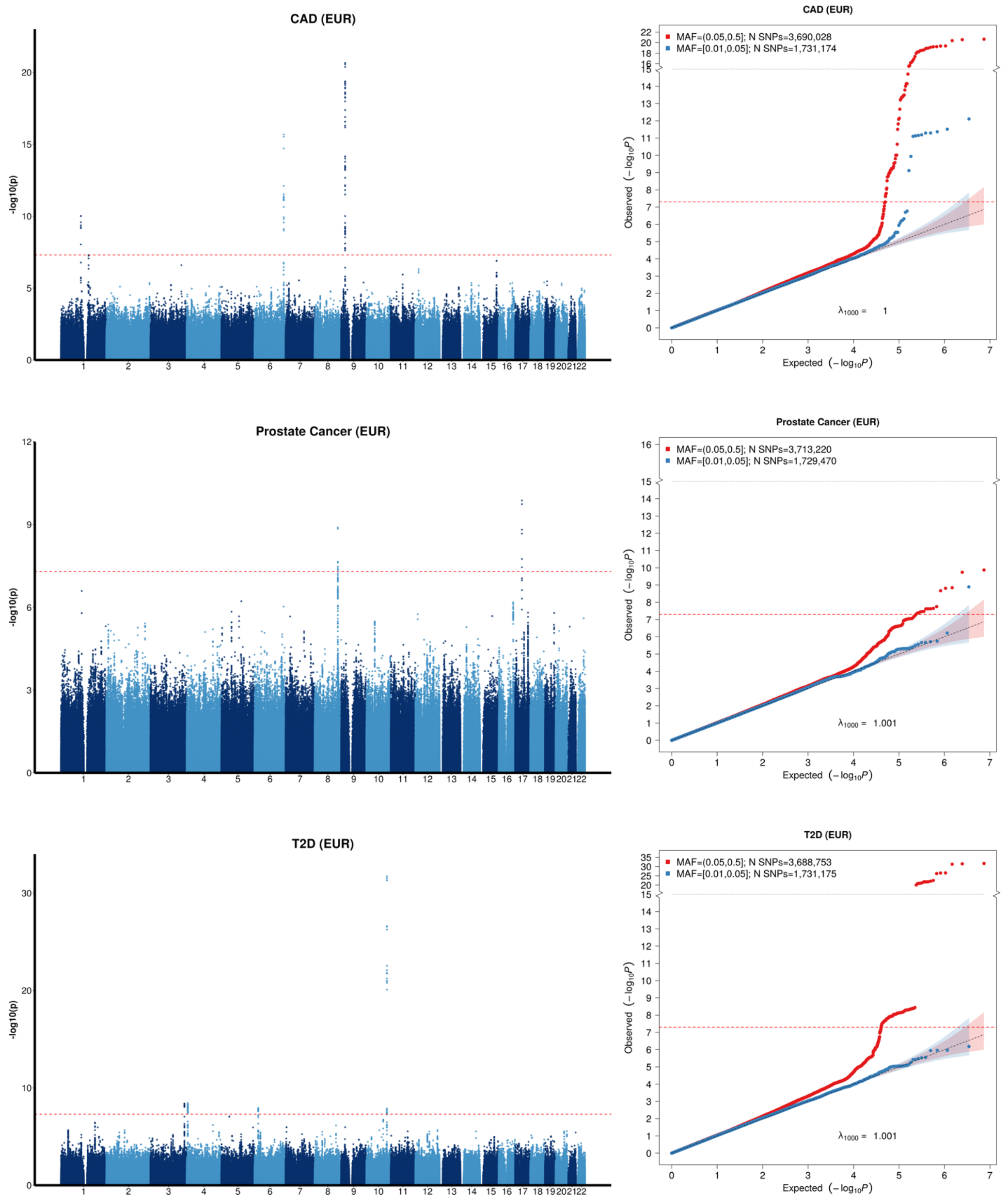

**Supplementary Figure 16.** Manhattan plot and QQ plots based on the UK Biobank whole genome sequencing (WGS) GWAS summary statistics computed using the training set consisting of only individuals of European ancestry (EUR) for six continuous traits: body mass index (BMI), high-density lipoprotein cholesterol (HDL), height, low-density lipoprotein cholesterol (LDL), natural logarithm of triglycerides (log(TG)), and total cholesterol (TC). The red and blue shaded regions around the diagonal line in the QQ plots indicate the 95% confidence intervals expected under the null hypothesis of no association between genetic markers and the trait of interest, for minor allele frequencies (MAF) within the ranges (0.05, 0.5] and [0.01, 0.05], respectively. Under the null hypothesis, the p-value follows a uniform (0,1) distribution. The jth order statistic follows a Beta (j, N-j+1) distribution, where N is the total number of variants given a specific MAF cutoff. For continuous traits, $\lambda_{1000}$ scales $\lambda$ to a study with 1000 cases and 1000 controls using $\lambda_{1000} = 1 + 1000 \times (\lambda - 1)/\mathrm{N}$. Genomic control factors are shown in **Supplementary Table 5**.

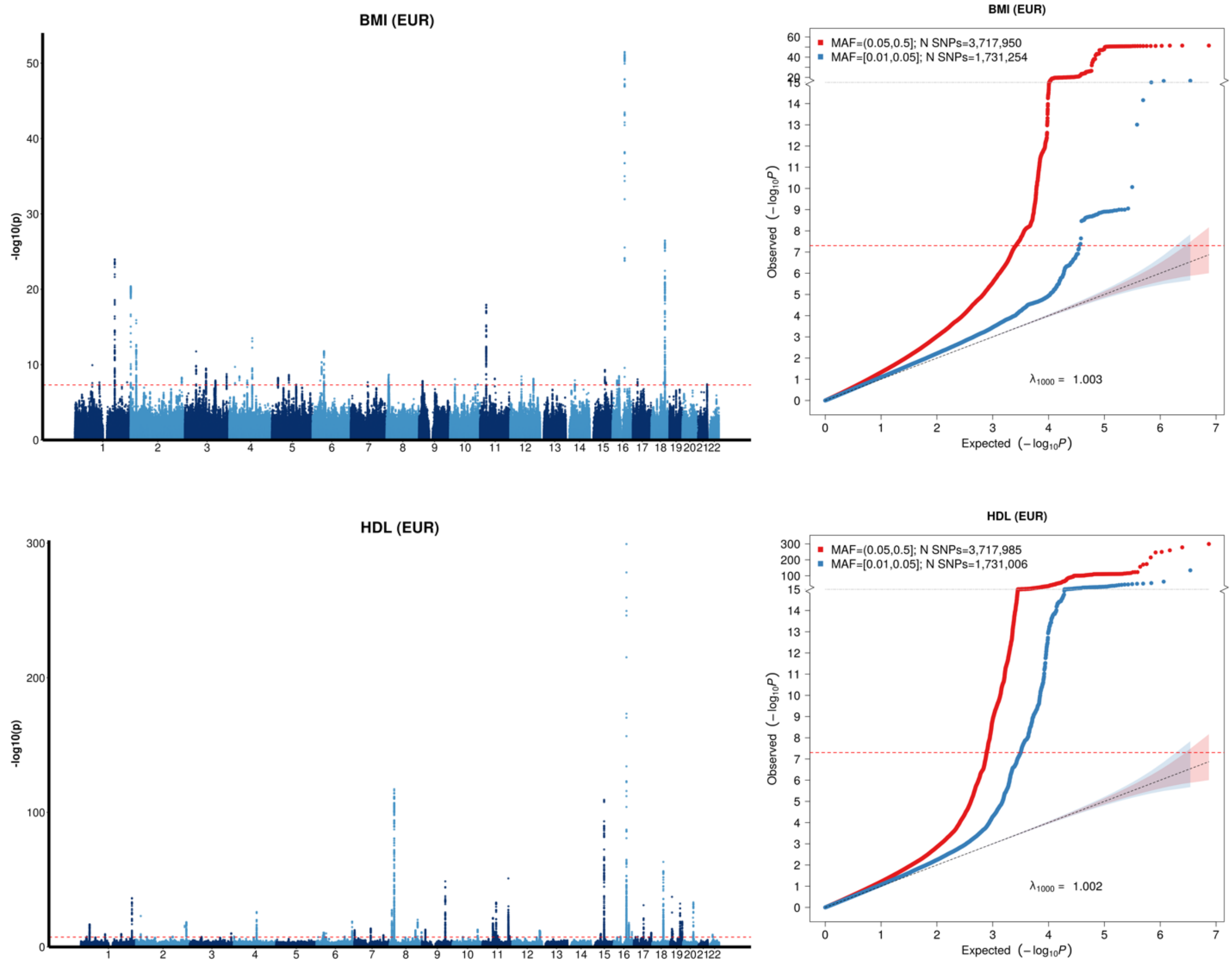

**Supplementary Figure 16 continued.** Manhattan and QQ Plots for height, LDL and log(TG) based on UK Biobank WGS GWAS summary statistics in European populations.

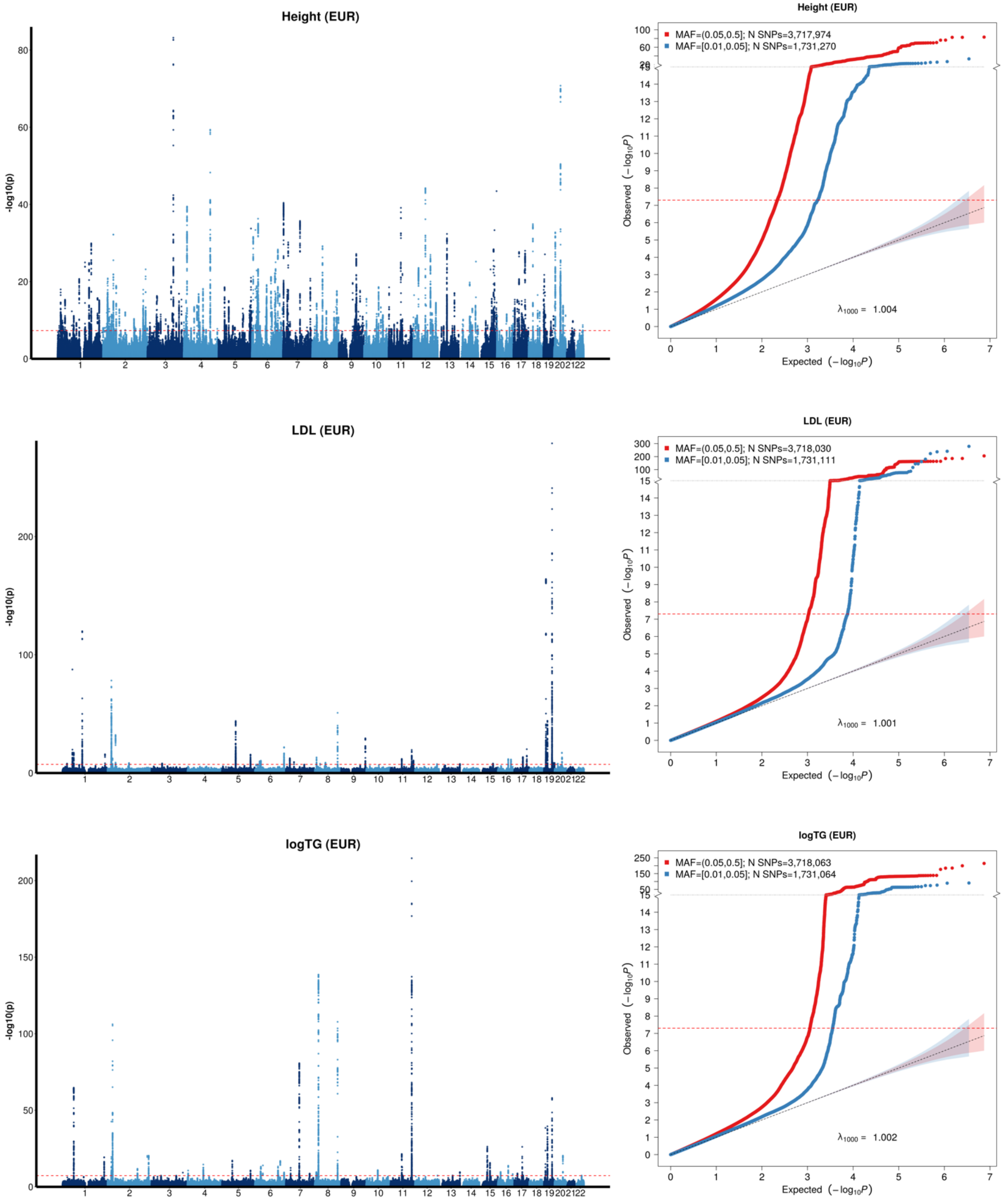

**Supplementary Figure 16 continued.** Manhattan and QQ Plots for TC based on UK Biobank WGS GWAS summary statistics in European populations.

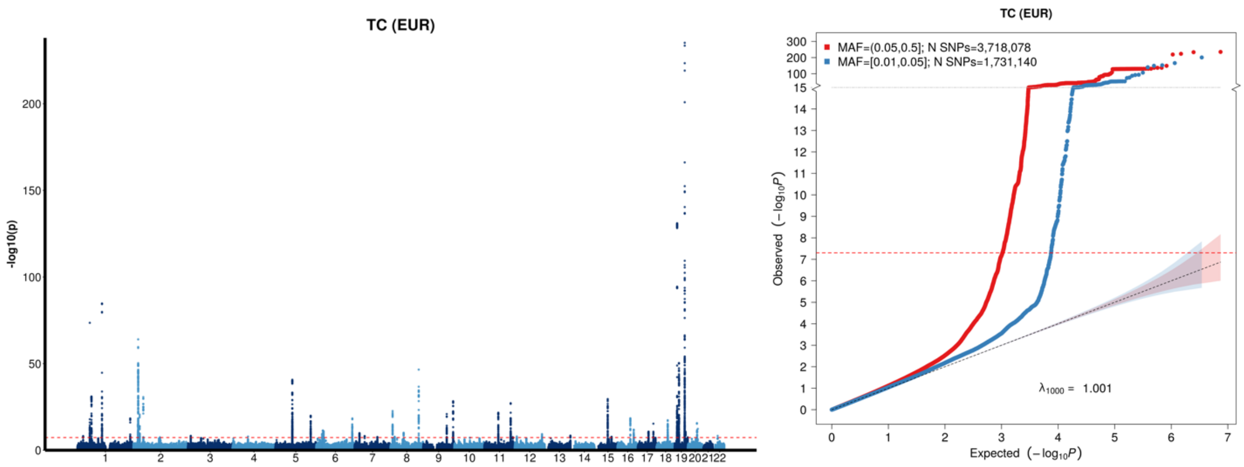

**Supplementary Figure 17.** QQ plots based on the UK Biobank whole genome sequencing rare variant association analysis for coding genes conducted with STAARpipeline using the training set consisting of only individuals of European ancestry (EUR) for 11 traits (sample sizes provided in **Supplementary Table 3**). The five binary traits analyzed and displayed in Supplementary Figure 17a: asthma, breast cancer, coronary artery disease (CAD), prostate cancer, and type 2 diabetes (T2D) and the six continuous traits analyzed and displayed in Supplementary Figure 17b: body mass index (BMI), high-density lipoprotein cholesterol (HDL), height, low-density lipoprotein cholesterol (LDL), natural logarithm of triglycerides (log(TG)), and total cholesterol (TC). P-values are split into the five functional categories investigated in the gene-centric coding analysis in the STAARpipeline: putative loss of function (pLoF), putative loss of function and disruptive (pLoF+D), missense, disruptive missense, and synonymous. Under the null hypothesis, the p-value follows a uniform (0,1) distribution.

**a)** QQ plots for five binary traits from the UKB WGS rare variant association analysis using coding genes with STAARpipeline.

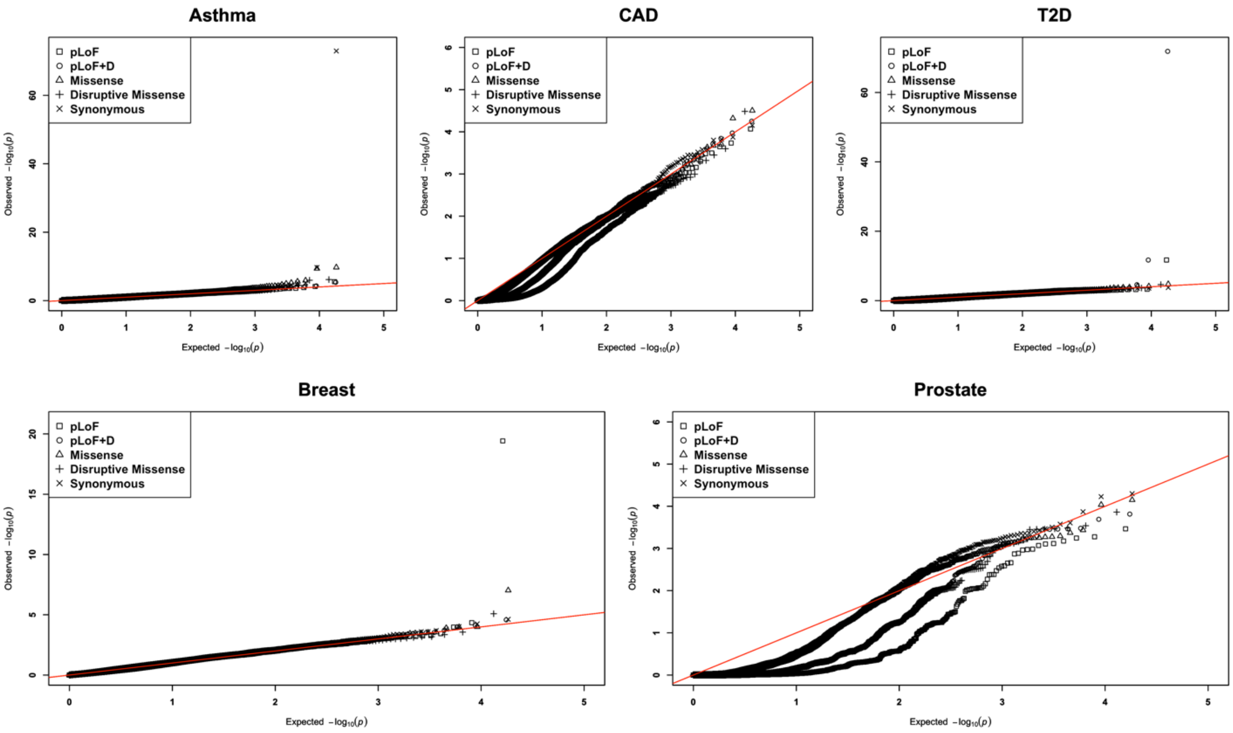

**Supplementary Figure 17 continued. b)** QQ plots for six continuous traits from the UKB WGS rare variant association analysis using coding genes with STAARpipeline.

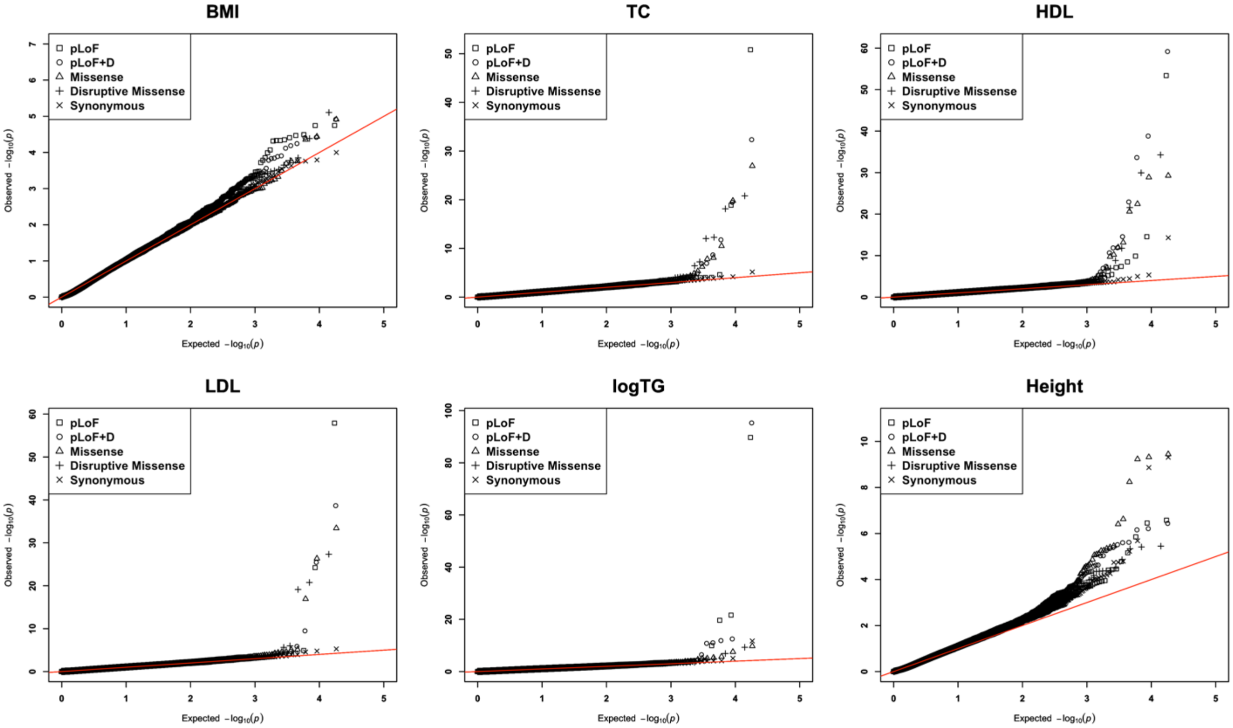

**Supplementary Figure 18.** QQ plots based on the UK Biobank whole genome sequencing rare variant association analysis for noncoding genes conducted with STAARpipeline using the training set consisting of only individuals of European ancestry (EUR) for 11 traits (sample sizes provided in **Supplementary Table 3**). The five binary traits analyzed and displayed in Supplementary Figure 18a: asthma, breast cancer, coronary artery disease (CAD), prostate cancer, and type 2 diabetes (T2D) and the six continuous traits analyzed and displayed in Supplementary Figure 18b: body mass index (BMI), high-density lipoprotein cholesterol (HDL), height, low-density lipoprotein cholesterol (LDL), natural logarithm of triglycerides (log(TG)), and total cholesterol (TC). P-values are split into the eight functional categories investigated in the gene-centric noncoding analysis in the STAARpipeline: upstream, downstream, noncoding RNA (ncRNA), untranslated regions (UTR), promoters within cap-analysis gene expression (promoter-CAGE), promoters within DNase 1 hypersensitive regions (promoter-DHS), enhancers within cap-analysis gene expression (enhancer-CAGE), and enhancers within DNase 1 hypersensitive regions (enhancer-DHS).Under the null hypothesis, the p-value follows a uniform (0,1) distribution.

**a)** QQ plots for five binary traits from the UKB WGS rare variant association analysis using noncoding genes with STAARpipeline.

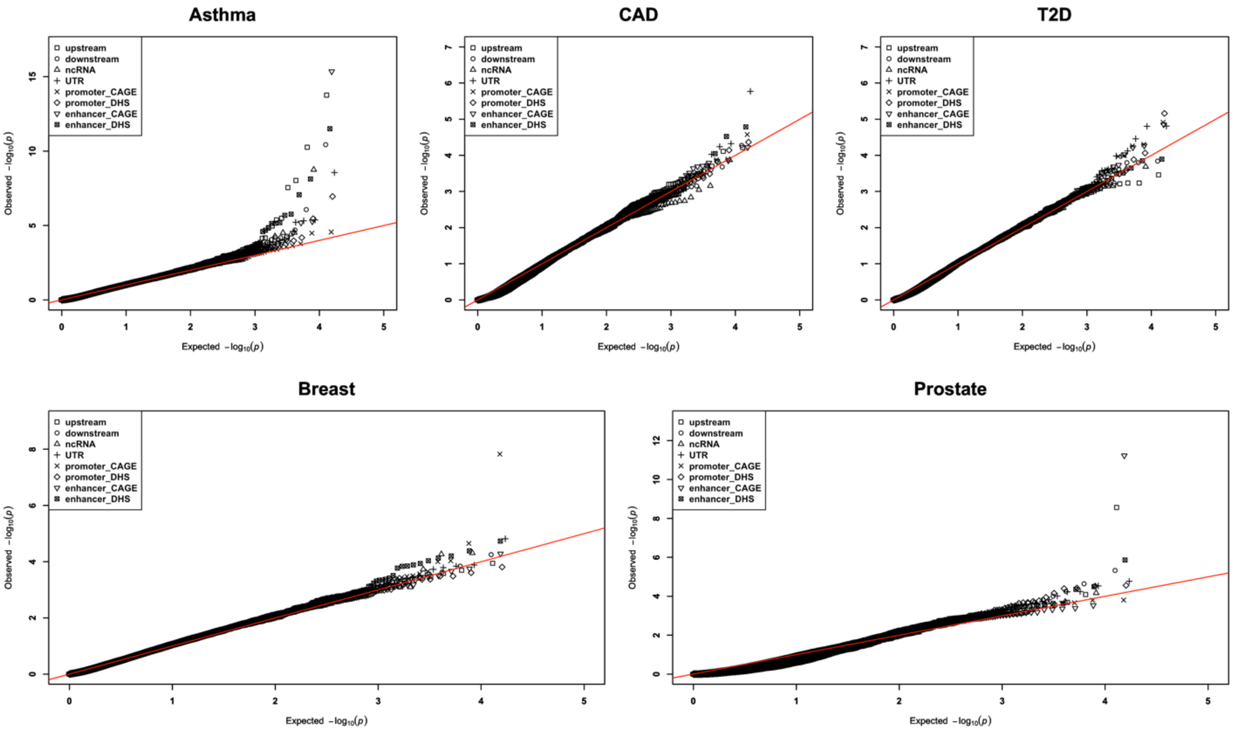

**Supplementary Figure 18 continued. b)** QQ plots for six continuous traits from the UKB WGS rare variant association analysis using noncoding genes with STAARpipeline.

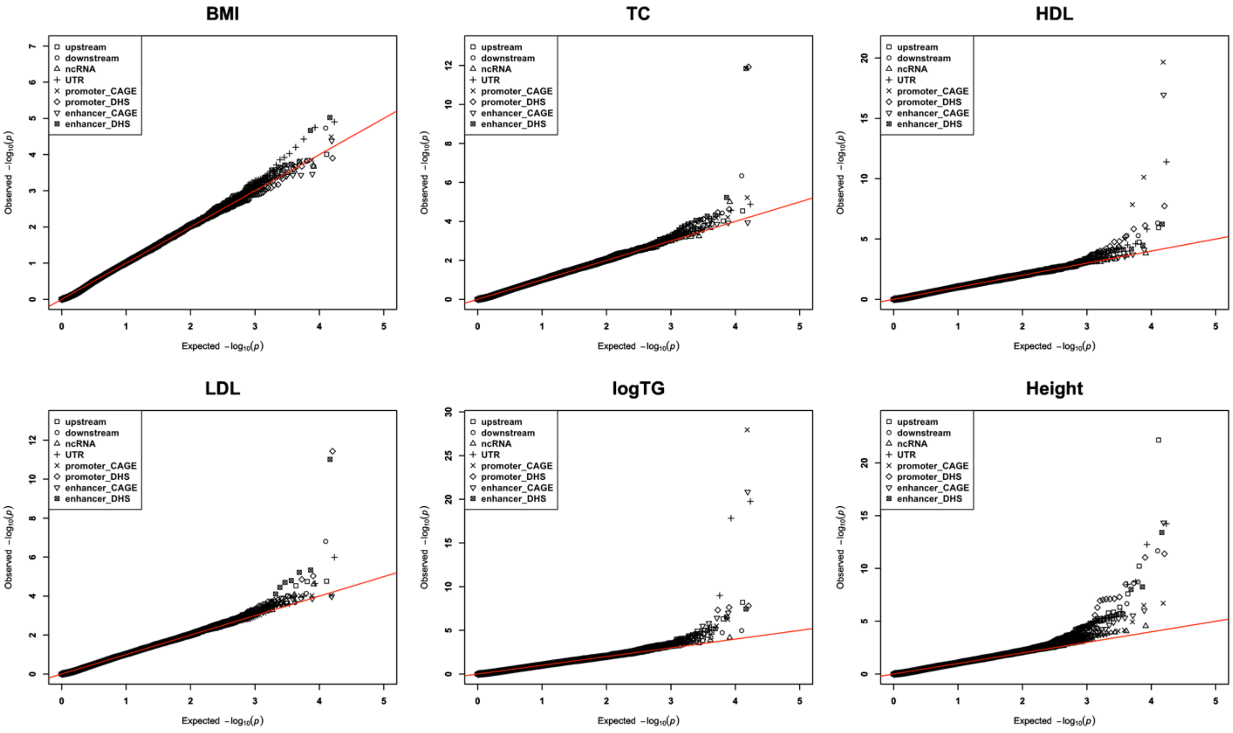

**Supplementary Figure 19. Relationship between common and rare variant PRSs and standardized traits across six ancestral groups from All of Us (AoU)) data.** The six continuous traits analyzed and shown: body mass index (BMI) (Supp. Fig. 19a), height (Supp. Fig. 19b), low-density lipoprotein cholesterol (LDL) (Supp. Fig. 19c), natural logarithm of triglyceride cholesterol (log(TG)) (Supp. Fig. 19d), and total cholesterol (TC) (Supp. Fig. 19e). PRS quantiles for RICE-CV (common variants) are plotted on the x-axis, and standardized trait on the y-axis. Data are stratified by rare variant PRS quantiles from RICE-RV (red: below 5%, green: 20–70%, blue: above 95%). Results are shown for individuals of African (AFR), Admixed American/Latino (AMR), East Asian (EAS), European (EUR), Middle Eastern (MID), and South Asian (SAS) ancestries. The training data consisted of individuals of African, Admixed American, and European ancestries, while the tuning and validation sets included all six ancestries. Full sample sizes details for each ancestry are provided in **Supplementary Table 4.**

**a)** Relationship between ancestry-adjusted common and rare variant PRSs and standardized body mass index (BMI) levels across six ancestral groups from AoU data.

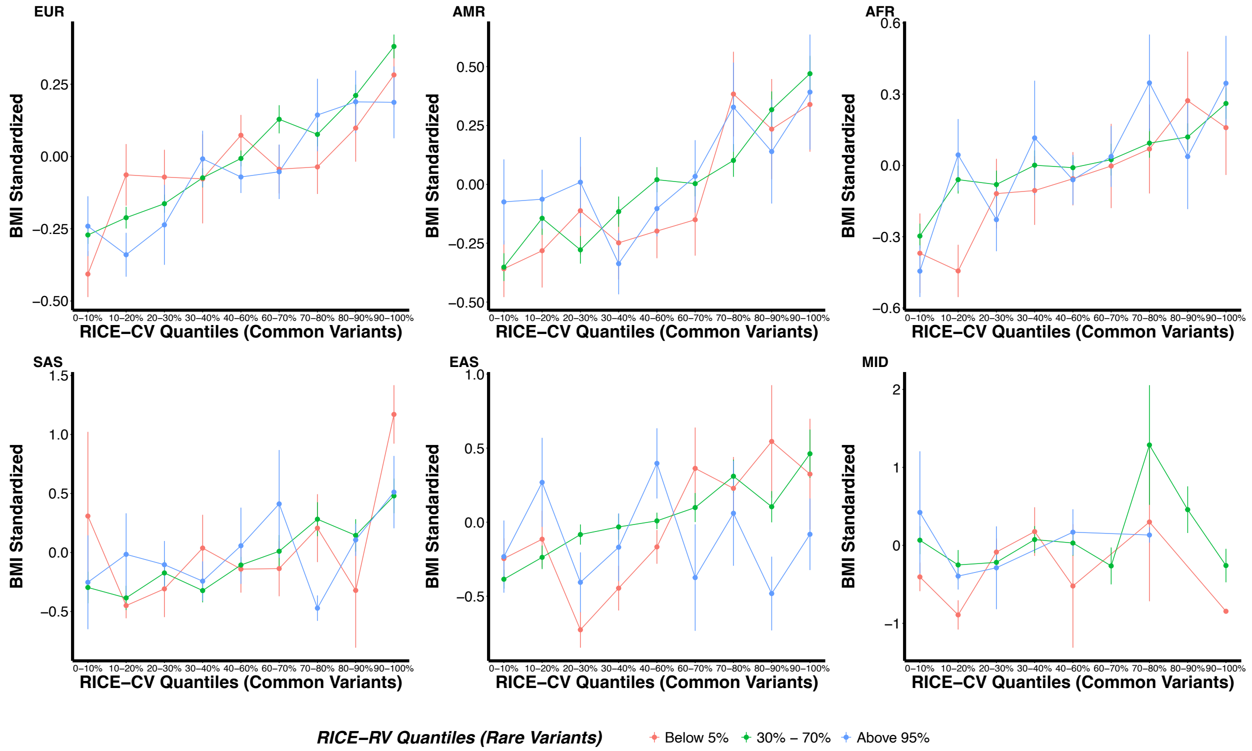

**Supplementary Figure 19 continued. b)** Relationship between ancestry-adjusted common and rare variant PRSs and standardized high-density lipoprotein cholesterol (HDL) levels across six ancestral groups from AoU data.

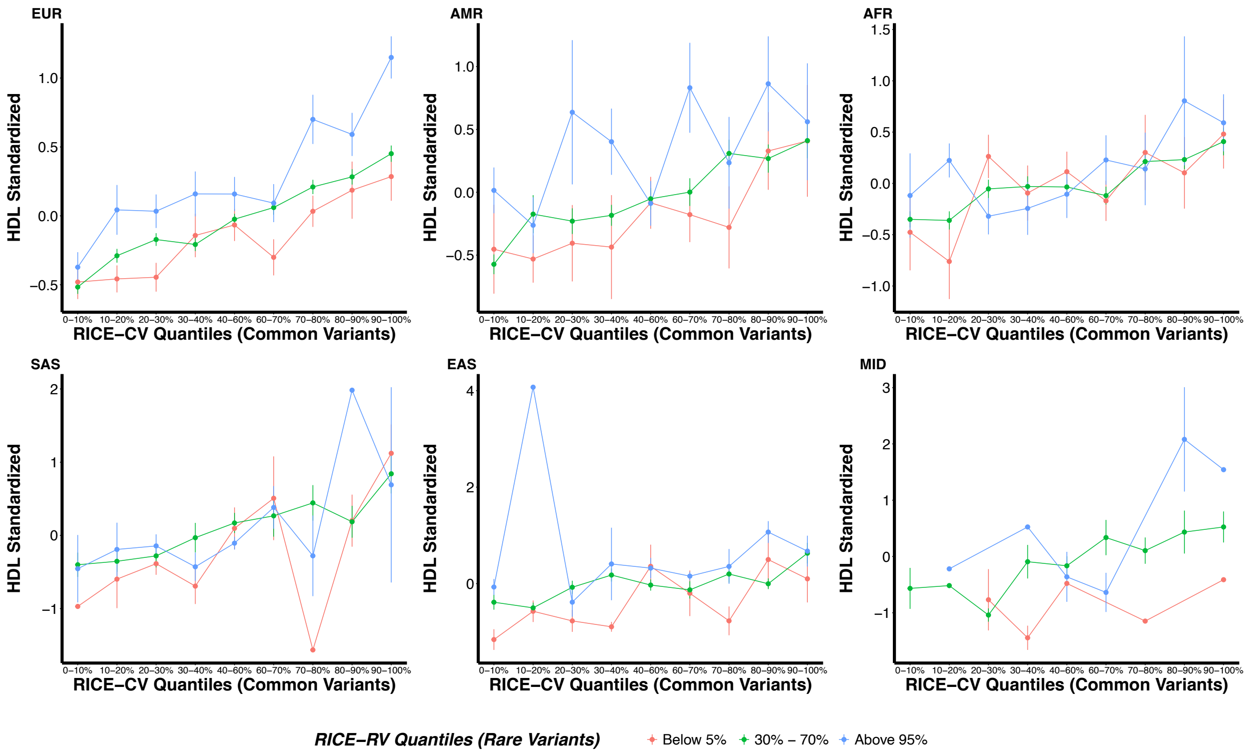

**Supplementary Figure 19 continued. c)** Relationship between ancestry-adjusted common and rare variant PRSs and standardized height across six ancestral groups from AoU data.

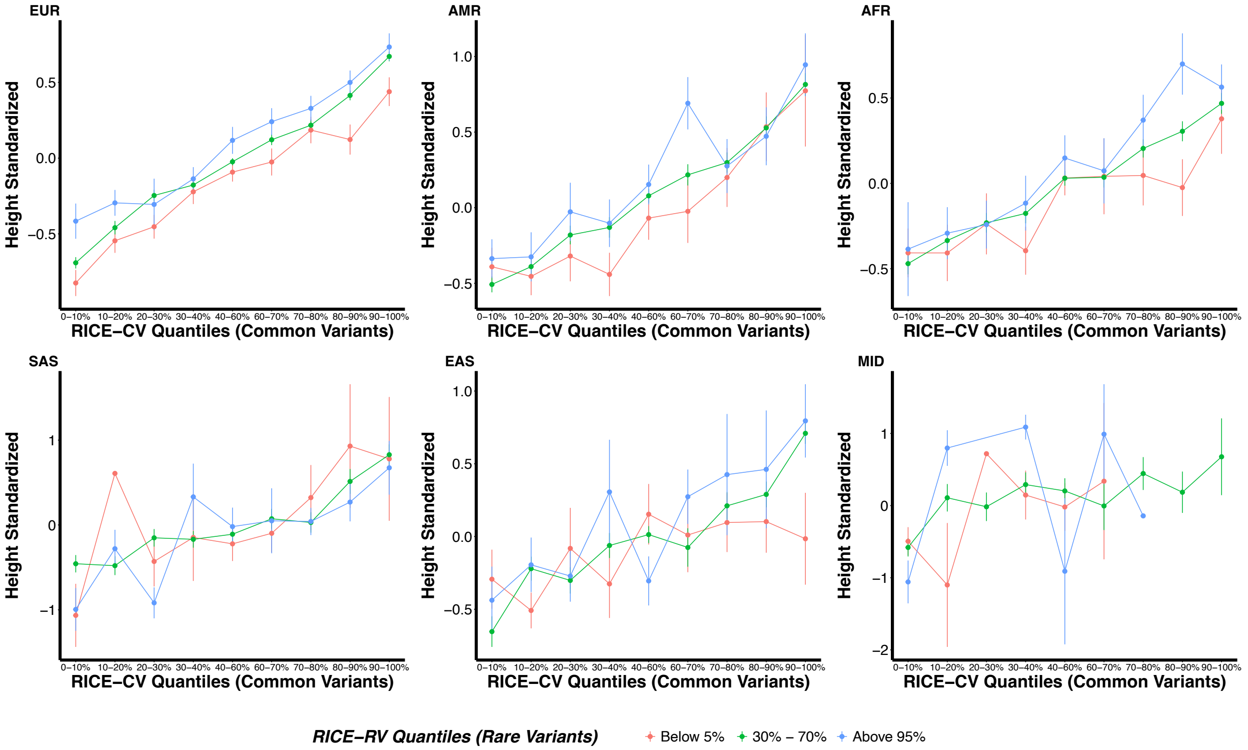

**Supplementary Figure 19 continued. d)** Relationship between ancestry-adjusted common and rare variant PRSs and standardized low-density lipoprotein cholesterol (LDL) levels across six ancestral groups from AoU data.

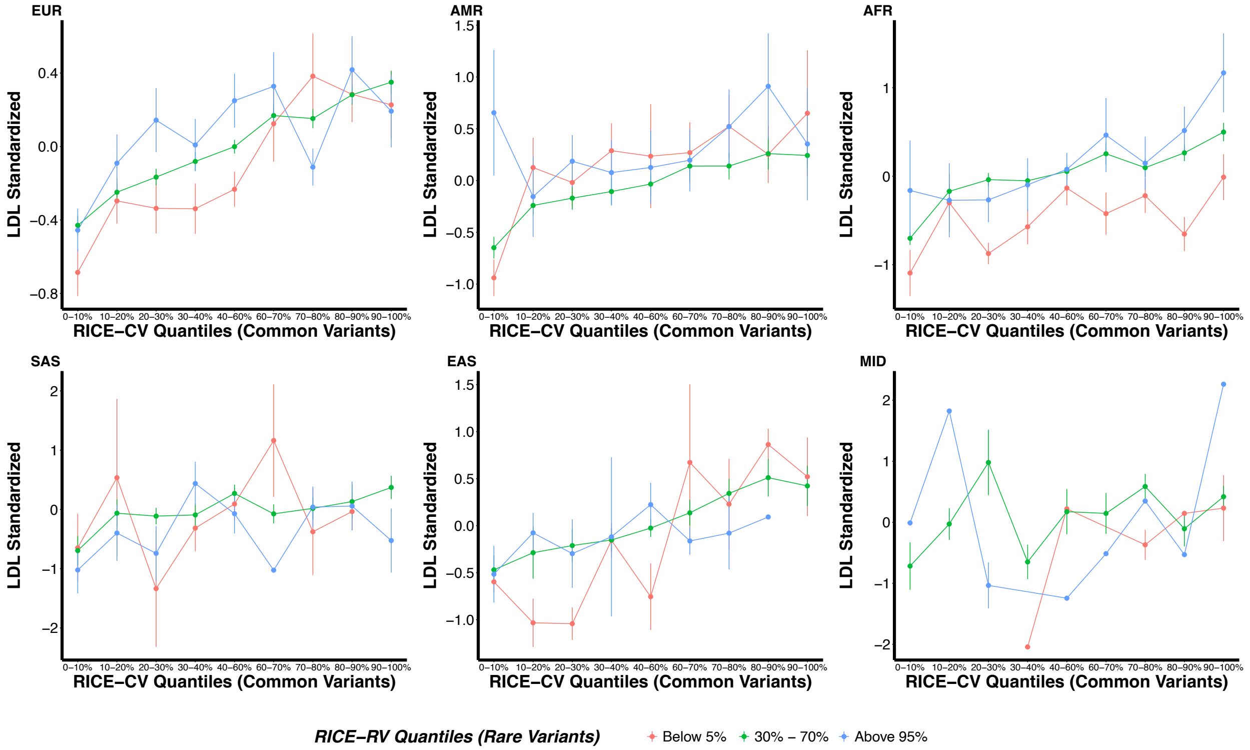

**Supplementary Figure 19 continued. e)** Relationship between ancestry-adjusted common and rare variant PRSs and standardized natural logarithm of triglycerides (log(TG)) levels across six ancestral groups from AoU data.

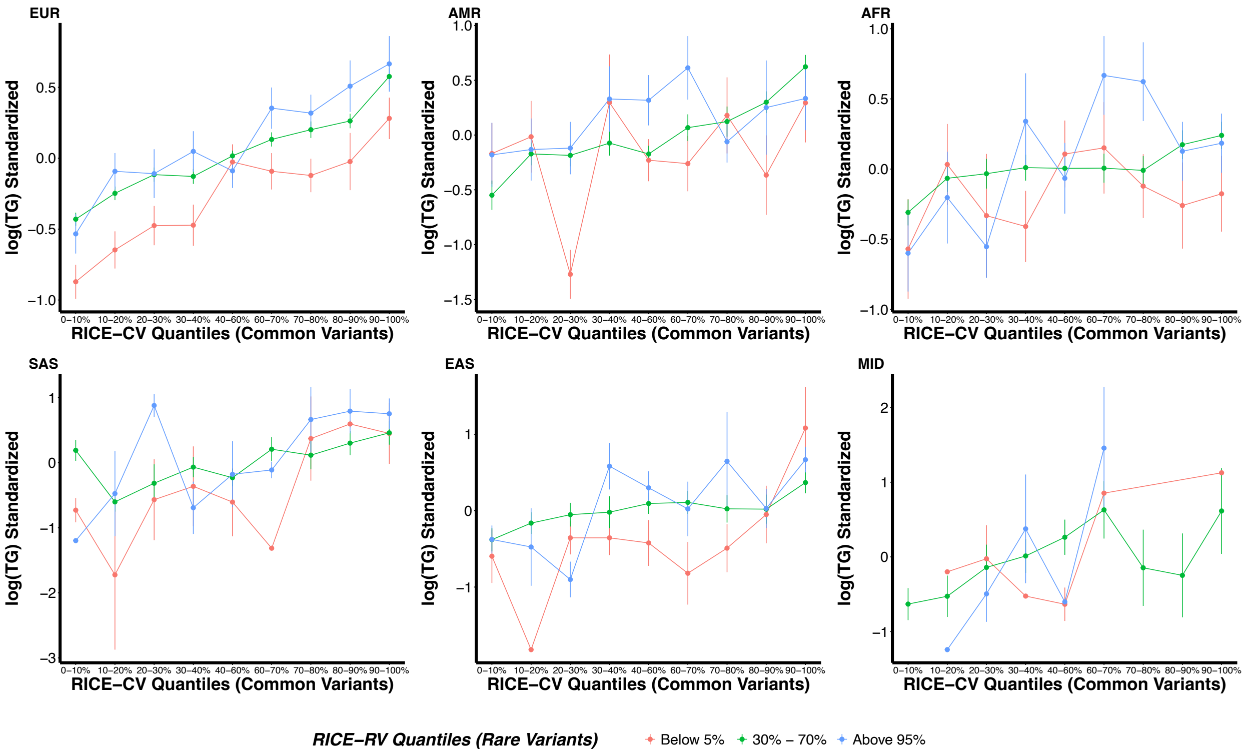

**Supplementary Figure 19 continued. f)** Relationship between ancestry-adjusted common and rare variant PRSs and standardized total cholesterol (TC) levels across six ancestral groups from AoU data.

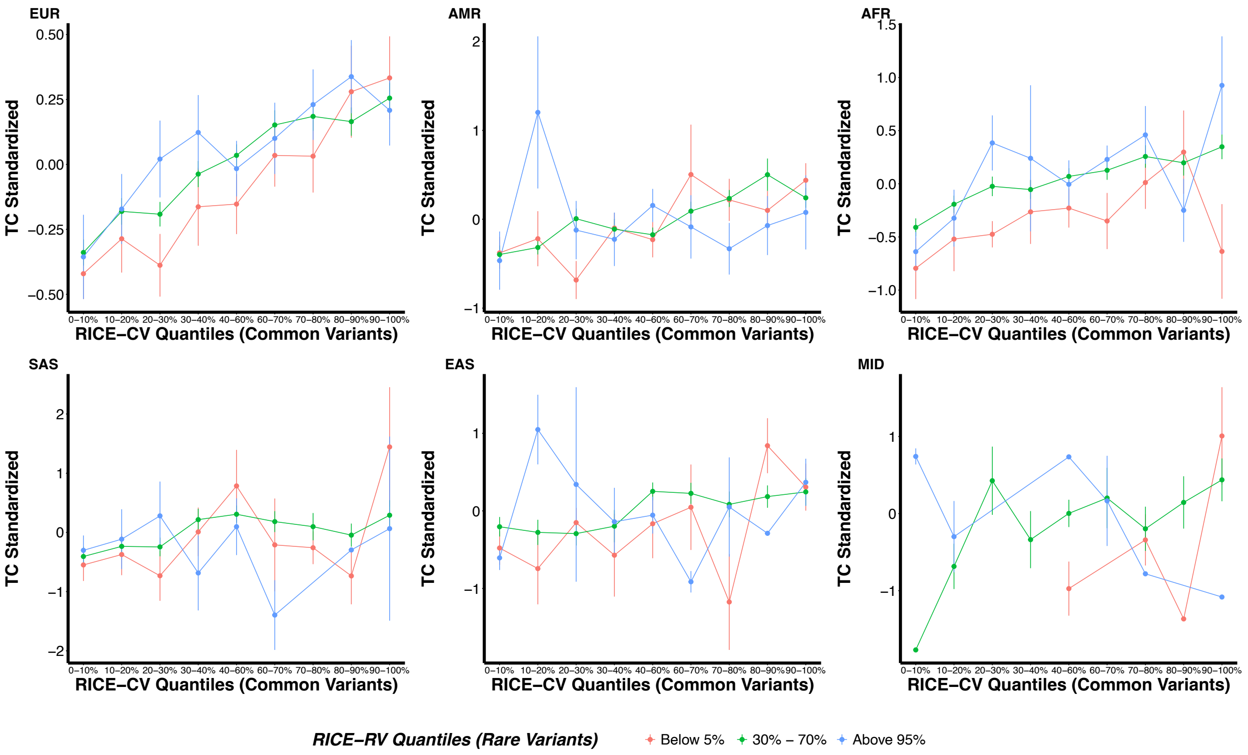

**Supplementary Figure 20.** Manhattan plot and QQ plots based on the All of Us (AoU) GWAS summary statistics computed using the training set consisting of individuals of either African (AFR), Admixed American (AMR), or European (EUR) ancestries for six continuous traits: body mass index (BMI), high-density lipoprotein cholesterol (HDL), height, low-density lipoprotein cholesterol (LDL), natural logarithm of triglycerides (log(TG)), and total cholesterol (TC). The red and blue shaded regions around the diagonal line in the QQ plots indicate the 95% confidence intervals expected under the null hypothesis of no association between genetic markers and the trait of interest, for minor allele frequencies (MAF) within the ranges (0.05, 0.5] and [0.01, 0.05], respectively. Under the null hypothesis, the p-value follows a uniform (0,1) distribution. The jth order statistic follows a Beta (j, N-j+1) distribution, where N is the total number of variants given a specific MAF cutoff. For continuous traits, $\lambda_{1000}$ scales $\lambda$ to a study with 1000 cases and 1000 controls using $\lambda_{1000} = 1 + 1000 \times (\lambda - 1)/N$. Genomic control factors are shown in **Supplementary Table 5**.

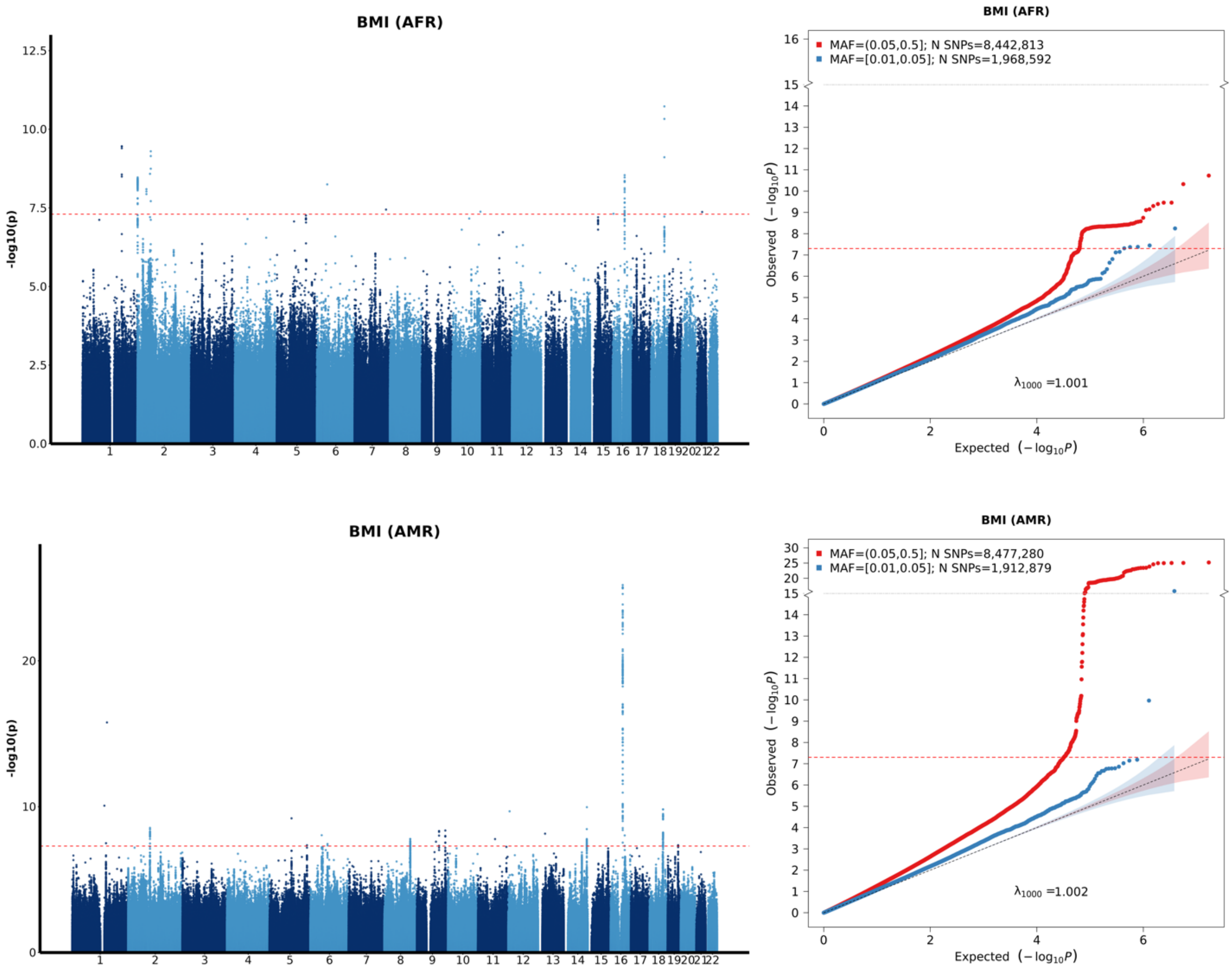

**Supplementary Figure 20 continued.** Manhattan and QQ Plots for BMI (EUR), HDL (AFR, AMR) based on All of Us GWAS summary statistics.

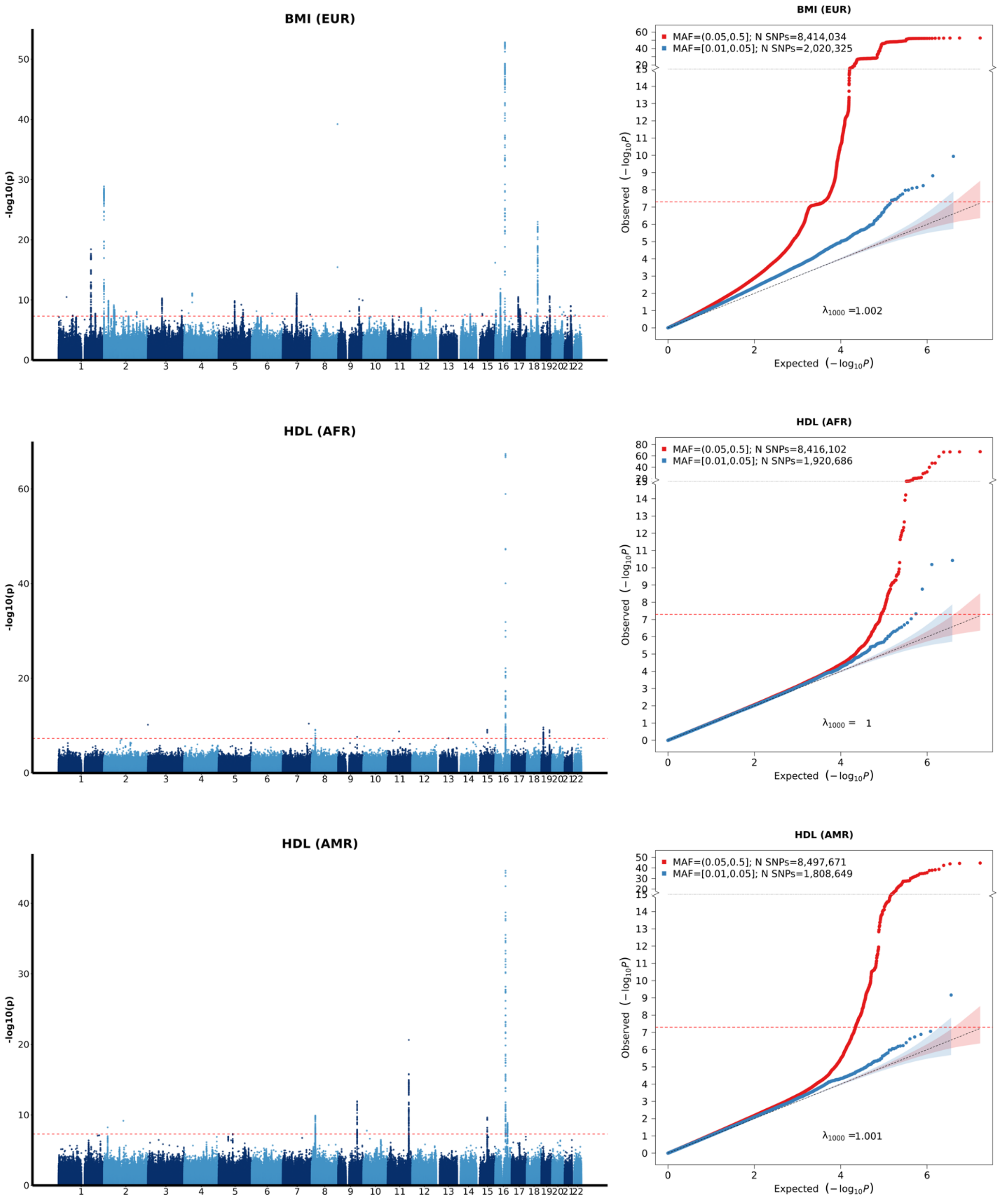

**Supplementary Figure 20 continued.** Manhattan and QQ Plots for HDL (EUR), height (AFR, AMR) based on All of Us GWAS summary statistics.

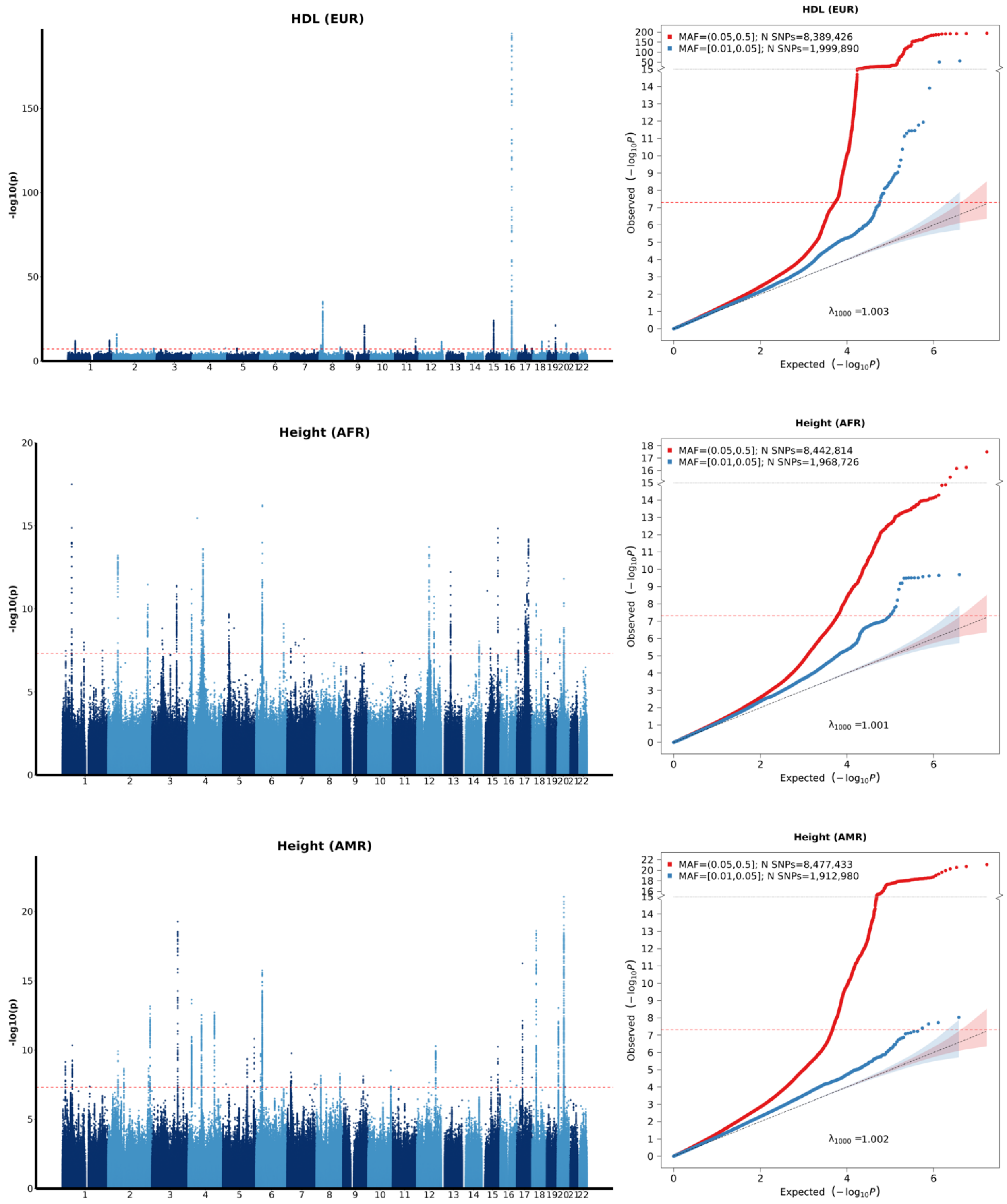

**Supplementary Figure 20 continued.** Manhattan and QQ Plots for height (EUR), LDL (AFR, AMR) based on All of Us GWAS summary statistics.

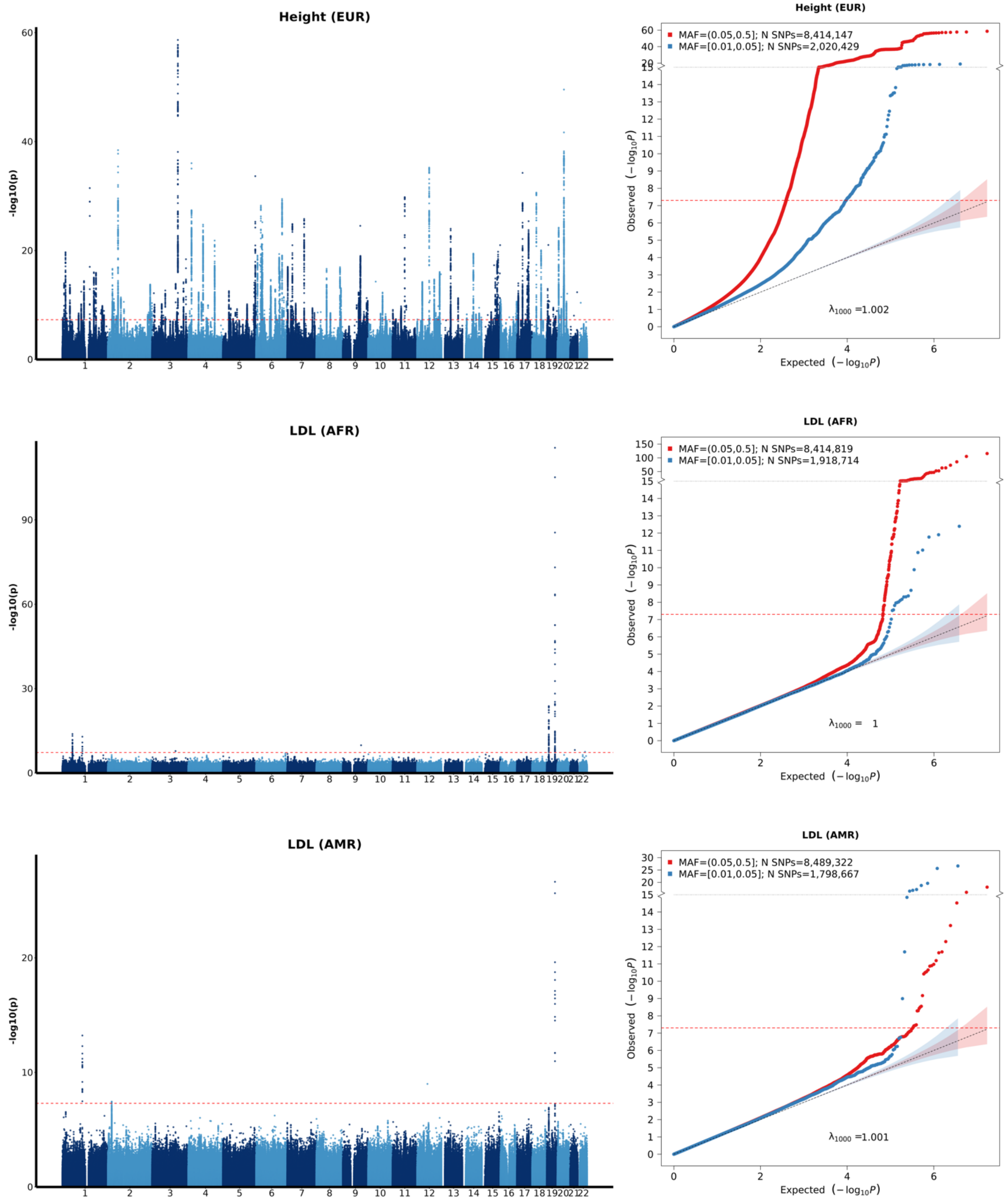

**Supplementary Figure 20 continued.** Manhattan and QQ Plots for LDL (EUR), log(TG) (AFR, AMR) based on All of Us GWAS summary statistics.

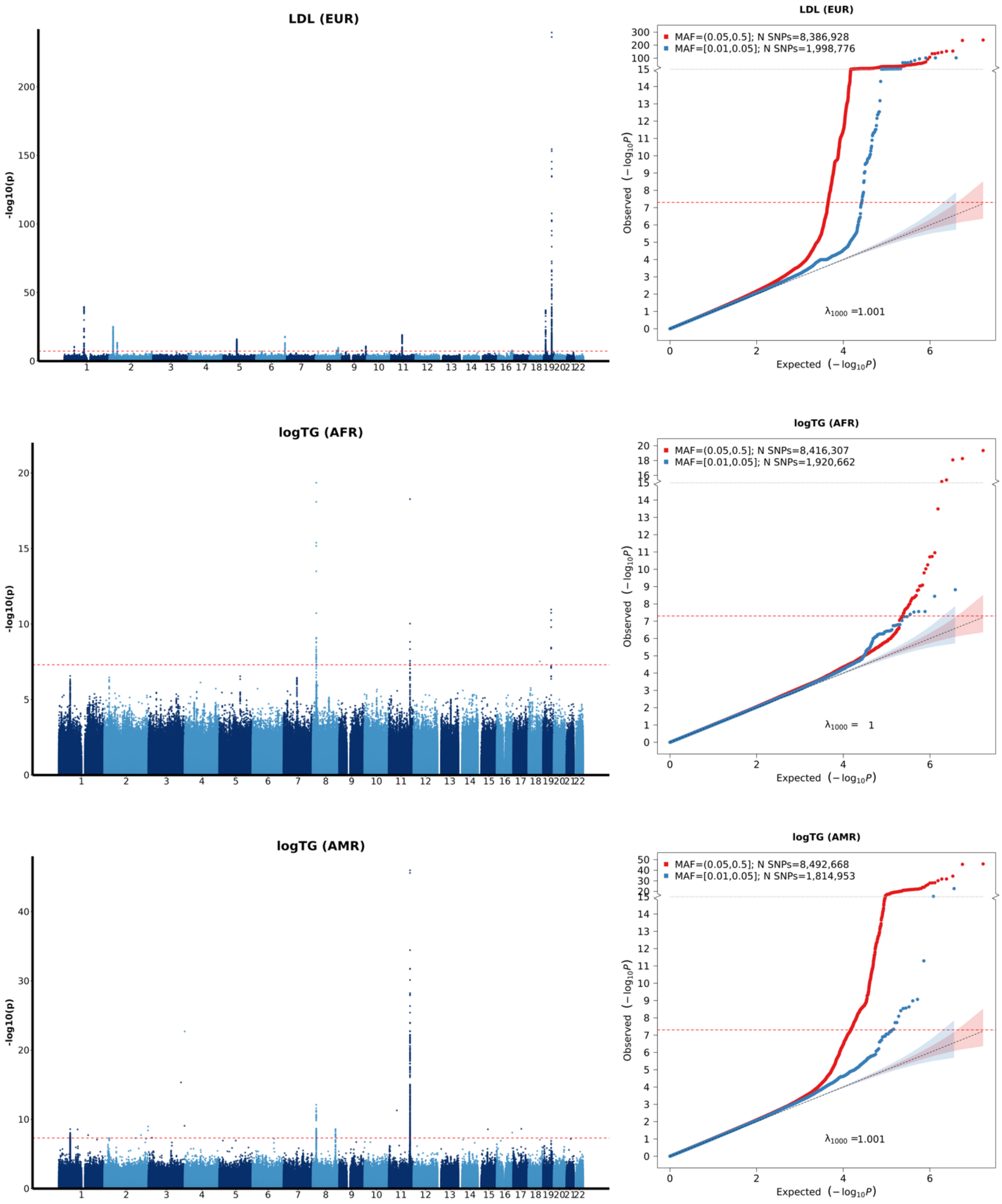

**Supplementary Figure 20 continued.** Manhattan and QQ Plots for log(TG) (EUR), TC (AFR, AMR) based on All of Us GWAS summary statistics.

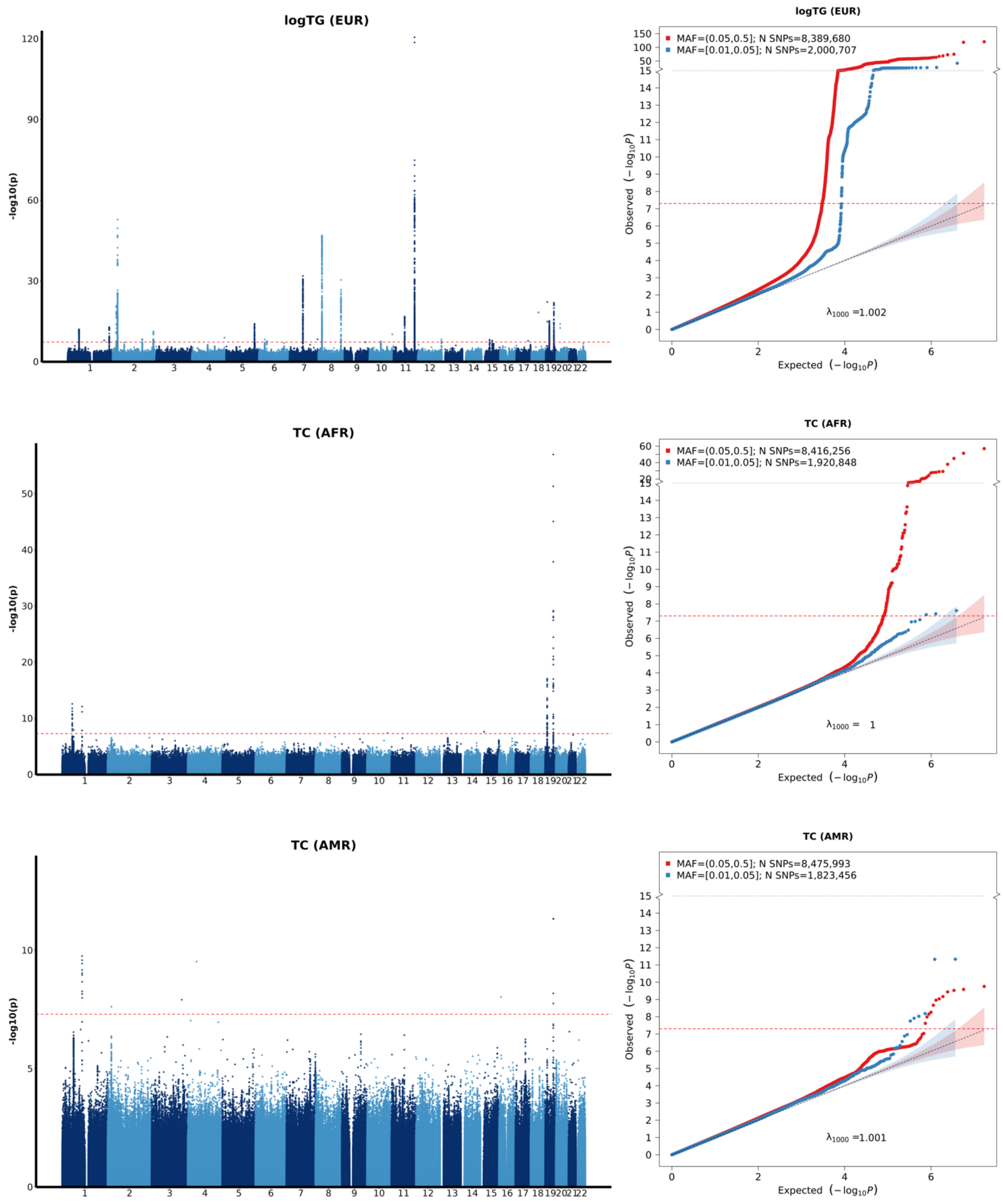

**Supplementary Figure 20 continued.** Manhattan and QQ Plots for TC (EUR) based on All of Us GWAS summary statistics.

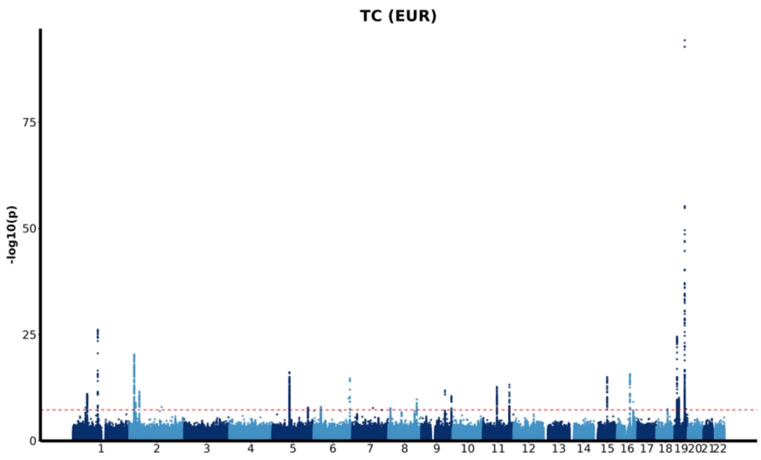


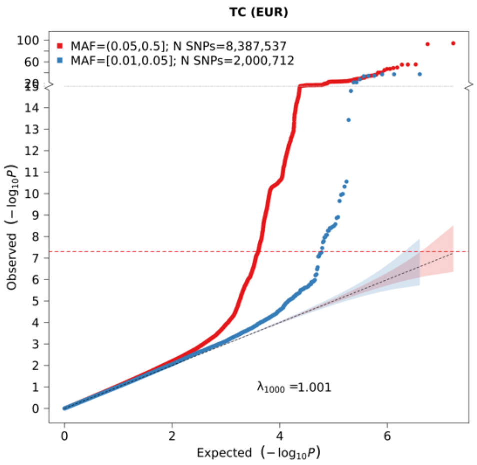

**Supplementary Figure 21.** QQ plots based on the All of Us rare variant association analysis for coding genes conducted with STAARpipeline using the training set consisting of only individuals of European ancestry (EUR) for training set consisting of individuals of either African (AFR), Admixed American (AMR), or European (EUR) ancestry for six continuous traits: body mass index (BMI), high-density lipoprotein cholesterol (HDL), height, low-density lipoprotein cholesterol (LDL), natural logarithm of triglycerides (log(TG)), and total cholesterol (TC) (sample sizes provided in **Supplementary Table 3**). P-values are split into the five functional categories investigated in the gene-centric coding analysis in the STAARpipeline: putative loss of function (pLoF), putative loss of function and disruptive (pLoF+D), missense, disruptive missense, and synonymous. Under the null hypothesis, the p-value follows a uniform (0,1) distribution.

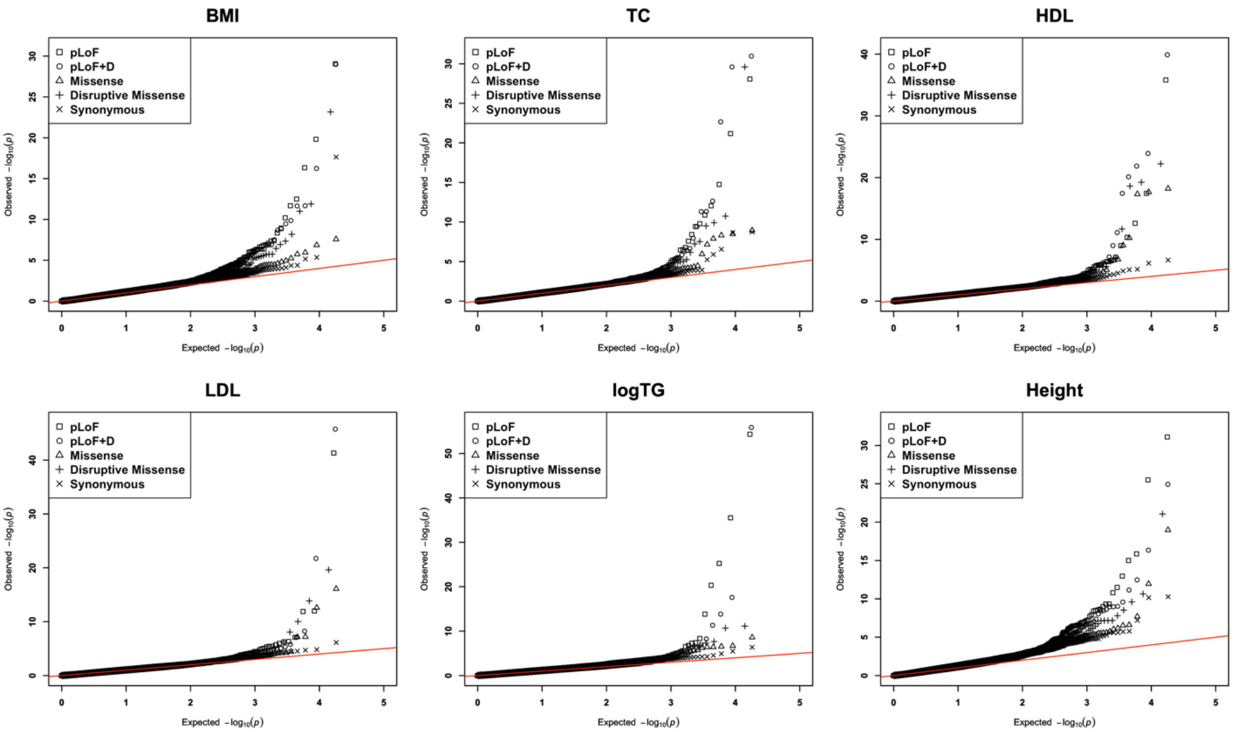

### Supplementary Note

#### Genotype quality control of the UK Biobank whole exome sequencing

UK Biobank (UKB) whole exome sequencing (WES) data were preprocessed following steps out lined in a prior manuscript[1]. VCF files for UK Biobank WES data for 200,643 participants were downloaded[2]. Quality control measures were performed in the following steps[3]. First, variants with Hardy–Weinberg equilibrium $P < 1 \times 10^{-15}$ were removed. Second, any SNV genotype with read depth less than seven reads (DP < 7) and indel genotype with DP < 15 was changed to a no-call. Third, any heterozygous genotype was changed to a no-call if any of the conditions are satisfied as follows: (1) genotype quality < 20, (2) allele balance < 0.15 for SNV and allele balance < 0.20 for indel, and (3) binomial test on allelic balance using allelic depth $P < 1 \times 10^{-3}$. Lastly, variants with more than 10% missing genotypes were excluded.

Rare variant association testing was implemented with the processed genotypes. Common variant association testing was performed with the processed genotype data with an additional minor allele frequency filter (MAF > 1%).

#### Genotype quality control of the UK Biobank whole genome sequencing

The analysis of rare variants used pVCF format files for whole genome sequencing (WGS) data of 200,004 UK Biobank participants (UK Biobank Field #24304) were used following the same quality control procedure in a previous study of UK Biobank WGS data[4]. All variants were kept that had pass indicated by QC label and AAScore greater than 0.5, where AAScore was generated by GraphTyper, the software used by the UK Biobank to perform genotype calling.

Common variant association testing was used with the Plink formatted genotype data process from the pVCF format files for WGS data (UK Biobank Field #24305). Quality control measures were applied to the genotype data using four criteria; (1) variants with minor allele frequency greater than 1% were kept, (2) variants with Hardy–Weinberg equilibrium $P < 1 \times 10^{-6}$ were removed, (3) variants with missing call rates exceeding 0.02 were removed, and (4) samples with missing call rate exceeding 0.05 were removed.

#### Genotype quality control of the All of Us

The analysis of rare variants used version 7.1 Plink formatted short read whole genome sequencing data that includes SNPs and indel variants that are within the exon regions provided by All of Us (srWGS: Exome)[5].

The analysis of common variants used version 7.1 Plink formatted short read whole genome sequencing data including SNPs and indel variants that are frequent in the All of Us computed subpopulations provided by All of Us (srWGS: ACAF Threshold)[5]. Genotype data was further constrained to only include variants that had minor allele frequency greater than 0.01 in the Admixed American, African, and European computed subpopulations.

#### Proof of Beta of PRS per SD

Assuming a model

$$Y = G\beta + \epsilon,$$

where $Y$ is a vector of observed phenotype values for individuals of a single ancestry and $G$ is a standardized genotype matrix. Under this equation, the true heritability explained by $G$ is defined as

$$h^2 = \frac{var(G\beta)}{var(Y)}.$$

Defining an estimate PRS as $PRS = G\hat{\beta}$ where $\hat{\beta} = (G^TG)^{-1}G^TY$. Then the standardized PRS can be defined as

$$PRS_{Stand} = \frac{G\hat{\beta}}{(var(G\hat{\beta}))^{1/2}},$$

and the standardized response as

$$Y_{Stand} = \frac{Y}{(var(Y))^{1/2}}.$$

The estimated coefficient of the standardized PRS from the linear model $Y_{Stand} = PRS_{Stand}\, b + \epsilon$ is given as

$$\begin{aligned}
\hat{b} &= ({PRS_{Stand}}^T PRS_{Stand})^{-1} {PRS_{Stand}}^T Y_{Stand} \\
&= \left(\frac{\hat{\beta}^T G^T G\hat{\beta}}{var\left(G\hat{\beta}\right)}\right)^{-1} \frac{\hat{\beta}^T G^T}{(var(G\hat{\beta}))^{\frac{1}{2}}} \frac{Y}{(var(Y))^{\frac{1}{2}}} \\
&= \left(\frac{var\left(G\hat{\beta}\right)}{var(Y)}\right)^{\frac{1}{2}} (Y^T G(G^TG)^{-1}G^TG(G^TG)^{-1}G^TY)^{-1}Y^TG(G^TG)^{-1}G^TY \\
&= \left(\frac{var\left(G\hat{\beta}\right)}{var(Y)}\right)^{\frac{1}{2}} \\
&= (h^2)^{\frac{1}{2}}
\end{aligned}$$

**Ancestry Adjusted PRS**

We standardize the PRS distributions for each PRS constructed, RICE-CV, RICE-RV, RICE, or conventional common variant methods, using a regression-based method to adjust for differences in distributions across ancestries[6]. The standardization involves two steps: mean adjustment and variance adjustment.

1. Mean adjustment

Linear regression is conducted of the raw PRS against the to five principal components (PC):

$$PRS_i = \alpha_0 + \alpha_1 PC_{i1} + \alpha_2 PC_{i2} + \cdots + \alpha_5 PC_{i5} + {\epsilon_i}^{mean}.$$

The residuals $r_i$ of the raw PRS accounting for mean differences in the PRS distributions across ancestry are computed as:

$$r_i = PRS_i - \hat{\alpha}_0 - \hat{\alpha}_1 PC_{i1} - \hat{\alpha}_2 PC_{i2} - \cdots - \hat{\alpha}_5 PC_{i5}.$$

2. Variance adjustment

Using the square residuals ${r_i}^2$ as a proxy for the variance of the PRS distribution, a second linear regression is ran:

$$r_i{}^2 = \gamma_0 + \gamma_1 PC_{i1} + \gamma_2 PC_{i2} + \cdots + \gamma_5 PC_{i5} + \epsilon_i{}^{var}.$$

Then the final ancestry-adjusted PRS for each individual $i$ was computed as:

$$PRS_i{}^{adj} = \frac{r_i}{\sqrt{\hat{\gamma}_0 + \hat{\gamma}_1 PC_{i1} + \hat{\gamma}_2 PC_{i2} + \cdots + \hat{\gamma}_5 PC_{i5}}}.$$

The resulting distribution of the standardized PRS has mean 0 and variance 1 within each ancestry, ensuring that PRS is accurately modeled regardless of ancestry.
